# Lipid peroxidation induced ApoE receptor-ligand disruption as a unifying hypothesis underlying sporadic Alzheimer’s disease in humans

**DOI:** 10.1101/2021.07.05.21259649

**Authors:** Christopher E. Ramsden, Gregory S. Keyes, Elizabeth Calzada, Mark S. Horowitz, Daisy Zamora, Jahandar Jahanipour, Andrea Sedlock, Fred E. Indig, Ruin Moaddel, Dimitrios Kapogiannis, Dragan Maric

## Abstract

**Background:** Sporadic Alzheimer’s disease (sAD) lacks a unifying hypothesis that can account for the lipid peroxidation observed early in the disease, enrichment of ApoE in the central core of neuritic plaques, the hallmark plaques and tangles, and the selective vulnerability of entorhinal-hippocampal structures.

**Objective:** We hypothesized that (1) high expression of ApoER2 (receptor for ApoE and Reelin) could help explain the selective anatomical vulnerability; and (2) lipid peroxidation of ApoE and ApoER2 contributes to sAD pathogenesis, by disrupting ApoE delivery and Reelin-ApoER2 signaling cascades.

**Methods:** We conducted *in vitro* biochemical experiments, single-marker immunohistochemistry (IHC), and multiplex fluorescence-IHC (MP-IHC) in postmortem specimens from 26 individuals who died cognitively normal, with Mild Cognitive Impairment or with sAD.

**Results:** In biochemical experiments, Lys- and His-enriched peptides within the binding domains of ApoE and ApoER2 and their corresponding recombinant proteins, were susceptible to attack by reactive lipid aldehydes, generating lipid-protein adducts and crosslinked ApoE-ApoER2 complexes. Using *in situ* hybridization alongside IHC and MP-IHC, we observed that: (1) ApoER2 is strongly expressed in terminal zones of the entorhinal-hippocampal ‘perforant path’ projections that underlie memory; (2) ApoE and lipid aldehyde-modified ApoE, Reelin, ApoER2 and several downstream components of Reelin-ApoER2 signaling cascades accumulated in the immediate vicinity of neuritic plaques in perforant path terminal zones in sAD cases; and (3) several ApoE/Reelin-ApoER2 pathway markers—including the ApoER2 ligand binding domain, Disabled homolog-1 (Dab1), and Thr19-phosphorylated PSD95 (marker of synaptic disassembly)—were higher in sAD cases than controls and positively correlated with histological progression and cognitive deficits.

**Conclusion:** Results provide proof-of-concept that ApoE and ApoER2 are vulnerable to lipid aldehyde induced adduction and crosslinking and demonstrate derangements in multiple ApoE/Reelin-ApoER2-Dab1 axis components in perforant path terminal zones in AD. Findings provide the foundation for a unifying hypothesis implicating lipid peroxidation of ApoE particles and ApoE receptors in sAD in humans.

## INTRODUCTION

Sporadic Alzheimer’s disease (sAD), which accounts for >95% of AD cases,[1] lacks effective, disease-modifying treatments. While the etiology of familial AD—including the genetic source of increased amyloid-beta (Aβ) synthesis—is well-understood, the mechanisms underlying sAD are more complex and the initiating molecular lesions are unknown.[2–5] The early stages of sAD are pathologically characterized by degeneration of the neural circuitry that underlies memory formation, including (1) neurons in superficial layers of the entorhinal cortex, (2) their entorhinal-hippocampal ‘perforant path’ projections, and (3) terminal zones of these projections within the molecular layer of the dentate gyrus and cornu ammonis (CA)[6–17] (**Fig 1**), here referred to as the entorhinal-hippocampal memory system. Identification of the molecular determinants underlying this selective anatomical vulnerability may provide clues to the origins of sAD.

**Fig 1.**
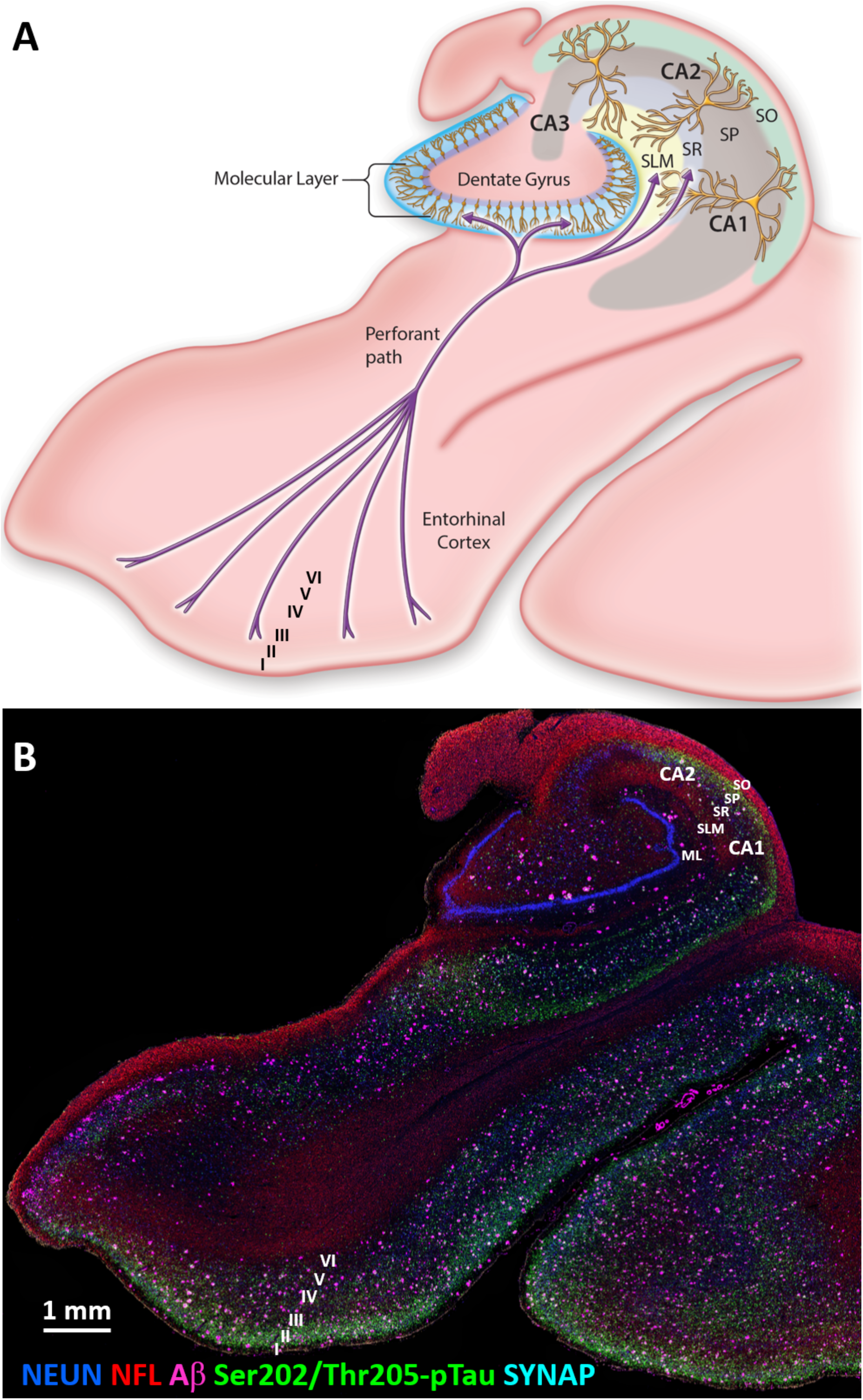
Vulnerability of the entorhinal-hippocampal memory system in sporadic Alzheimer’s disease. (**A**) Schematic diagram illustrates the neural circuitry that underlies memory formation, which includes neurons in Layers II/III of the entorhinal cortex, their entorhinal-hippocampal ‘perforant path’ projections, and the terminal arborization fields of these projections in the dentate gyrus and the cornu ammonis. Degeneration of these entorhinal-hippocampal structures occurs early in sporadic AD (sAD), however the molecular determinants underlying this anatomical vulnerability are unknown. Reelin, which is transported by perforant path axons and secreted by its terminal arbors into the molecular layers of the dentate gyrus and CA1-2 subregions of the hippocampus, is required for synapse formation. Lipid cargo transported by ApoE is also required to shape and protect synapses. ApoE/Reelin-ApoER2-Dab1 pathway activation plays a central role in memory formation in experimental models. Here we propose that high ApoER2 expression, and lipid peroxidation-induced ApoE/Reelin-ApoER2-Dab1 pathway disruption, account for the selective vulnerability of entorhinal-hippocampal structures in AD. (**B**) Multiplex fluorescence IHC image composite in the corresponding sAD case shows the anatomical distribution of the Aβ-positive plaques and Ser202/Thr205-pTau positive tangles that define sAD, with neuronal soma (NEUN), axons (NFL) and synapses (SYNAP) labeled for cytoarchitectural context. CA, cornu ammonis; ML, molecular layer of dentate gyrus; SO, stratum oriens; SP, stratum pyramidale; SR, stratum radiatum; SLM, stratum lacunosum-moleculare; NEUN, neuronal nuclear/soma antigen; NFL, neurofilament light chain; pTau, phosphorylated Tau; SYNAP, synaptophysin.

ApoE is the main apolipoprotein that supplies neurons with the cholesterol and phospholipids required to synthesize and remodel synaptic membranes (**Fig 2A-D**).[18–20] ApoE is enriched in the central core of neuritic plaques [21, 22], and APOE variants are the strongest genetic risk factor for sAD.[23–26] Reelin is a large, secreted glycoprotein that is highly expressed in the entorhinal-hippocampal memory system.[27–29] Reelin signaling shapes and protects synapses by stabilizing actin and microtubule cytoskeletons, and postsynaptic receptor complexes (**Fig 2E-H**).[30–35] In experimental models, Reelin strongly suppresses Tau phosphorylation,[31, 36] and delays Aβ fibril formation.[37, 38] Thus, disruption of Reelin signaling may contribute to genesis of the neurofibrillary tangles and amyloid plaques that define sAD. It is noteworthy that ApoE and Reelin are both ligands for two synaptic ApoE receptors—ApoE receptor type 2 (ApoER2) and very low-density lipoprotein receptor (VLDLR)[39–41] (**Fig 2**)—that promote dendritic arborization and regulate memory formation in rodent models.[35, 42–46] Therefore, it is not surprising that disruption of either ApoE-mediated delivery of lipids [18, 20, 47] or Reelin signaling [35] leads to synapse dysfunction or loss in experimental models.

**Fig 2.**
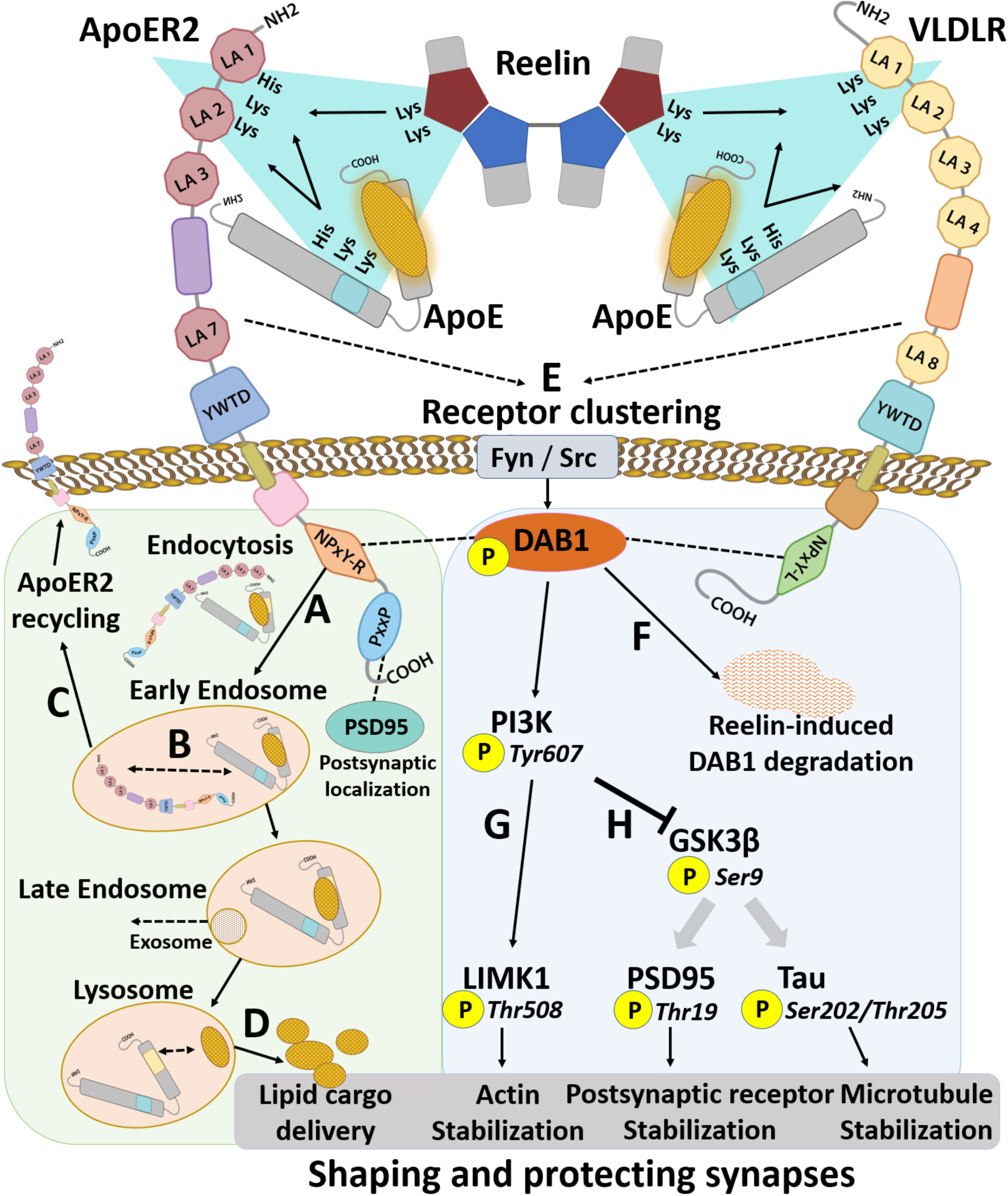
Molecular pathways central to cytoskeletal and synaptic integrity converge at the ApoE/Reelin-ApoE receptor interface. Human versions of two synaptic ApoE receptors—ApoER2 and VLDLR—contain double-Lys motifs between LDL receptor type A (LA) repeats 1 and 2. Reelin and ApoE use double-Lys motifs to bind to ApoER2 and VLDLR. Convergence of these receptors and ligands (**teal triangles**) governs both ApoE-mediated delivery of lipids, and Reelin-ApoER2-Disabled homolog 1 (Dab1) signaling cascades that are essential for cytoskeletal and synaptic integrity. **ApoE cascade** (**A-D, light green shading**): Upon receptor binding, lipid-loaded ApoE particles are internalized (**A**) and trafficked to the early endosome where acidic (pH 6) conditions induce ApoE-ApoE receptor dissociation (**B**) followed by rapid recycling of ApoER2 to the cell surface (**C**). ApoE is trafficked to the late endosome (pH 5.5) and the lysosome (pH<5), which releases the lipid cargo (**D**) that is required to shape synaptic membranes. **Reelin cascade (E-H, light blue shading):** Reelin binding evokes ApoE receptor clustering (**E**), leading to phosphorylation, activation and proteasomal degradation of Dab1 (**F**), and lysosomal degradation of ApoER2 (not shown). Ensuing activation of the Dab1-PI3K-LIMK1-arm of the Reelin cascade stabilizes the actin cytoskeleton (**G**). Reelin-induced Dab1-PI3K phosphorylation inhibits GSK3β by inducing its phosphorylation (**H**), which in turn inhibits Thr19-phosphorylation of PSD95 (faded arrow) and Ser202/Thr205-phosphorylation of Tau (faded arrow) to stabilize postsynaptic receptor complexes and the microtubule cytoskeleton, respectively. Efficient function of these ApoE/Reelin-ApoE receptor pathways is needed to deliver the lipid cargo and to activate the signaling cascades that shape and strengthen synapses.

A marked increase in lipid peroxidation is present even in the earliest stages of sAD.[48–58] Lipid-loaded ApoE particles and the human hippocampus are enriched in polyunsaturated phospholipids that are vulnerable to peroxidation.[20, 59–62] However, specific mechanisms and targets linking lipid peroxidation to AD pathogenesis in general, and mechanisms involving ApoE in particular, are not yet well-understood. Lys and His residues are uniquely vulnerable to attack by aldehydic products of lipid peroxidation [63, 64]. Remarkably, multiple double-Lys and His-enriched sequences within the binding domains of ApoE, Reelin, VLDLR, and human ApoER2 converge at the synaptic ApoE-ApoE receptor interface (depicted by teal triangles in **Fig 2**), where they regulate both ApoE-mediated delivery of lipids, and Reelin-ApoER2-Disabled homolog 1 (Dab1) signaling cascades that are essential for cytoskeletal and synaptic integrity [65–70]. These double-Lys and His-enriched sequences may be consequential targets of lipid peroxidation. Intriguingly, the double-Lys and His-enriched sequence within the first two LDL receptor class A (LA) repeats in human ApoER2 (ApoER2 LA1-2) is absent in mice and rats,[71] suggesting that this putative vulnerability to lipid peroxidation would not be recapitulated by rodent models.

To date, no single hypothesis for sAD has been proposed that can account for the: (1) selective anatomical vulnerability of entorhinal-hippocampal structures; (2) the major role of ApoE (reflected by the genetic link to APOE variants and enrichment of ApoE in the central core of neuritic plaques); (3) hallmark amyloid plaques and neurofibrillary tangles; and (4) evidence of lipid peroxidation in the earliest stages of sAD. Here, we address this gap by proposing a unifying hypothesis wherein the convergence of the lipid cargo transported by ApoE, together with these double-Lys and His-enriched sequences, at the ApoE-ApoE receptor interface creates a microenvironment that is highly vulnerable to lipid peroxidation, and that ensuing disruptions in both ApoE internalization and Reelin-ApoER2-Dab1 signaling trigger a disease-cascade that ultimately manifests as sAD. This ApoE-ApoER2 peroxidation cascade hypothesis, which proposes that high ApoER2 expression and the continuous need for Reelin/ApoE-ApoER2-Dab1 pathway activation to form memories, are the molecular features that account for the anatomical vulnerability of entorhinal-hippocampal structures, is summarized in **Fig 3**, and is contrasted with the amyloid cascade hypothesis in **Table S1**.

**Fig 3.**
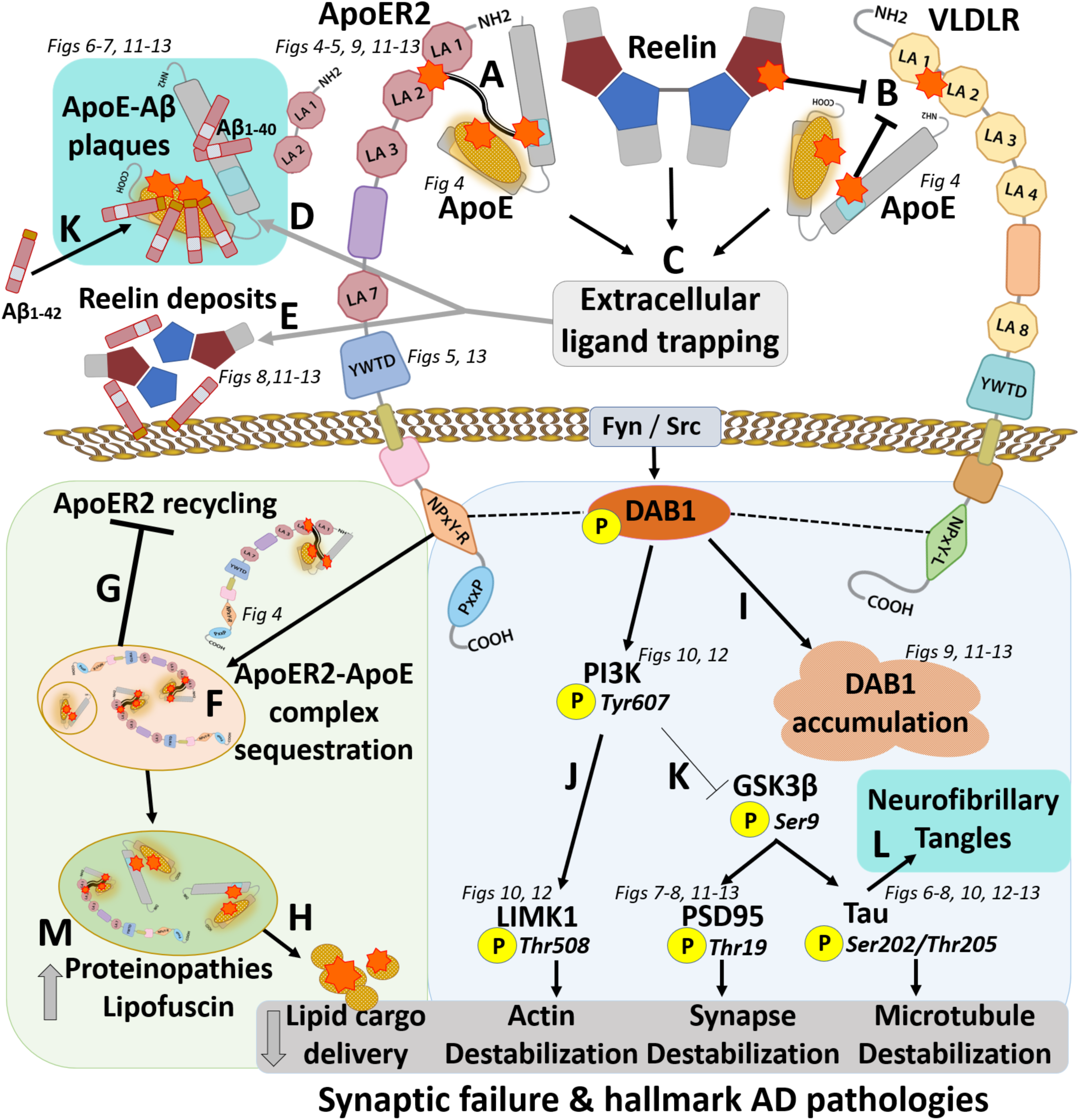
Lipid peroxidation-induced ApoE/Reelin-ApoE receptor-Dab1 axis disruption as a unifying hypothesis underlying sporadic Alzheimer’s disease. As shown in Fig 2, the ApoE cascade and Reelin cascade are depicted by light green and light blue shading, respectively. Aldehydic products of lipid peroxidation (orange stars) derived from lipid-loaded ApoE particles attack Lys and His-enriched sequences within ApoER2, VLDLR, and ApoE, generating lipid-protein adducts and crosslinked ApoE-ApoE receptor complexes. Crosslinks (**A**) and adducts (**B**) compromise ApoE receptor-ligand binding (**C**), leading to extracellular trapping of ApoE (**D**) and Reelin (**E**). Adducts and crosslinks disrupt ApoE-ApoER2 complex dissociation in early endosomes (**F**), thereby disrupting ApoER2 recycling (**G**), and delivery of the lipid cargo needed to remodel synaptic membranes (**H**). Impaired Reelin binding disrupts proteasomal degradation of Dab1, and lysosomal degradation of ApoER2, leading to localized accumulation of Dab1 (**I**) and ApoER2 (not shown). Ensuing compromise of the Reelin-Dab1-PI3K cascade destabilizes the actin cytoskeleton (**J**) and activates GSK3β by decreasing its phosphorylation (thin inhibition sign) (**K**). GSK3β-mediated phosphorylation of PSD95 and Tau destabilizes postsynaptic receptor complexes and the microtubule cytoskeleton, and promotes neurofibrillary tangle formation (**L**). Extracellular trapping of peroxidized ApoE (**C**) provides a nidus for Aβ deposition and oligomerization, leading to ApoE-Aβ plaque formation (**K**). Lysosomal accumulation of crosslinked lipid-protein polymers exacerbates proteinopathies (**M**). Collective derangements are proposed to account for our study findings and the defining hallmarks of AD (teal shading in **L, K**).

To provide proof-of-concept for the proposed initiating molecular lesions depicted in **Fig 3A-B**, we used *in vitro* studies to demonstrate that these double-Lys and His-enriched sequences within the binding domains of ApoE and human ApoER2, and recombinant ApoE and ApoER2 proteins, are vulnerable to lipid aldehyde-induced adduction and crosslinking. To search for these proposed ApoE/Reelin-ApoER2-Dab1 axis pathologies, we custom-designed and validated antibodies targeting the aforementioned double-Lys and His-enriched sequence within human ApoER2 LA1-2 and lipid peroxidation-modified ApoE (see antibody validation sections in methods/supplementary methods). We then used single-marker immunohistochemistry (IHC), multiplex fluorescence-IHC (MP-IHC) with multi-epitope labeling, and ISH/RNA-protein codetection to label core components of the ApoE/Reelin-ApoER2-Dab1 axis (**Fig 2**), alongside hallmark AD pathologies and cytoarchitectural markers in entorhinal-hippocampal specimens from 26 rapidly autopsied individuals who died cognitively normal, with Mild Cognitive Impairment (MCI), or with AD dementia. We found that: (1) ApoER2 is highly expressed in the neurons and dendritic arbors that are the major synaptic recipients of perforant path projections in humans; (2) ApoER2 LA1-2, ApoE and lipid aldehyde-modified ApoE, Reelin, Dab1 and four downstream Reelin-ApoER2-Dab1 signaling partners accumulate in the immediate vicinity of neuritic plaques; (3) several of these markers are more abundant in sAD cases; and (4) accumulations correlated with histological progression and degree of antemortem cognitive impairment. Among these, ApoER2 LA1-2, ApoE, Reelin, Dab1, and Thr19-pPSD95 (a marker of synaptic disassembly and long-term depression (LTD)[72]) exhibited striking expression patterns within these perforant path target zones, and strong correlations with hallmark AD pathologies and cognitive deficits.

Collective findings identify functional domains of ApoE and ApoER2 that are vulnerable to lipid peroxidation and reveal a pathogenic nexus centered around ApoER2 disruption, evidenced by neuritic plaque-associated accumulation of: (1) ApoER2; (2) its extracellular ligands ApoE and Reelin; and (3) several downstream Reelin/ApoE-ApoER2-Dab1 signaling partners, within the human entorhinal-hippocampal memory system in sAD. These findings identify missing molecular links and provide the basis for a unifying hypothesis that could help explain the selective anatomical vulnerability, genetic and environmental risk factors, neuropathological features, and clinical presentation of sAD in humans.

## MATERIALS AND METHODS

### Biochemical experiments

Peptides, proteins, lipids, aldehydes, and other reagents were obtained as indicated in Table S5. Detailed methods used for antigen conjugate preparation, peptide adduction and pH-dependent reversibility experiments, peptide crosslinking experiments, and protein crosslinking experiments are provided in the supplementary materials and methods (pages 38-46).

### Liquid chromatography with mass spectrometry (LC-MS) and time-of-flight mass spectrometry (TOF-MS) analysis of lipid-peptide adducts and crosslinked peptides

LC-MS analyses were performed on an Agilent 1100 LC system with DAD detector and 6120 quadrupole mass spectrometer, using an Eclipse Plus C18 column (4.6mm x 50mm, 5 µm) for separation. The mobile phase was acetonitrile:water 5:95% (v/v) containing 0.2% formic acid (A) and acetonitrile:water 95:5 (v/v) containing 0.2% formic acid (B) with the following gradient elution program: 5% B from 0-3 min, 75% B from 3-12 min, and 5% B at 12.01 min, with a total runtime of 15 min. The mobile phase flow rate was 0.4 mL/min with injection volume of 15 µl. ESI MS scans with capillary voltage of 4000V, nebulizer pressure at 45psig and gas flow at 12 L/min were analyzed using ChemStation software (Agilent). The ApoE peptide + ApoER2 LA1-2 peptide 4-ONE crosslink experiment was characterized on a Triple TOF^TM^5600+ system with Duo-Spray^TM^ion source (Sciex, Framingham, MA) in positive (ESI) mode with the following settings: ion spray voltage, 5500 V; turbo spray temperature, 550°C; declustering potential 60 V; collision energy 10 eV; gas 1, gas 2 and curtain gas were 60, 60 and 20 psi respectively. The sample, dissolved in 10 mM ammonium acetate was directly infused using the on-board syringe pump at a flow rate of 10 µL/min. MS scans were collected across various mass ranges between *m/z*= 500 to 3800 and analyzed with Sciex OS and Analyst software (Sciex).

### Custom antibody generation and validation

A rabbit polyclonal IgG antibody raised against the ApoER2 LA1-2 domain was custom-generated and affinity purified by Vivitide (Gardner, MA, USA). Chicken polyclonal IgY polyclonal antibodies were raised against lipid aldehyde-modified ApoE (made by incubating ApoE monomers with either HNE, CRA, KOdiA-PC, or acetaldehyde). IgY material purified from eggs was subjected to multiple ‘negative’ affinity purifications (with native ApoE columns), sequentially repeated until no further antibodies bound to the native ApoE columns (determined by an OD of <0.1). These were followed by ‘positive’ affinity purification using lipid aldehyde-modified ApoE specific columns (Vivitide). Rat monoclonal antibodies were raised against lipid peroxidation-modified ApoE by incubating ApoE monomers with Cu-oxidized 1-palmitoyl-2-arachidonoyl-sn-phosphatidylcholine (PAPC), a phospholipid that it is particularly enriched in hippocampus.[60] Rat antibodies were custom-generated and affinity purified by Precision Antibody (Columbia, MD, USA). Two clones (15E8 [PxPAPC-ApoEa], and 2B7 [PxPAPC-ApoEb]) were selected based on selective immunoreactivity for Cu-oxidized PAPC-modified ApoE by ELISA. Custom-generated antibodies then underwent orthogonal validation using western blot, positive and negative control immunostaining, single-marker IHC, and MP-IHC with multi-epitope labeling using independent antibodies, as described in the supplementary materials and methods (page 42) and **Extended Fig 2.1**. The ApoER2 LA1-2 antibody was also validated with ISH/RNA-protein codetection. Collective findings using these orthogonal methods indicate that the immunoprobes targeting ApoER2 LA1-2, HNE-ApoE and PxPAPC-ApoE can be used together with commercially available antibodies to selectively label targets needed to investigate the putative pathological nexus (**Fig 3**).

### Characterization of protein crosslinks and antibody specificity by Immunoblot

Recombinant human ApoER2 ectodomain protein (R&D, Cat# 3520-AR-050) was reconstituted at 100µg/mL in sterile PBS. Recombinant human ApoE4 protein (Abcam, Cat# ab50243) was reconstituted at 1mg/mL in water. Recombinant human ApoER2 including C-terminal domain (Origene, TP320903) was supplied at 180µg/mL. Recombinant human VLDLR protein (Sigma, Cat# SRP6469) was reconstituted at 50 µg/mL in water. All protein stocks were aliquoted and stored at -80⁰C. Recombinant proteins were diluted to 25 ng/µL in 1X Laemmli Buffer (Bio-Rad Laboratories, Inc., Cat# 1610747) for SDS-PAGE analysis. Recombinant protein lipid aldehyde reaction mixtures were diluted to 50 ng/µL or 25 ng/µL in 1X Laemmli Buffer and samples were boiled for 5 min at 100⁰C. Ten µL from each sample were loaded onto 8-16% Mini-PROTEAN TGX Precast Protein Gels (Bio-Rad Laboratories, Inc., Cat# 4561106) and proteins were separated by electrophoresis according to the manufacturer’s instructions. Gels were transferred to Immobilon-FL PVDF membrane (MilliporeSigma,Cat# IPFL00010) for low background fluorescence western blotting. Total protein transfer efficiency was determined using AzureRed Fluorescent Total Protein Stain (Azure Biosystems, Inc., Cat# AC2124) immediately after transfer following manufacturer’s instructions with some modification. Membranes were stained for 20 min and washes were performed 3 times for 5min with 100% ethanol. Membranes were kept in dark storage containers protected from light until imaging. Stained membranes were immediately blocked with Azure Fluorescent Blot Blocking Buffer (Azure Biosystems, Inc., Cat# AC2190) for 1 hour and primary antibodies were diluted in fresh blocking buffer and incubated with each membrane overnight. Primary antibodies and dilutions used in these experiments are provided in **Table S5**. Multiplex fluorescent immunoblotting was accomplished by co-incubating rat and mouse or rat and rabbit primary antibodies together in overnight incubations. The next day, membranes were washed 3x for 10 min using PBS containing 0.2% Tween-20 (Sigma-Aldrich, Inc., Cat# P9416). AzureSpectra fluorescent secondary antibodies were applied after washes at a 1:10,000 dilution in fluorescent blocking buffer and incubated for 1 hr. Membranes were washed 3x for 10 min using PBS containing 0.2% Tween-20, and 1x for 5 min using PBS. PVDF membranes were dried by placing on Kimwipes and stored in a drawer until completely dry. Blots were imaged using the smart scan function of the Sapphire Biomolecular Imager (Azure Biosystems, Inc.).

### Postmortem specimens

6 µm-thick coronal medial temporal lobe paraffin sections at the level of the body of the hippocampus were obtained from a total of 29 cases spanning the clinicopathological spectrum of sAD, from the Brain and Body Donation Program (BBDP) at the Banner Sun Health Research Institute (http://www.brainandbodydonationprogram.org) (**Table 1** and **Suppl Tables 2-4**). To mitigate limitations due to tissue degradation, BBDP employed a rapid on-call autopsy team to achieve short PMI (mean 3h).

Standardized fixation procedures were employed with 1 cm sections fixed in 10% formalin for 48 hours. All BBDP subjects provided written consent for study procedures, autopsy and sharing of de-identified data prior to enrollment. The study and its consenting procedures were approved by the Western Institutional Review Board of Puyallup, Washington. The Banner BBDP population has been extensively described by Beach et al.[73–76] Briefly, most BBDP donors were enrolled as cognitively normal volunteers residing in the retirement communities near Phoenix, Arizona, USA. Following provision of informed consent, donors received standardized medical, neurological, and neuropsychological assessments during life. Neuropathological and cognitive examinations and endpoints captured by BBDP are detailed in a previous publication.[73] Ante-mortem Mini-Mental Status Exam (MMSE) test had the least missing data and was the main cognitive endpoint of for the present study.

### Antibody selection and multi-epitope labeling approach

The antibodies and ISH probes used in the present study targeted cytoarchitectural markers, hallmark AD pathologies, and core components of ApoE/Reelin-ApoER2-Dab1 signaling pathways (**Figs 3-4**). A multi-epitope strategy was used to label ApoER2, native ApoE, and lipid-peroxidation modified ApoE (**Extended Fig 2**). IHC-validated antibodies were used whenever available. Custom-designed antibodies were validated using an orthogonal approach as described above. MP-IHC allowed us to compare staining patterns for several independent antibodies targeting different epitopes within targeted proteins, to include several targets within a given molecular pathway of interest, and to concurrently label multiple cytoarchitectural markers. MP-IHC was used together with single-marker IHC in serial sections to assist in validation, and to provide spatial, morphological, and cytoarchitectural context for observations made using single antibodies.

**Fig 4.**
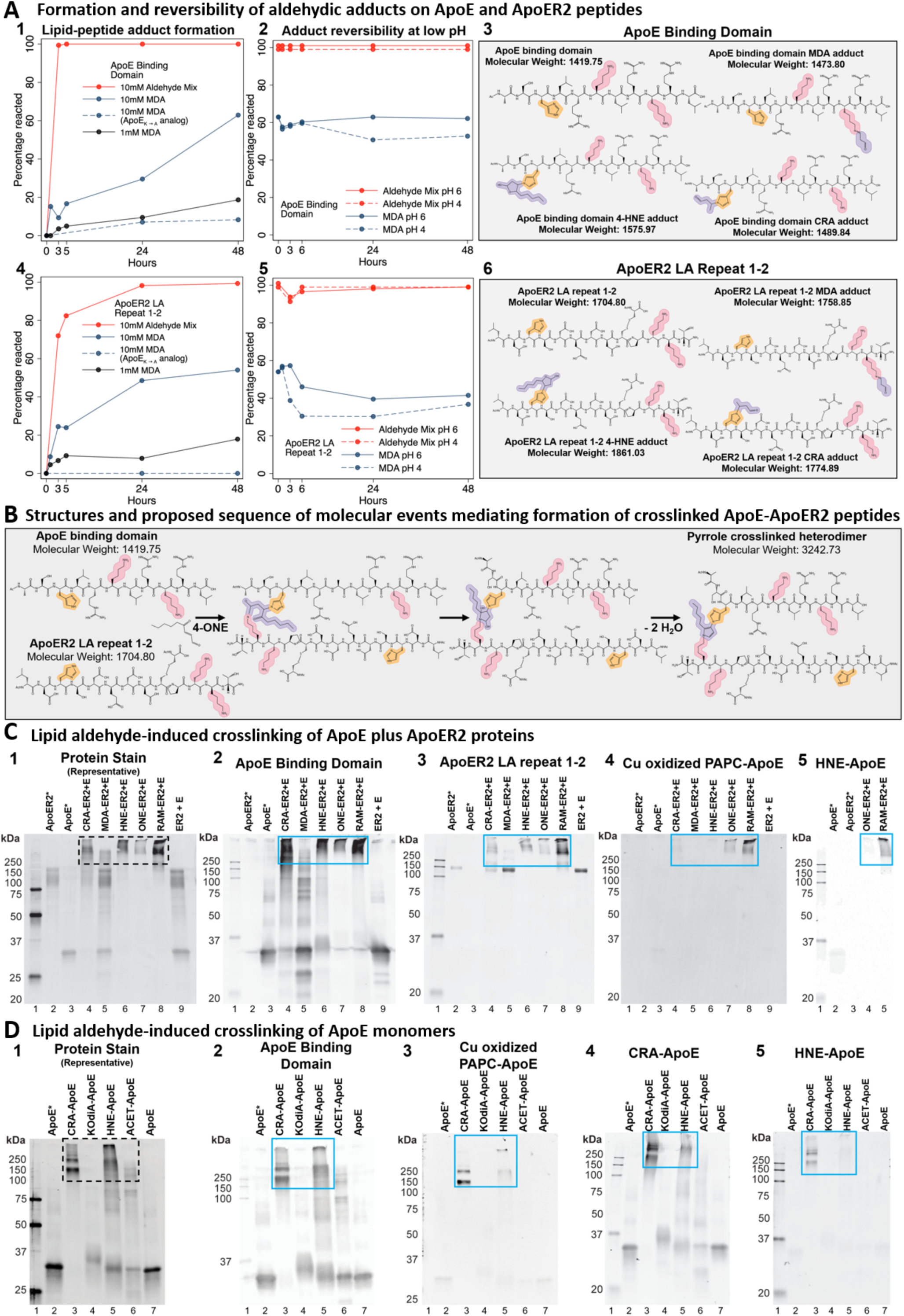
Lipid aldehyde-induced adduction and crosslinking of ApoE and ApoER2 peptides and proteins detected by mass spectrometry, high molecular weight protein migration, and lipid peroxidation-specific antibodies. Lys and His-enriched peptides within the binding regions of ApoE (**A_1-3_**) and LA repeats 1-2 of ApoER2 (**A_4-6_**), and their respective analogs absent the double-Lys motifs (dashed lines in **A_1-2, 4-5_**), were incubated with MDA or Reactive Aldehyde Mix (RAM) for 48 hours, followed by 48 hours of adduct reversal under conditions modeling the early endosome (pH 6) or lysosome (pH 4). Time courses for adduct formation and pH-dependent reversal were monitored by LC-MS. Yellow, pink and purple shading in **A_3_, A_6_** and **B** designate the reactive amines in His and Lys, and aldehydic adducts, respectively. Panel **B** depicts the structures and proposed sequence of molecular events mediating formation of crosslinked ApoE-ApoER2 peptides. Following incubation of ApoE and ApoER2 with 4-ONE, an acid-stable crosslinked ApoE-ApoER2 peptide was observed as its triple charged ion (MW = 3242, [(M+3H^+^)/3= 1081] and its quadruple charged ion ([(M+4H^+^)/4= 811] (**Extended Fig 4.1**) corresponding to a pyrrole crosslink. Parallel experiments with peptide analogs established that peptide crosslinking was dependent on His and Lys residues. Lipid aldehyde-induced crosslinking of ApoE plus ApoER2 proteins was detected by fluorescence immunoblotting against ApoE (**C_2_**), ApoER2 (**C_3_**), Cu-oxidized PAPC modified-ApoE (**C_4_**), and HNE-ApoE (**C_5_**). 250ng of ApoER2 (lane 2) and ApoE4 (lane 3) were loaded per gel to compare with 500ng of protein from each combined lipid aldehyde reaction. Crotonaldehyde (CRA-ER2+E, Lane 4), malondialdehyde (MDA-ER2+E, Lane 5), 4-hydroxy-2-nonenal (HNE-ER2+E, lane 6), 4-oxo-2-nonenal (ONE-ER2+E, lane 7), Reactive Aldehyde Mixture (RAM-ER2+E, lane 8), and native ApoER2 and ApoE proteins co-incubated without aldehydes (lane 9, ER2 + E) are represented in immunoblots **C_2-5_**. ApoER2 (lane 2) and ApoE4 (lane 3), 4-oxo-2-nonenal (ONE-ER2+E, lane 4), Reactive Aldehyde Mixture (RAM-ER2+E, lane 5) are represented in immunoblot **C_3_**. Lipid aldehyde-induced crosslinking of ApoE monomers was detected by fluorescence immunoblotting against ApoE (**D_2_**), Cu-oxidized PAPC modified-ApoE (**D_3_**), CRA-ApoE (**D_4_**), and HNE-ApoE (**D_5_**). 250ng of ApoE was loaded per gel. Crotonaldehyde (CRA-ApoE, Lane 3), KOdiA-PC (KOdiA-ApoE, Lane 4), 4-hydroxy-2-nonenal (HNE-ApoE, lane 5), acetaldehyde (ACET-ApoE, lane 6), and native ApoE protein co-incubated without aldehydes (lane 7, ApoE) are represented in immunoblots **D_1-5_**. **C_1_** and **D_1_**, total protein per membrane was detected by AzureRED total protein stain. The blue boxes denote higher molecular weight proteins detected. Western blots are representative of experiments performed three times. Results were also confirmed with additional ApoER2 and ApoE antibodies (**Extended Fig 4.2-4.3**). ApoER2* indicates that the ApoER2 ectodomain protein was used; ApoE* indicates that the ApoE4 monomer was used to prevent confounding by non-aldehydic disulfide-linked dimerization.

### Single-marker immunohistochemistry (IHC)

Single-marker IHC was performed by Histoserv (Gaithersburg, MD, USA). Combined blocks containing temporal and/or frontal cortex from sAD cases and non-AD controls were sectioned into 6μm-thick coronal sections. Optimal immunostaining conditions were then empirically determined using an incremental heat induced epitope retrieval (HIER) method using 10mM Na/Citrate pH6 and/or TRIS/EDTA pH9 buffer under following heating conditions: 70℃/10 min, 70℃/20 min, 70℃/40 min. Selected antibodies with suboptimal HIER treatment of up to 40 minutes underwent additional retrieval rounds using formic acid (88%, 20 min) or proteinase K (Dako S3020) for 5-10 min at room temperature (RT). Secondary antibodies used for single IHC chromogenic staining were appropriately matched to the host class/subclass of the primary antibody. These combined sAD plus Control blocks were used to generate negative controls (comparing staining results versus pre-immune serum and with the primary antibody omitted) and positive controls (comparing staining results in temporal or frontal cortex specimens from confirmed AD cases with known region-specific Aβ and tangle scores versus non-AD controls). These empirically determined optimal conditions were then used to immunolabel sets of entorhinal-hippocampal slides using IHC and MP-IHC (as described below). Briefly, for IHC, sections were first deparaffinized using standard Xylene/Ethanol/Rehydration protocol followed by antigen unmasking with 10-40 min HIER step in 10 mM Tris/EDTA buffer pH9 (Tris/EDTA buffer) as described above (see **Table S5** for antibody sources and technical specifications).

### Multiplex Fluorescence Immunohistochemistry and In Situ Hybridization

Multiplex fluorescence immunohistochemistry (MP-IHC) and multiplex fluorescence *in situ* hybridization (MP-ISH) were performed on 6 µm-thick coronal sections that were collected on Leica Apex Superior adhesive slides (VWR, 10015-146) to prevent tissue loss. We completed up to 6 iterative rounds of sequential immunostaining with select antibody panels targeting the ApoE/Reelin-ApoE receptor-Dab1 axis and classical biomarkers of AD pathology, alongside classical cytoarchitectural biomarkers to map brain tissue parenchyma (see **Table S5**). Fluorophore conjugated targets from each round were imaged by multispectral epifluorescence microscopy followed by antibody stripping and tissue re-staining steps to repeat the cycle, as previously described [77], each time using a different antibody panel. MP-ISH was carried out on a subset of sections to screen for ApoER2 mRNA using a custom designed RNA probe and standard RNAscope Multiplex Fluorescent V2 hybridization kits (Advanced Cell Diagnostics, Inc.) per manufacturer’s instructions. Sections probed with RNAscope were subsequently processed with 2 sequential rounds of MP-IHC to facilitate RNA-protein codetection. Briefly, for screening tissues by MP-IHC and/or MP-ISH, the sections were first deparaffinized using standard Xylene/Ethanol/Rehydration protocol followed by HIER antigen unmasking step in Tris/EDTA buffer for 10-15 min using an 800W microwave set at 100% power. Sections intended for MP-ISH were then processed using the RNAscope V2 kit (Advanced Cell Diagnostics, Inc.). All sections to be sequentially processed for iterative MP-IHC screening were first incubated with Human BD Fc Blocking solution (BD Biosciences) to saturate endogenous Fc receptors, and then in True Black Reagent (Biotium) to quench intrinsic tissue autofluorescence. Sections were then immunoreacted for 1 hour at RT using cocktail mixture of immune-compatible antibody panels targeting ApoE/Reelin-ApoE receptor-Dab1 signaling cascade pathways, classical biomarkers of AD pathology, and brain cytoarchitectural biomarkers (see **Table S5** for antibody sources and technical specifications). This step was followed by washing off excess primary antibodies in PBS supplemented with 1 mg/ml bovine serum albumin (BSA) and staining the sections using a 1 µg/ml cocktail mixture of the appropriately cross-adsorbed secondary antibodies (purchased from either Thermo Fisher, Jackson ImmunoResearch or Li-Cor Biosciences) conjugated to one of the following spectrally compatible fluorophores: Alexa Fluor 430, Alexa Fluor 488, Alexa Fluor 546, Alexa Fluor 594, Alexa Fluor 647, IRDye 600LT, or IRDye 800CW. After washing off excess secondary antibodies, sections were counterstained using 1 µg/ml DAPI (Thermo Fisher Scientific) for visualization of cell nuclei. Slides were then coverslipped using Immu-Mount medium (Thermo Fisher Scientific) and imaged using a multispectral epifluorescence microscope (see below). After imaging, tissue bound primary and secondary antibodies were both stripped off the slides after a 5-minute incubation at RT in NewBlot Nitro 5X Stripping buffer (Li-Cor Biosciences) followed by 1-minute additional HIER step in Tris/EDTA buffer. The above processing cycle beginning with re-blocking of tissues in Human BD Fc Blocking solution was repeated and the same sections then incubated using an additional panel of antibodies of interest.

### Multiplex Fluorescence Immunohistochemistry Image Acquisition and Computational Reconstruction

Images were acquired from MP-IHC and/or MP-ISH probed specimen sections using the Axio Imager.Z2 slide scanning fluorescence microscope (Zeiss) equipped with a 20X/0.8 Plan-Apochromat (Phase-2) non-immersion objective (Zeiss), a high resolution ORCA-Flash4.0 sCMOS digital camera (Hamamatsu), a 200W X-Cite 200DC broad band lamp source (Excelitas Technologies), and 8 customized filter sets (Semrock) optimized to detect the following fluorophores: DAPI, Alexa Fluor 430, Alexa Fluor 488 (or Opal 520), Alexa Fluor 546 (or Opal 570), Alexa Fluor 594 (or Opal 620), Alexa Fluor 647, IRDye 680LT (or Opal 690) and IRDye 800CW. Image tiles (600 x 600 µm viewing area) were individually captured at 0.325 micron/pixel spatial resolution, and the tiles seamlessly stitched into whole specimen images using the ZEN 2 image acquisition and analysis software program (Zeiss), with an appropriate color table having been applied to each image channel to either match its emission spectrum or to set a distinguishing color balance. Pseudo-colored stitched images acquired from 1 round of MP-ISH and/or up to 6 rounds of MP-IHC staining and imaging were then exported to Adobe Photoshop and overlaid as individual layers to create multicolored merged composites. Multi-channel images acquired from multi-round IHC staining and repeatedly imaged tissue samples were computationally registered at the subpixel level using affine transformation, and the aligned fluorescence images corrected for photobleaching, autofluorescence, background signals, non-uniform illumination, and spectral bleed-through artifacts, as previously described by Maric et al.[77]

### Regional Annotation and Quantitation

IHC images were uploaded into HALO 3.1 image analysis software (Indica Labs, Corrales, NM). Boundaries of each anatomical region of interest (hippocampus, molecular layer of the dentate gyrus, gray matter of middle temporal gyrus) were identified and annotated using a combination of anatomical landmarks (lateral ventricle, fimbria, fimbria-dentate and hippocampal fissures, collateral sulcus, dentate granule cells, cornu ammonis) and cytoarchitectonic mapping as described by Amunts et al.[78] MP-IHC, which enabled immunolabeling of numerous cytoarchitectural markers in each section (**Table S5**), assisted in the identification of anatomical landmarks such as the dentate granule cell layer and boundaries including white matter tracts and the gray matter-white matter interfaces. Stain positive area as a percentage of each annotated region was quantified using the HALO Area Quantification v2.2.1 module. Plaque-associated objects per mm^2^ within each annotated region were identified and quantified using the HALO Object Colocalization v1.3 module with classifier function enabled. Representative examples depicting applications of these modules are provided in the supplementary methods (page 43).

### Statistical analysis

For each annotated region, differences between sAD, MCI, and Control groups according to each marker were quantified using Kruskal-Wallis tests. Variable transformations (e.g., natural log) were used as necessary. A Spearman’s correlation coefficient between each immunohistochemical marker and each AD endpoint (Braak score, Total Aβ plaque load, MMSE) was calculated. Statistical analyses were conducted in Stata Release 16 (College Station, TX). No adjustments were made for multiple comparisons. Four-panel graphs showing individual data points in each group and their relationships to AD endpoints are provided in **Figs 5-12**.

**Fig 5.**
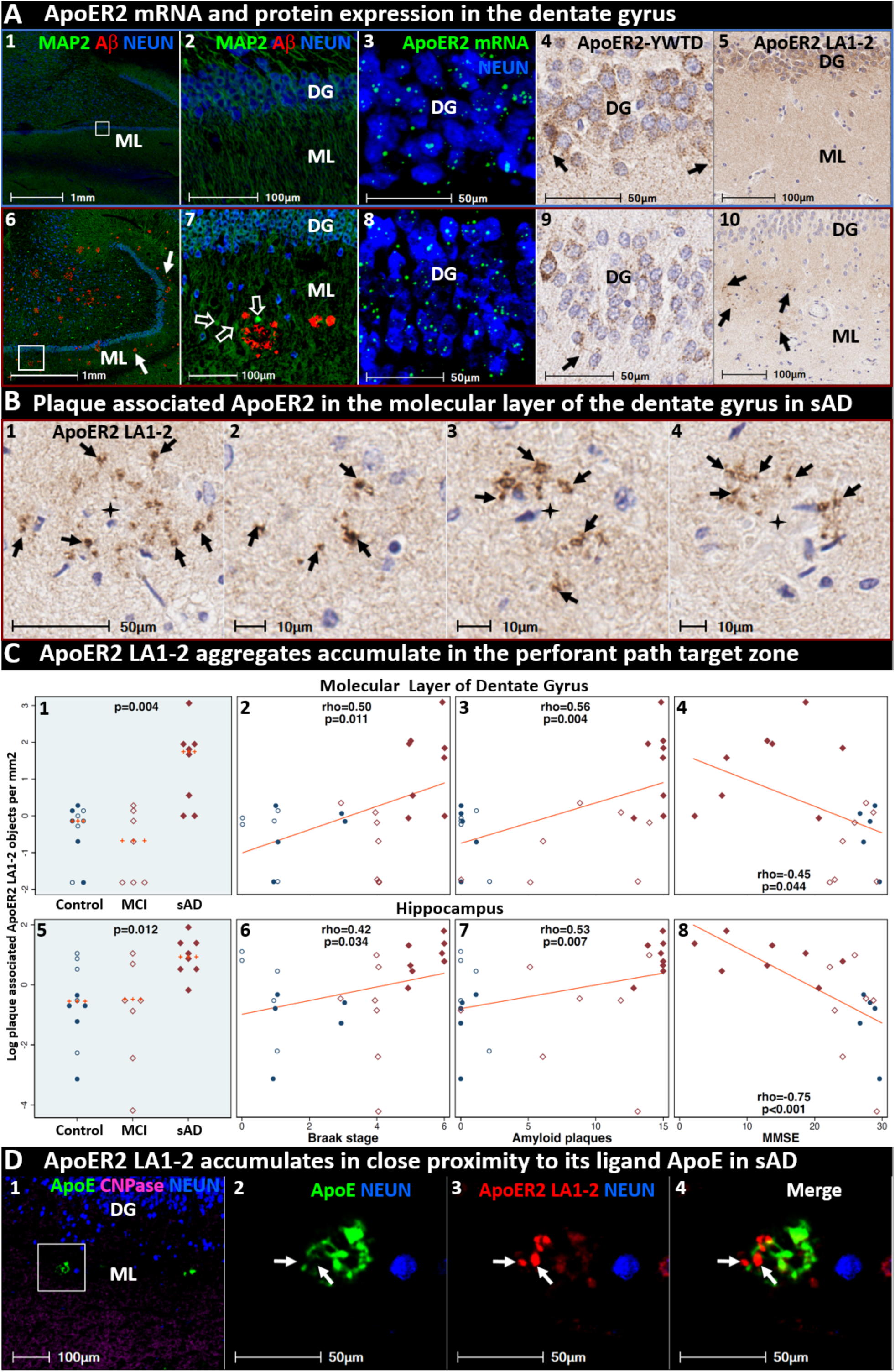
ApoER2 LA1-2 accumulates as discrete aggregates in terminal zones of perforant path projections in sporadic AD. Panels **A_1-5_** and **A_6-10_** are coronal sections from representative non-AD control case and (Braak stage V, ApoE3/3) sAD case, respectively. Panels **A_6-7_** depict MAP2-positive dentate granule cells and their dense dendritic arbors emanating into the molecular layer of the dentate gyrus, with dystrophic dendrites surrounding Aβ plaques in the sAD case (open arrows in **A_7_**). Panels **A_3_** and **A_8_** show ApoER2 mRNA expression in dentate granule cells. Panels **A_4_** and **A_9_** depict ApoER2-YWTD protein domain expression in dentate cells their dendritic projections emanating into the molecular layer (open arrows in **A_4_**, **A_9_**). Panels **A_5_** and **A_10_** depict ApoER2 ligand binding domain (ApoER2 LA1-2) expression within the molecular layer of the dentate gyrus. Controls exhibit diffuse, homogenous ApoER2 LA1-2 expression (**A_5_**). By contrast, AD cases exhibit numerous discrete ApoER2 LA1-2 aggregates throughout the molecular layer (arrows in **A_10_** and **B_1-4_**). In both the molecular layer of dentate gyrus and hippocampus, ApoER2 aggregates accumulate in sAD cases and correlate with histochemical progression and cognitive deficits (**C_1-8_**). MP-IHC labeling (**D_1-4_**) reveals that ApoER2 LA1-2 and ApoE accumulate in proximity to one another (white arrows), but have minimal colocalization, in the vicinity of neuritic plaques. Additional examples of plaque associated ApoER2 LA1-2 aggregates in dentate gyrus and cornu ammonis are provided in **Extended Figs 5-7** and **9-13**. **Panel C**: open and closed blue circles indicate young controls and age-matched controls; open and closed red diamonds indicate MCI cases and sAD cases, respectively. **Abbreviations**: DG, dentate granule cell layer; ML, molecular layer; MAP2, microtubule associated protein-2; NEUN, neuronal nuclear/soma antigen; CNPase, Cyclic-nucleotide-phosphodiesterase.

## RESULTS

### Lipid aldehyde-induced adduction and crosslinking of Lys and His-enriched sequences within ApoE and ApoER2

To determine whether Lys and His-enriched sequences within the binding domains of ApoE and ApoER2 are vulnerable to attack by aldehydic products of lipid peroxidation *in vitro*, ApoE and ApoER2 peptides were incubated with malondialdehyde (MDA) or a mixture of aldehydes. Native ApoE and ApoER2 peptides were observed to be vulnerable to adduction (**Fig 4A_1_, A_4_** solid lines). By contrast, ApoE and ApoER2 peptide analogs lacking double-Lys motifs were resistant to adduct formation (**Fig 4A_1_, A_4_** dashed lines), implying a crucial role for Lys in MDA-induced adduction. To determine the extent of reversibility of ApoE and ApoER2 peptide-aldehyde adducts, pH conditions of the early endosome (pH 6) and lysosome (pH 4) were modeled. Partial, but incomplete, pH-dependent (pH 4>6) reversibility of MDA adducts of ApoE and ApoER2 was observed (**Fig 4A_2_, A_5_**), however adducts formed with the mixture of aldehydes had minimal reversibility.

To determine if these Lys and His-enriched sequences within ApoE and ApoER2 are susceptible to lipid aldehyde-induced crosslinking, both peptides were incubated together with 4-ONE, a reactive 4-ketoaldehyde derived from peroxidation of polyunsaturated fatty acids,[79] and monitored with LC-MS and TOF-MS. Peaks were observed with corresponding molecular weights indicating presence of crosslinked ApoE-ApoER2 heterodimers and intra-chain crosslinks within ApoE and ApoER2 peptides (**Fig 4B** and **Extended Fig 4.1**). Molecular weights of each crosslinked peptide indicated addition of one 4-ONE molecule with the loss of two water molecules, suggesting the formation of pyrrole crosslinks.[80, 81] To determine whether His and/or Lys residues mediated the crosslinking process, the experiments were repeated using combinations of ApoE and ApoER2 analogs lacking Lys or His residues. We observed that both the His and Lys in ApoE, and either the His or Lys in ApoER2, were required to form these crosslinked peptides. To determine if aldehyde-peptide crosslinks can be reversed by acidic conditions modeling the lysosome, crosslinked peptides were incubated at pH 4 for 24 hours. There was no detectable change in crosslinked peptides, further supporting the presence of stable pyrrole crosslinks.[82] The proposed biosynthetic sequence of reactions and the chemical structures including these stable His-Lys pyrrole crosslinks are shown in **Fig 4B**.

### ApoE and ApoER2 proteins are vulnerable to lipid aldehyde-induced crosslinking and formation of high molecular weight ApoE-ApoER2 complexes and non-disulfide linked ApoE multimers

We next sought to determine whether full-length ApoE and the ectodomain of ApoER2 are susceptible to aldehyde-induced crosslinking and the formation of heteromeric ApoE-ApoER2 complexes. To model this *in vitro*, we incubated ApoE and ApoER2 proteins alone, together without reactive aldehydes, or together in combination with either a single lipid aldehyde or a mixture of aldehydes. In samples containing ApoE and ApoER2 alone or together without aldehydes, western blotting with antibodies targeting ApoE and ApoER2 detected only their respective lower molecular weight bands (**Fig 4C_2-3_**). By contrast, in samples containing both proteins incubated with CRA, HNE, ONE or a mixture of reactive aldehydes, higher molecular weight bands were detected by total protein stain as well as by antibodies targeting either ApoE and ApoER2, indicating the presence of aldehyde-crosslinked ApoE and ApoER2 complexes (indicated by high molecular weight bands in **Fig 4C_1_** and **C_2-3_**, respectively). These findings were confirmed with additional antibodies targeting different ApoER2 domains (**Extended Figs 4.2-3**).

To determine whether ApoE monomers are susceptible to aldehyde-induced crosslinking and the formation of (non-disulfide linked) ApoE multimers, we incubated recombinant ApoE protein without reactive aldehydes, or together in combination with single lipid aldehydes. In samples containing ApoE without aldehydes, western blotting with antibodies targeting native ApoE detected only the ≈35kDa molecular weight band corresponding to ApoE monomers (**Fig 4D_2_**). By contrast, in samples with ApoE monomers incubated with CRA or HNE, high molecular weight bands were detected by total protein stain as well as by antibodies targeting native ApoE epitopes, indicating the presence of aldehyde-crosslinked ApoE multimers (**Fig 4D_1-5_**, respectively). These findings were confirmed with additional antibodies targeting different ApoE domains (**Extended Figs 4.3**).

### Antibodies raised against lipid peroxidation-modified ApoE selectively label peroxidation-crosslinked ApoER2-ApoE complexes and aldehyde-crosslinked ApoE multimers

Western blotting with either of two rat monoclonal antibodies raised against Cu-oxidized PAPC (a phospholipid that it is particularly enriched in hippocampus,[59–61] or the chicken polyclonal antibody raised against HNE-modified ApoE monomers, selectivity detected only the high-molecular weight aldehyde-crosslinked heteromeric ApoE-ApoER2 complexes and aldehyde-crosslinked ApoE multimers, with minimal or no detection of native ApoE (**Fig 4C_4-5_, D_3_, D_5_** and **Extended Figs 4.2-4.3**). By contrast, despite multiple ‘negative’ affinity purifications with native ApoE columns, chicken polyclonal antibodies raised against ApoE monomers incubated with crotonaldehyde (CRA-ApoE) (**Fig 4D_4_**), or with KOdiA-PC (KOdiA-ApoE) or acetaldehyde (ACET-ApoE) maintained some reactivity for native ApoE (**Extended Fig 4.3**).

The collective observations in sections 2.1-2.3 provide *in vitro* proof-of-concept that ApoE and ApoER2 are vulnerable to aldehyde-induced adduction and crosslinking, generating (non-disulfide linked) ApoE multimers and heteromeric ApoE-ApoER2 complexes. Such adducts and complexes, if present *in vivo* in entorhinal-hippocampal structures, are predicted to compromise memory formation by disrupting both ApoE internalization and Reelin-ApoE receptor-Dab1 signaling cascades that shape and protect synapses (see **Fig 3**).

Having demonstrated *in vitro* proof-of-concept for the proposed initiating molecular lesions shown in **Figs 3-4**, we next used single-marker IHC, RNA-protein codetection and MP-IHC to search for evidence of ApoER2 expression and ApoE-ApoER2 pathway disruption, in postmortem human dentate gyrus and hippocampus specimens from 26 individuals who died cognitively normal, with MCI, or with AD dementia (**Table 1**).

### ApoER2 LA1-2 accumulates as discrete aggregates in terminal zones of perforant path projections in sAD

ApoER2 is highly expressed in the rodent hippocampus [39] and plays a central role in dendritic arborization and memory in rodent models.[83] Although ApoER2 is known to be expressed in the human brain,[39, 84] little is known about its distribution within specific entorhinal-hippocampal structures. To address this gap, we examined expression and morphology of ApoER2 in the dentate gyrus and hippocampus using single-marker IHC, ISH/RNA-protein codetection, and MP-IHC (**Fig 5**). ISH and IHC revealed abundant ApoER2 mRNA expression and immunolabeling of the midchain beta-propeller domain of ApoER2 protein (ApoER2-YWTD) within somata of granule cells and pyramidal basket cells in the dentate gyrus (**Fig 5A**), and in pyramidal neurons within the CA1-2 regions of the hippocampus. As depicted in **Fig 1**, dendritic arbors emanating from these labeled neurons project into the molecular layer of the dentate gyrus, and the stratum radiatum (SR) and stratum lacunosum-moleculare (SLM) of CA1-2, respectively, where they receive synaptic inputs from the perforant path projections.[12, 15, 85]

Single-marker IHC revealed numerous discrete ApoER2 LA1-2 positive aggregates within the molecular layer of the dentate gyrus (**Fig 5B**), and the stratum pyramidale (SP) and SR subregions of CA1-2 (**Extended Fig 5.1**) in sAD cases, which were rare or absent in non-AD controls. Plaque-associated ApoER2 LA1-2 aggregates in the molecular layer of the dentate gyrus and hippocampus were significantly increased in sAD cases, positively correlated with Braak stage and Aβ plaque load, and inversely correlated with MMSE score (**Fig 5C**). MP-IHC revealed that these ApoER2 aggregates are in the immediate vicinity of, but generally do not directly colocalize with, ApoE aggregates (**Fig 5D**). Antibodies targeting ApoER2-YWTD and other ApoER2 domains labeled neuronal and dendritic architecture within these regions (**Extended Fig 5.2**), however the antibody targeting the ApoER2 LA1-2 domain produced the most robust labeling of these plaque-associated aggregates. Additional examples of ApoER2 LA1-2 aggregates are provided in **Figs 12-13** and **Extended Figs 5-7 & 9-13**.

**Fig 6.**
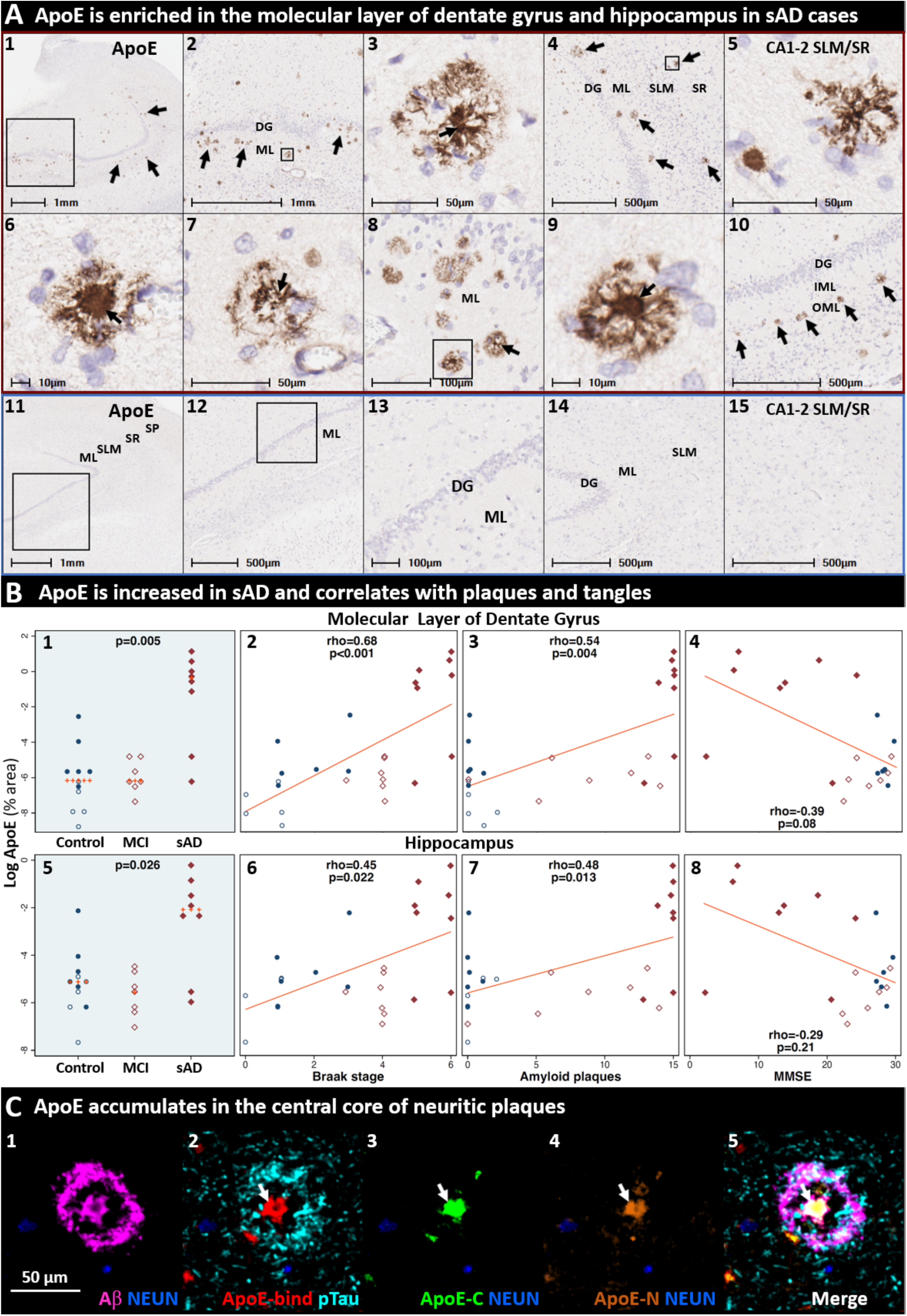
ApoE-enriched plaques accumulate in the terminal zones of perforant path projections in sAD. Coronal sections of the molecular layer of the dentate gyrus and hippocampus from a representative (Braak stage V, APOE3/3) sAD case (**A_1-10_**) and a non-AD control case (**A_11-15_**) were stained with an anti-ApoE antibody. ApoE immunoreactive plaques were observed in the molecular layer of the dentate gyrus (arrows in **A_1-4_, A_6-10_**) and the molecular layers of CA1-2 (**A_5_**) in sAD cases, with minimal staining in controls. ApoE expression was increased in sAD cases and positively correlated with Braak stage and Aβ plaque load (but not MMSE score) in both annotated regions (**B_1-8_**). MP-IHC staining confirmed that antibodies targeting ApoE-binding domain (**C_2_**), and ApoE C-terminal (**C_3_**) and N-terminal (**C_4_**) domains strongly label the central core of neuritic plaques (white arrows in **C_2-5_**). Additional cytoarchitectural, spatial, and morphological context is provided in **Extended Fig 6.2**. Red and blue boxed panels in **A_1-10_** indicate and **A_11-15_** indicate sAD cases and non-AD controls, respectively. **Panel B**: open and closed blue circles indicate young controls and age-matched controls; open and closed red diamonds indicate MCI cases and sAD cases, respectively. **Abbreviations**: DG, dentate granule cell layer; ML, molecular layer; OML, outer molecular layer; IML, inner molecular layer; SP, stratum pyramidale; SR, stratum radiatum; SLM, stratum lacunosum-moleculare; CA, cornu ammonis; NEUN, neuronal nuclear/soma antigen.

**Fig 7.**
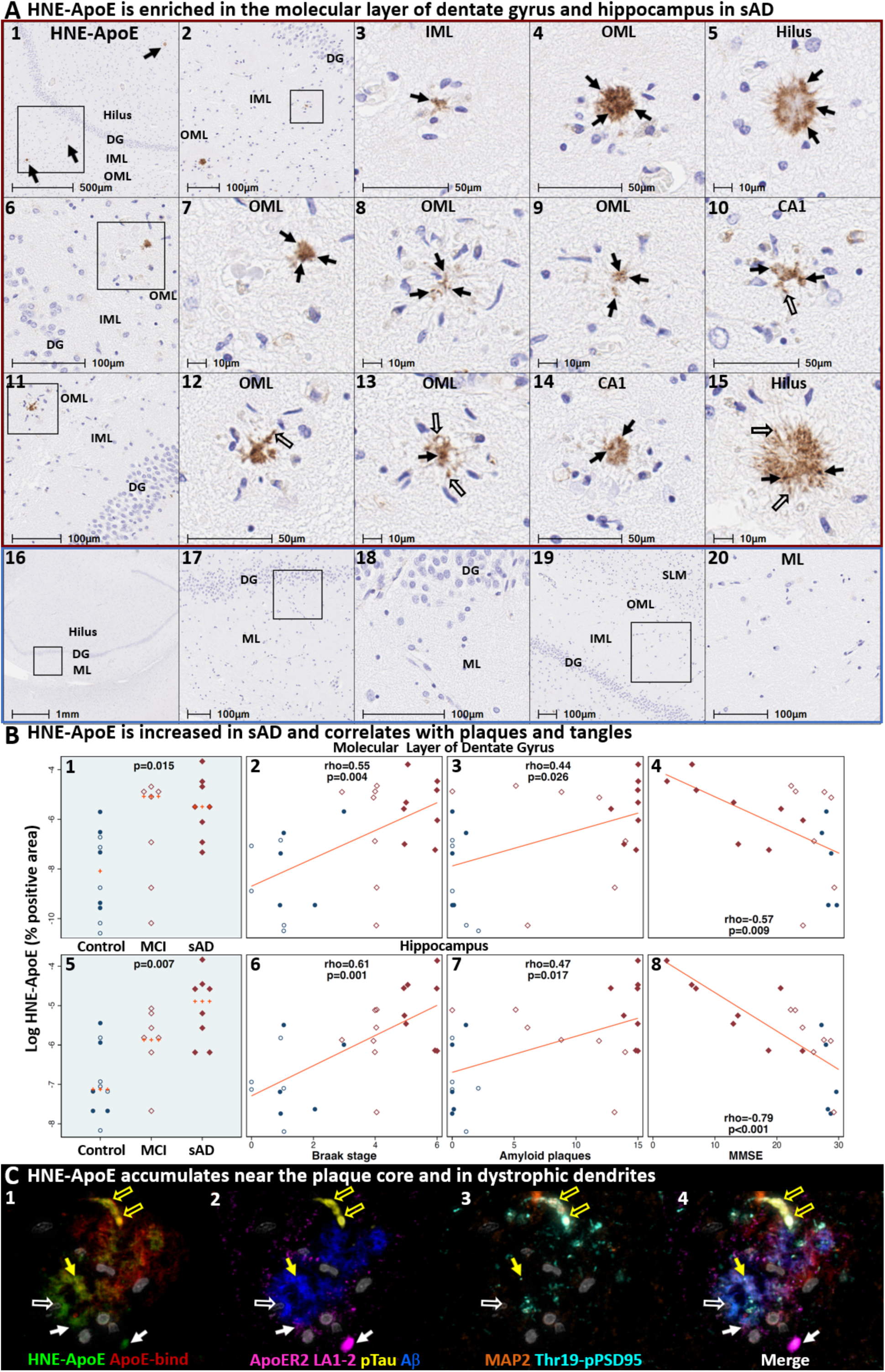
HNE-ApoE accumulates in the terminal zones of perforant path projections in sporadic AD. Coronal sections of the hippocampal formation from three (Braak stage V-VI, APOE3/3) sAD cases (**A_1-15_**) and non-AD controls (**A_16-20_**) were stained with an anti-HNE-ApoE antibody. HNE-ApoE immunoreactive plaques were observed in the outer molecular layer (OML) of the dentate gyrus (**A_1-4_**, **A_6-9_**, **A_11-13_**), inner molecular layer (IML) of dentate gyrus (**A_1,3_**) CA1 region (**A_14_**) and the hilus of the hippocampus (**A_5_**, **A_15_**) in AD cases, but not in controls (**A_16-20_**). HNE-ApoE immunolabeled granular structures near the plaque core (arrows in **A**) and elongated structures projecting outside of the plaque core (open arrows in **A**). HNE-ApoE accumulated in sAD cases and correlated with histochemical progression and cognitive deficits in both the molecular layer of dentate gyrus and hippocampus (**B**). MP-IHC labeling (**C_1-4_**) revealed partial, but incomplete overlap between HNE-ApoE and native ApoE and Aβ immunolabeling (yellow arrows in **C_1-2_**). Open yellow arrows in **C_1-4_** depict elongated HNE-, Ser202/Thr205-pTau and MAP2-positive structures that are consistent with dystrophic neurites. Closed white arrows depict close spatial relationships between HNE-ApoE and ApoER2 LA1-2 expression. Gray objects in **C_1-4_** represent nuclei stained with DNA dye (DAPI). Red and blue boxed panels in **A_1-15_** indicate and **A_16-20_** indicate sAD cases and non-AD controls, respectively. **Panel B**: open and closed blue circles indicate young controls and age-matched controls; open and closed red diamonds indicate MCI cases and sAD cases, respectively. **Abbreviations**: DG, dentate granule cells; ML, molecular layer; OML, outer molecular layer; IML, inner molecular layer; SLM, stratum lacunosum-moleculare; CA, cornu ammonis; NEUN, neuronal nuclear/soma antigen; MAP2, microtubule associated protein-2; pTau, Ser202/Thr205-pTau.

### ApoE accumulates in the terminal zones of perforant path projections in sAD

IHC and MP-IHC staining confirmed that antibodies targeting several domains in ApoE strongly label plaques in the molecular layer of dentate gyrus and hippocampus in sAD (**Fig 6A,C**). ApoE expression in both annotated regions was increased in sAD cases and positively correlated with Braak stage and Aβ plaque load (but not MMSE score) (**Fig 6B**). Using single-marker IHC in serial sections, we observed prominent ApoE expression in plaques specifically within the perforant path terminal zones including the molecular layer of the dentate gyrus (**Fig 6A_4_**) and the SLM subfield of CA1 (**Extended Fig 6.1K**). Single-marker IHC also revealed regional colocalization of ApoE-enriched plaques and ApoER2 LA1-2 aggregates within perforant path terminal zones (**Extended Fig 6.1**). MP-IHC confirmed that multiple domains of ApoE are enriched in the central core of many neuritic plaques (**Fig 6C**), and revealed close spatial relationships between ApoE, lipid-laden microglia and reactive astrocytes in the neuritic plaque niche (**Extended Fig 6.2**).

### Lipid peroxidation modified ApoE accumulates in entorhinal-hippocampal structures in sAD

As depicted in **Fig 3**, we hypothesized that lipid peroxidation-induced adduction and crosslinking of ApoE particles and ApoE receptors could disrupt ApoE receptor-mediated delivery of lipids to neurons, resulting in extracellular trapping and deposition of peroxidized ApoE particles. Single-marker IHC revealed that the anti-HNE-ApoE antibody labels discrete granular structures near the plaque core (black arrows in **Fig 7A**), elongated structures projecting outside of the plaque core (open arrows in **Fig 7A** and **Extended Fig 7A**), and unspecified peri-vascular components (**Extended Fig 7.1.A_14-15_**) in regions affected by sAD (**Fig 7A_1-15_**), with little or no labeling in non-AD controls (**Fig 7A_16-20_**). HNE-ApoE expression was higher in sAD compared to non-AD controls and positively correlated with Braak stage and Aβ plaque load (**Fig 7B**). HNE-ApoE expression in dentate gyrus and hippocampus (but not temporal cortex) also negatively correlated with MMSE (**Fig 7B** and **Extended Fig 7.1**). MP-IHC revealed partial, but incomplete overlap between HNE-ApoE and native ApoE immunolabeling (**Fig 7C_1_**), suggesting that a portion of ApoE in the plaque environment is modified by lipid peroxidation. Open yellow arrows and closed white arrows in **Fig 7C_1-4_** depict HNE-, Ser202/Thr205-pTau and MAP2-positive structures that are consistent with dystrophic neurites, and close spatial relationships between HNE-ApoE and ApoER2 LA1-2 expression, respectively. Additional IHC and MP-IHC examples of HNE-ApoE expression in perforant path terminal zones and temporal cortex are provided in **Extended Figs 7.1-7.2 and Figs 12-13**.

### Plaque associated Reelin accumulation in the terminal zones of perforant path projections

According to our model (**Fig 3**), adduction and crosslinking of ApoE and ApoER2 disrupts Reelin-ApoER2 binding, thereby trapping Reelin in the extracellular space. Reelin has previously been reported to accumulate in the vicinity of Aβ-containing plaques in rodent and primate models of aging,[86, 87] and in the human frontal cortex [88] and stratum oriens of the hippocampus in AD.[89] To probe for evidence of Reelin aggregates in the human perforant path target zones, we immunolabeled Reelin in the hippocampal formation of 24 cases (**Fig 8**). Representative IHC and MP-IHC images from two Braak stage V-VI sAD cases reveal prominent plaque-associated aggregates of Reelin (**Fig 8A**) that were most prominent in the SP subfield of CA1-2 and the molecular layer dentate gyrus. Plaque-associated Reelin aggregates in the molecular layer of the dentate gyrus were more abundant in sAD cases than in non-AD cases and negatively correlated with MMSE scores; Reelin aggregates in the dentate gyrus and hippocampus correlated with histological progression (**Fig 8B**). MP-IHC revealed intimate spatial and morphological relationships between Reelin aggregates, Aβ, Ser202/Thr205-pTau, MAP2 and Thr19-pPSD95 in the vicinity of neuritic plaques (**Fig 8C**). Additional IHC examples of Reelin aggregates in the perforant path terminal zones in a sAD case with early sAD (1 year history of dementia, MMSE 24) are provided in **Extended Fig 12**. Spatial and morphological relationships of Reelin with other ApoE-ApoER2-Dab1 pathway components for the same early sAD case are provided in **Fig 12**.

**Fig 8.**
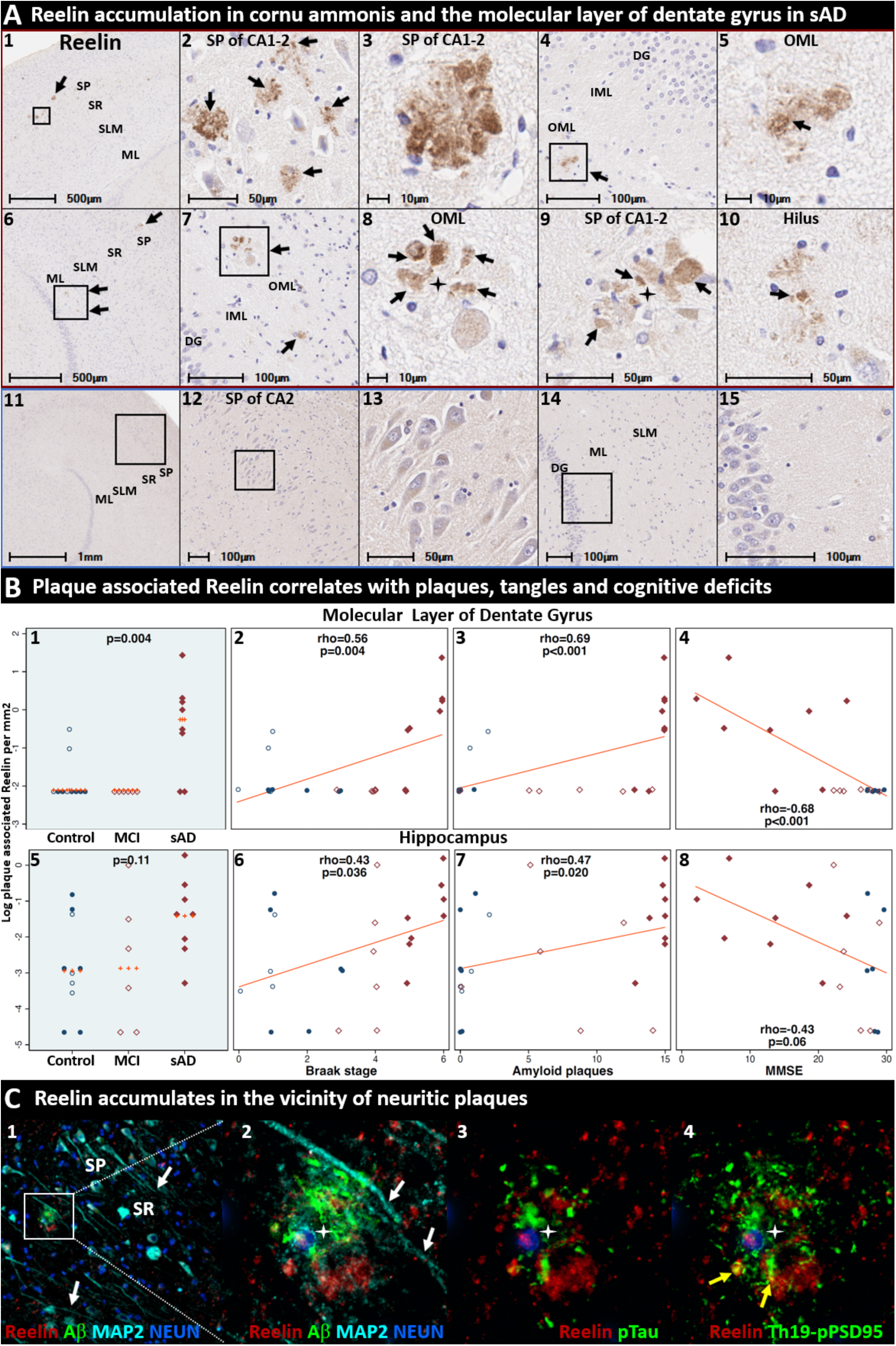
Reelin accumulates in the terminal zones of perforant path projections in sporadic AD. Coronal sections of the molecular layer of the dentate gyrus and hippocampus from two representative (Braak stage V-VI, APOE3/3) sAD cases (**A_1-10_**) and a non-AD control case (**A_11-15_**) were stained with an antibody targeting a C-terminal domain of Reelin. Reelin aggregates (black arrows) were observed in the CA2 subregion of the hippocampus (**A_1-3_, A_9_**), outer molecular layer of the dentate gyrus (**A_4-8_**) and hilus of the hippocampus (**A_10_**) in sAD cases, but not in controls. Plaque-associated Reelin aggregates in the molecular layer of the dentate gyrus were more abundant in sAD cases than in controls and negatively correlated with MMSE scores; Reelin aggregates in the hippocampus correlated with histological progression (**B_1-8_**). MP-IHC labeling in the CA2 subregion (**C_1-4_**) revealed Reelin aggregates are in close proximity to Aβ, Se202/Thr205-pTau, and Thr19-pPSD95. White arrows in **C_1-2_** depict MAP2-positive dendritic processes emanating toward the SR sublayer of CA2. Yellow arrows in **C_4_** depict close spatial relationships and partial co-localization of Reelin with Thr19-pPSD95. Red and blue boxed panels in **A_1-10_** and **A_11-15_** indicate sAD cases and non-AD controls, respectively. **Panel B**: open and closed blue circles indicate young controls and age-matched controls; open and closed red diamonds indicate MCI cases and sAD cases, respectively. **Abbreviations**: DG, dentate granule cells; ML, molecular layer; SP, stratum pyramidale; SR, stratum radiatum; SLM, stratum lacunosum-moleculare; CA, cornu ammonis.

### Plaque associated Dab1 accumulation in the terminal zones of perforant path projections

According to our model (**Fig 3**), adduction and crosslinking of ApoE and ApoER2 disrupts Reelin-ApoER2-Dab1 signaling cascades in neurons affected by sAD. Since Reelin signaling through ApoER2 induces Dab1 (and ApoER2) degradation,[90] impaired binding of Reelin to ApoER2 is expected to lead to accumulation of Dab1 and ApoER2 proteins. Representative IHC and MP-IHC images from a Braak stage V-VI sAD cases reveal prominent plaque-associated aggregates of Dab1 that were most prominent in CA2 and CA1 subregions of the hippocampus and the molecular layer dentate gyrus (**Fig 9A** **1-10, Extended Fig 9**). Large Dab1 complexes (diameters of up to 50-75µm) were observed in CA2 in some sAD cases (see **Fig 9A_8_**, **Extended Fig 9.1**). By contrast, in non-AD controls Dab1 expression was localized to neuronal somata and projections with no plaque associated aggregates observed (**Fig 9A_11-15_**). Plaque-associated Dab1 aggregates in the molecular layer of the dentate gyrus and hippocampus were more abundant in AD cases than in non-AD cases, positively correlated with histological progression, and negatively correlated with MMSE scores. (**Fig 9B**). IHC with serial sections revealed regional colocalization of Dab1 and ApoER2 LA1-2 (**Fig 9C_1-4_**).

**Fig 9.**
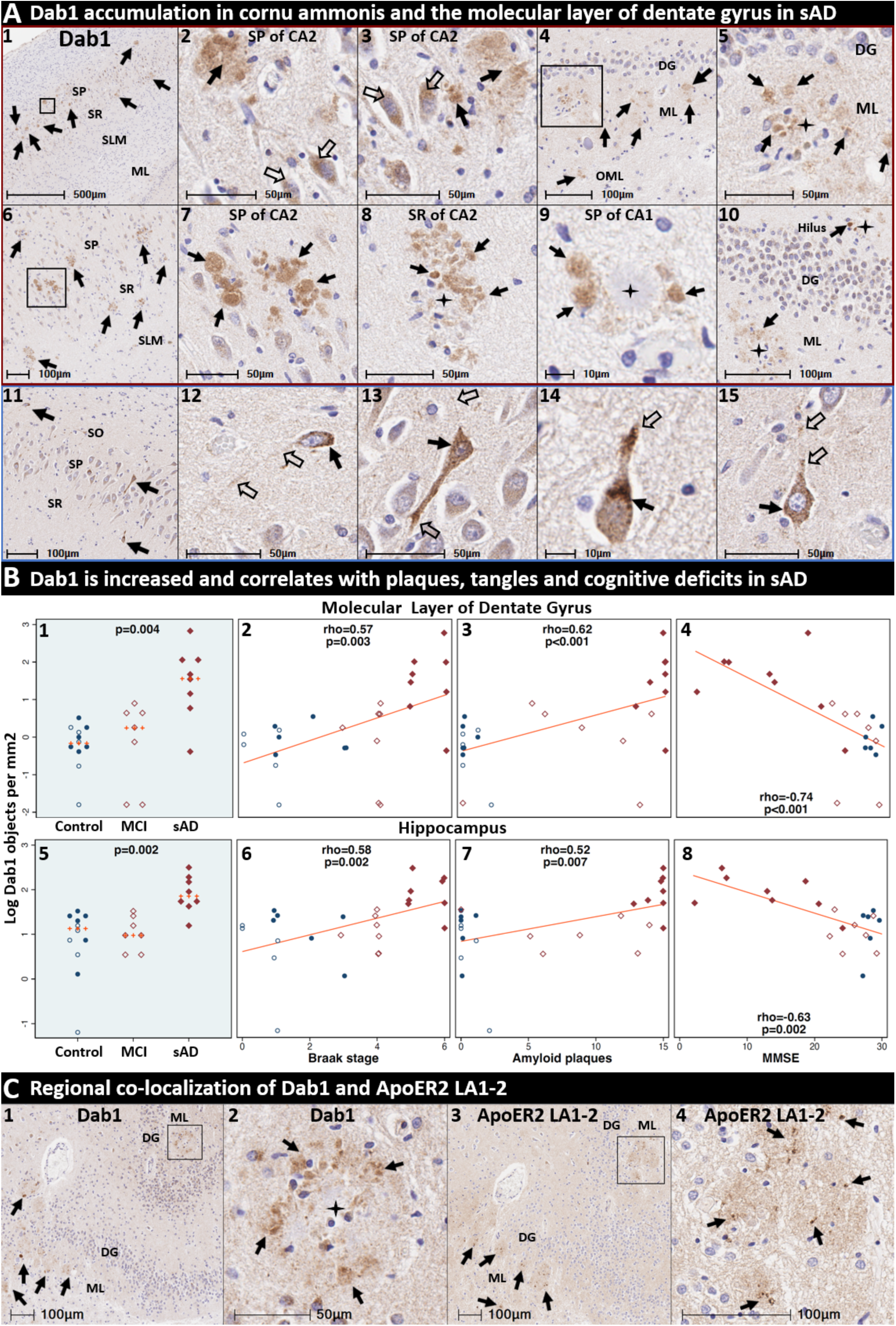
Dab1 accumulates in the terminal zones of perforant path projections in sporadic AD. Coronal sections of the molecular layer of the dentate gyrus and hippocampus from two representative (Braak stage V-VI, APOE3/3) sAD cases (**A_1-10_**) and a non-AD control case (**A_11-15_**) were stained with an anti-Dab1 antibody. In sAD cases, plaque-associated Dab1 aggregates were observed in the SP and SR of the CA2 (**A_1-3, 6-8_**) and CA1 (**A_9_**) subregions of the hippocampus, the molecular layer of the dentate gyrus (A**_4-5, 10_**), and the hilus of the hippocampus (**A_10_**) in sAD cases. Arrows in **A_1-10_** shows Dab1 expression in both plaques (black arrows) and neurons (open arrows). By contrast, in non-AD controls Dab1 expression is localized to somata (black arrows) and projections (open arrows) of a subset of neurons within the SO (**A_11-12_**), SP (**A_11_**, **A_13_**) and SR (**_A11_**, **A_14-15_**) subfields of CA1 and CA2 with no plaque associated aggregates observed. Dab1 aggregates in both the molecular layer of dentate gyrus and hippocampus were more abundant in sAD cases compared to controls, positively correlated with histochemical progression and negatively correlated with antemortem MMSE scores (**B_1-8_**). IHC with serial sections revealed regional colocalization of Dab1(**C_1-2_**) and ApoER2 LA1-2 (**C_3-4_**). Red and blue boxed panels in **A_1-10_** and **A_11-15_** indicate sAD cases and non-AD controls, respectively. **Panel B**: open and closed blue circles indicate young controls and age-matched controls; open and closed red diamonds indicate MCI cases and sAD cases, respectively. **Abbreviations**: DG, dentate granule cells; ML, molecular layer; SP, stratum pyramidale; SR, stratum radiatum; SLM, stratum lacunosum-moleculare; CA, cornu ammonis.

These observations, which imply failure of ApoER2 to convey the Reelin signal across the membrane leading to accumulation of all three markers (Reelin, ApoER2, Dab1), provide additional evidence for the proposed pathogenic nexus centered around ApoER2 disruption. Additional examples of plaque associated Dab1 aggregates in subfields of the hippocampus are provided in **Fig 12** and **Extended Figs 9-10** and **13**.

**Fig 10.**
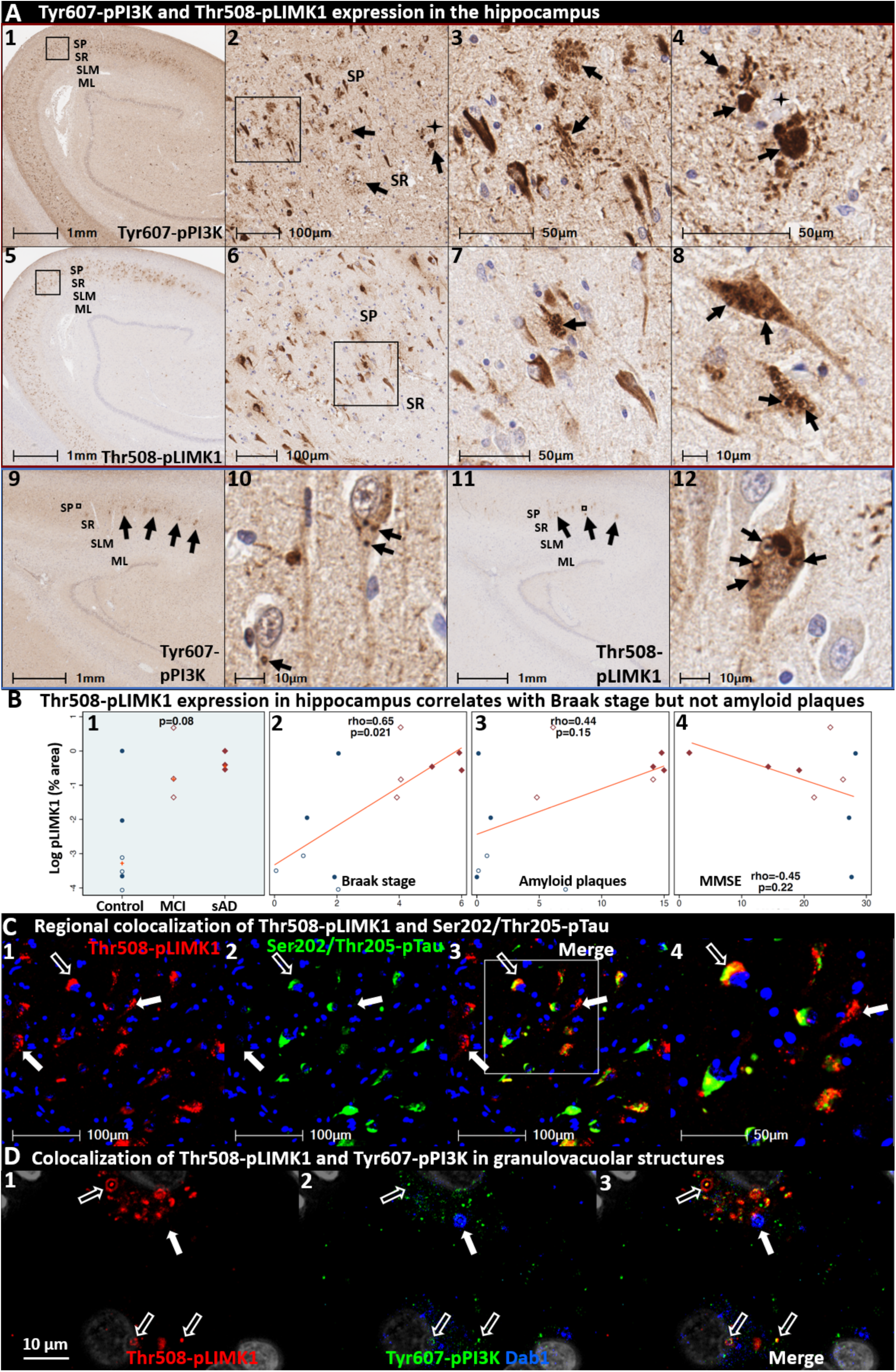
Phosphorylated PI3K and LIMK1 accumulate in the cornu ammonis in sporadic AD. Coronal sections through the hippocampus from a representative (Braak stage V, APOE3/3) sAD case (**A_1-8_**) and a non-AD control case (**A_9-12_**) were stained with antibodies targeting Tyr607-pPI3K (**A_1-4, 9-10_**) and Thr508-pLIMK1 (**A_5-8, 11-12_**). Tyr607-pPI3K was abundant in neuritic plaques (arrows in **A_2-4_**) in the CA1-2 subregions of the hippocampus and the molecular layer of the dentate gyrus (see Fig 12) in sAD cases, which were rare or absent in non-AD controls (**A_9-10_**). Tyr607-pPI3K and Thr508-pLIMK1 were both abundant in a subset of abnormal neurons (arrows in **A_1-4_** and **A_5-8_**, respectively) in sAD cases. Thr508-pLIMK1 was most prominent within granulovacuolar structures in CA1-2 pyramidal neurons that were observed in both sAD cases (arrows in **A_7-8_**) and non-AD controls (arrows in **A_11-12_**). Thr508-pLIMK1 expression correlated with Braak stage (**B_2_**) but not Aβ plaque load or MMSE (**B_3-4_**). MP-IHC (**C_1-4_**) revealed that despite strong regional colocalization, Thr508-pLIMK1 and Ser202/Thr205-pTau have only partial cellular colocalization. Open arrows in **C_1-4_** depict cellular colocalization of both markers; closed arrows identify neurons that strongly express Thr508-pLIMK1 with minimal or no Ser202/Thr205-pTau expression. MP-IHC in CA2 pyramidal neurons (**D_1-3_**) revealed that Thr508-pLIMK1 and Tyr607-pPI3K partially co-localize within granulovacuolar structures (open white arrows), however neither marker colocalized with Dab1 (closed arrows). Gray objects in **D_1-3_** represent nuclei stained with DNA dye (DAPI). Red and blue boxed panels in **A_1-8_** and **A_9-12_** indicate sAD cases and non-AD controls, respectively. **Panel B**: open and closed blue circles indicate young controls and age-matched controls; open and closed red diamonds indicate MCI cases and sAD cases, respectively. **Abbreviations**: DG, dentate granule cells; ML, molecular layer; SP, stratum pyramidale; SR, stratum radiatum; SLM, stratum lacunosum-moleculare.

### Tyr607-pPI3K and Thr508-pLIMK1 accumulate in the vicinity of neuritic plaques in the entorhinal-hippocampal memory system

As depicted in **Fig 3**, ApoER2 disruption is proposed to impair downstream Reelin-ApoER2-Dab1 signaling cascades involving the phosphorylation of PI3K, LIMK1 and Tau. To examine this concept in the human entorhinal-hippocampal memory system, we labeled the core downstream Reelin signaling partners Tyr607-pPI3K and Thr508-pLIMK1 in 12 cases spanning the clinicopathological spectrum of sAD. Hippocampus images from representative Braak stage V sAD case and non-AD control are provided in **Fig 10** and **Extended Figs 10 and 12**. In sAD cases, Tyr607-pPI3K and Thr508-pLIMK1 were observed to accumulate in the immediate vicinity of neuritic plaques in CA1-2 and the molecular layer of the dentate gyrus, in a similar regional distribution as observed for Reelin, ApoER2 and Dab1 accumulation (**Fig 10, Extended Figs 10** and **12**). Tyr607-pPI3K was ubiquitously expressed by neurons in CA1-3 regions in both sAD cases and controls, with particularly strong expression observed in abnormal neurons surrounding neuritic plaques in sAD cases. Thr508-pLIMK1 expression was present in a smaller subset of neurons, with the most prominent expression observed in granulovacuolar structures in CA1-2 neurons that are consistent with granulovacuolar degeneration bodies (**Fig 10A**). Thr508-pLIMK1 expression in hippocampus positively correlated with Braak score but not total Aβ plaque load (**Fig 10B**).

Since LIMK1 and Tau are downstream components of the Reelin-ApoER2-Dab1-PI3K signaling cascades that stabilize actin (via Thr508-pLIMK1) [91, 92] and microtubules (via Tau),[93] respectively (**Fig 2**), we next used MP-IHC to gain insight into the spatial, morphological and cytoarchitectural relationships between Thr508-pLIMK1 and Ser202/Thr205-pTau. Representative MP-IHC images from the CA2 region of a Braak stage V sAD case reveal strong regional co-expression of both markers localized to neuritic plaques and subsets of surrounding neurons (**Fig 10C**); however only partial cellular co-localization of Thr508-pLIMK1 and Ser202/Thr205-pTau was observed. MP-IHC revealed colocalization of Tyr607-pPI3K and Thr508-pLIMK1 (but not Dab1) within a subset of these granulovacuolar structures (**Fig 10D**). Additional IHC examples of Tyr607-pPI3K and Thr508-pLIMK1 expression in hippocampus and molecular layer of the dentate gyrus are provided in **Extended Fig 10** and in **Fig 12**, respectively.

### Thr19-pPSD95 accumulates within and around ApoE-enriched neuritic plaques in sAD

Postsynaptic ApoER2-PSD95-NMDA receptor complexes play a pivotal role of in LTP and memory in experimental models (**Fig 2**).[42, 43, 94–97] Disruption of Reelin-ApoER2-Dab1-PI3K signaling activates GSK-3β, which in turn phosphorylates Tau. Activated GSK3β also phosphorylates PSD95 at Thr19 to induce disassembly of postsynaptic receptor complexes,[72] suggesting that PSD95 and Tau phosphorylation may both be regulated by the Reelin-ApoER2 axis. Synapse disassembly and loss are major neurobiological correlates of memory loss, and Reelin deficiency has been shown to decrease PSD95 at the post-synapse,[34] however to our knowledge Thr19-pPSD95 has not previously been investigated in AD. To address this gap, we labeled Thr19-pPSD95 in hippocampus and temporal cortex specimens from cases spanning the clinicopathological spectrum of sAD. Images from these regions in a representative Braak stage VI sAD case and non-AD control are provided in **Fig 11A**. In sAD cases, we observed striking accumulation of Thr19-pPSD95 in the vicinity of neuritic plaques and within surrounding abnormal neurons in the molecular layer of dentate gyrus, CA1-2 subfields of the hippocampus, and the temporal cortex (**Fig 11A** and **Fig 12**). Non-AD controls had little or no expression of Thr19-pPSD95. MCI cases exhibited more modest expression of Thr19-pPSD95 that was mostly localized to abnormal neurons. Thr19-pPSD95 expression in all three annotated regions was strongly, positively correlated with Braak stage and Aβ plaque load, and negatively correlated with MMSE scores (**Fig 11B** and **Fig 12A**). MP-IHC revealed that Thr19-pPSD95 is primarily localized to MAP2-positive dendritic compartments (**Fig 11C**). Unlike Aβ, Thr19-pPSD95 and ApoE are abundant in neuritic plaques but not diffuse plaques (**Extended Fig 11.1**). Additional IHC and MP-IHC examples demonstrating plaque-associated accumulation of Thr19-pPSD95, and spatial relations with other ApoE/Reelin-ApoER2-Dab1 pathway components are provided in **Figs 7-8 & 12-13**.

**Fig 11.**
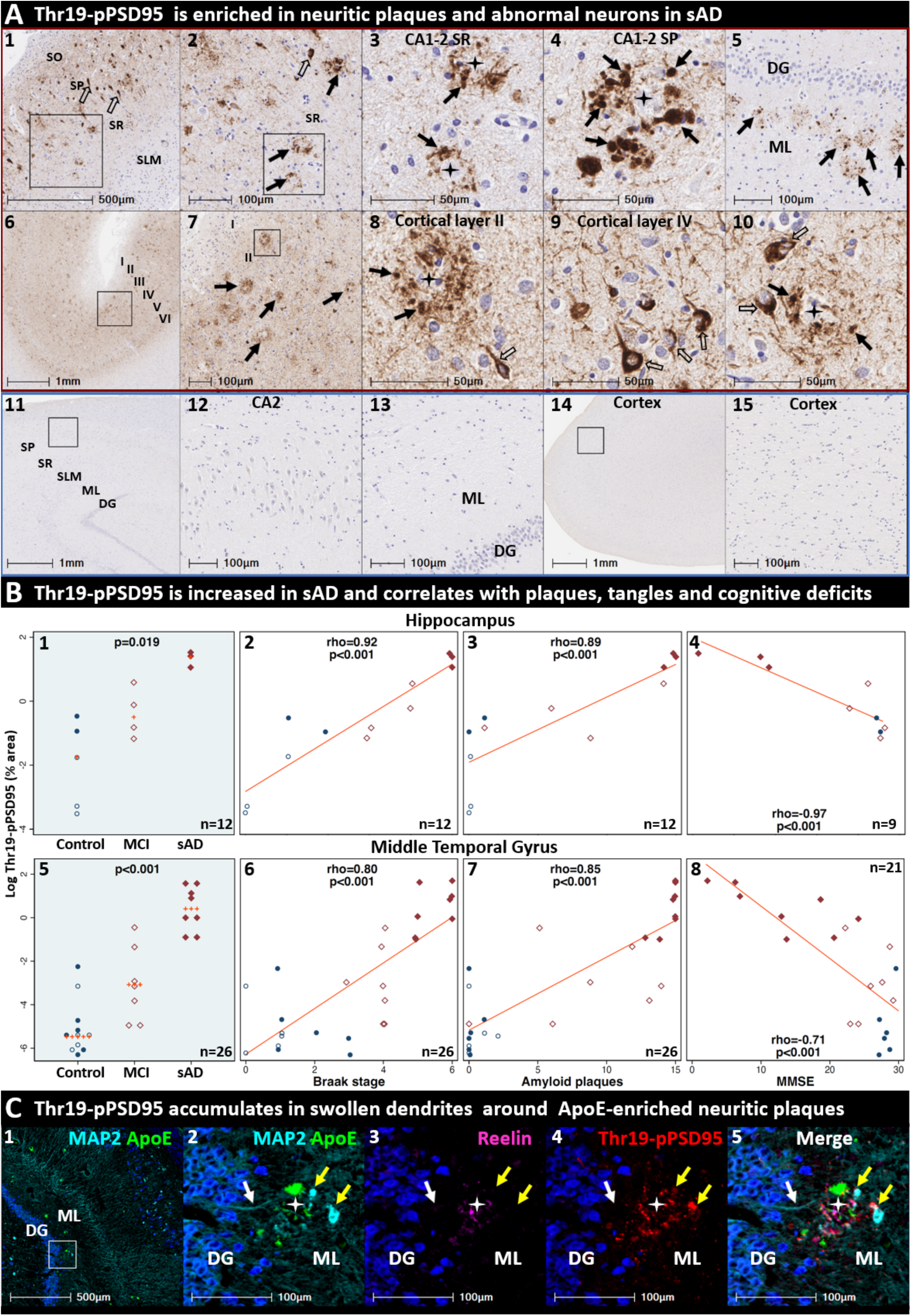
Thr19-pPSD95 accumulation as evidence for synapse disassembly in sporadic AD. Coronal sections through the hippocampus and middle temporal gyrus from a representative (Braak stage VI, APOE3/3) sAD case (**A_1-10_**) and a non-AD control case (**A_11-15_**) were stained with antibodies against Thr19-pPSD95, a marker of post-synapse disassembly that is regulated by GSK3β. Thr19-pPSD95 was abundant in prominent globular structures (black arrows) and smaller fibrillar structures (open arrows) in the vicinity of neuritic plaques and surrounding abnormal neurons in the cornu ammonis (**A_1-4_**), molecular layer of the dentate gyrus (**A_5_**) and middle temporal gyrus (**A_6-10_**) in AD cases with minimal or no expression in controls. Thr19-pPSD95 expression strongly correlated with histochemical progression and cognitive deficits (**B_2-4, 6-8_**). MP-IHC revealed complex spatial and morphological relationships between Thr19-pPSD95 and other ApoE/Reelin-ApoER2 pathway components in the neuritic plaque niche (**C_1-5_**), and substantial co-localization of Thr19-pPSD95 with MAP2 in swollen, dystrophic dendrites (yellow arrows in **C_2_**, **C_4-5_**). White arrows in **C_2-5_** show a MAP2-positive dendritic projection emanating from the DG into the ML. Red and blue boxed panels in **A_1-10_** and **A_11-15_** indicate sAD cases and non-AD controls, respectively. **Panel B**: open and closed blue circles indicate young controls and age-matched controls; open and closed red diamonds indicate MCI cases and sAD cases, respectively. **Abbreviations**: DG, dentate granule cells; ML, molecular layer; SP, stratum pyramidale; SR, stratum radiatum.

**Fig 12.**
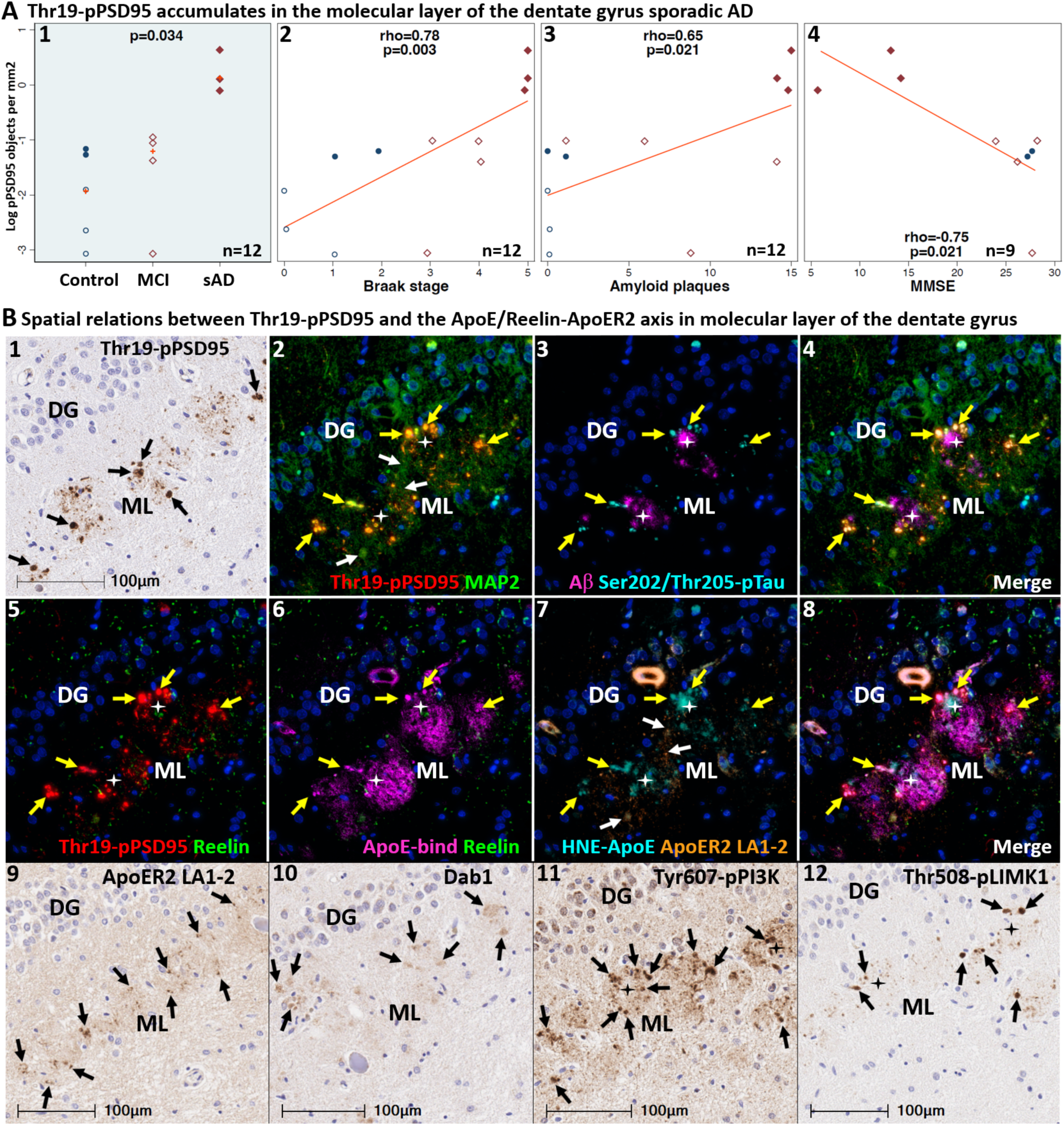
Convergence of ApoE/Aβ/Reelin-ApoER2-Dab1-PI3K-PLIMK1-Tau-PSD95 pathologies in the molecular layer of the dentate gyrus in sAD. Thr19-pPSD95 expression in the molecular layer of the dentate gyrus was increased in sAD cases, positively correlated with Braak stage and Aβ plaque load, and negatively correlated with MMSE scores (**A_1-4_**). Single marker IHC (**B_1_**) and MP-IHC (**B_2-12_**) were used to label serial coronal sections of the dentate gyrus from the same sporadic AD case (Braak stage VI, APOE3/3) shown in **Fig 11A_1-5_**. Single marker IHC revealed prominent Thr19-pPSD95-positive globular structures (open arrows in **B_1_**) amid numerous smaller fibrillar structures and discrete puncta in this region. MP-IHC revealed that these globular structures co-express Thr19-pPSD95 and MAP2 (arrows in **B_2_**), suggesting localization to dystrophic dendrites. Colocalization of Ser202/Thr205-pTau (arrows in **B_3-4_**) within a subset of these Thr19-pPSD95 and MAP2-positive globular structures is consistent with somatodendritic localization of Tau. Expression of ApoE, HNE-ApoE, and Reelin was primarily localized to extracellular plaques (depicted by stars in **B_5-8_**). HNE-ApoE exhibited partial, incomplete colocalization with native ApoE in the molecular layer of the dentate gyrus (**B_6-8_**). Panels **B_5-8_** reveal partial co-localization of HNE-ApoE and Thr19-pPSD95 within swollen dendrites (yellow arrows) that surround BAP and ApoE enriched plaques (white stars). Multiplex and single-marker IHC revealed that ApoER2 LA1-2 expression in this region was localized to fibrillar structures and discrete puncta (**B_7-9_**) in the immediate vicinity of neuritic plaques, while single-marker IHC revealed localized accumulation of Dab1, and strong expression of Tyr607-pPI3K and Thr508-pLIMK1 in unspecified globular structures that resemble dystrophic neurites. **Panel A**: open and closed blue circles indicate young controls and age-matched controls; open and closed red diamonds indicate MCI cases and sAD cases, respectively. **Abbreviations**: DG, dentate granule cells; ML, molecular layer.

**Fig 13.**
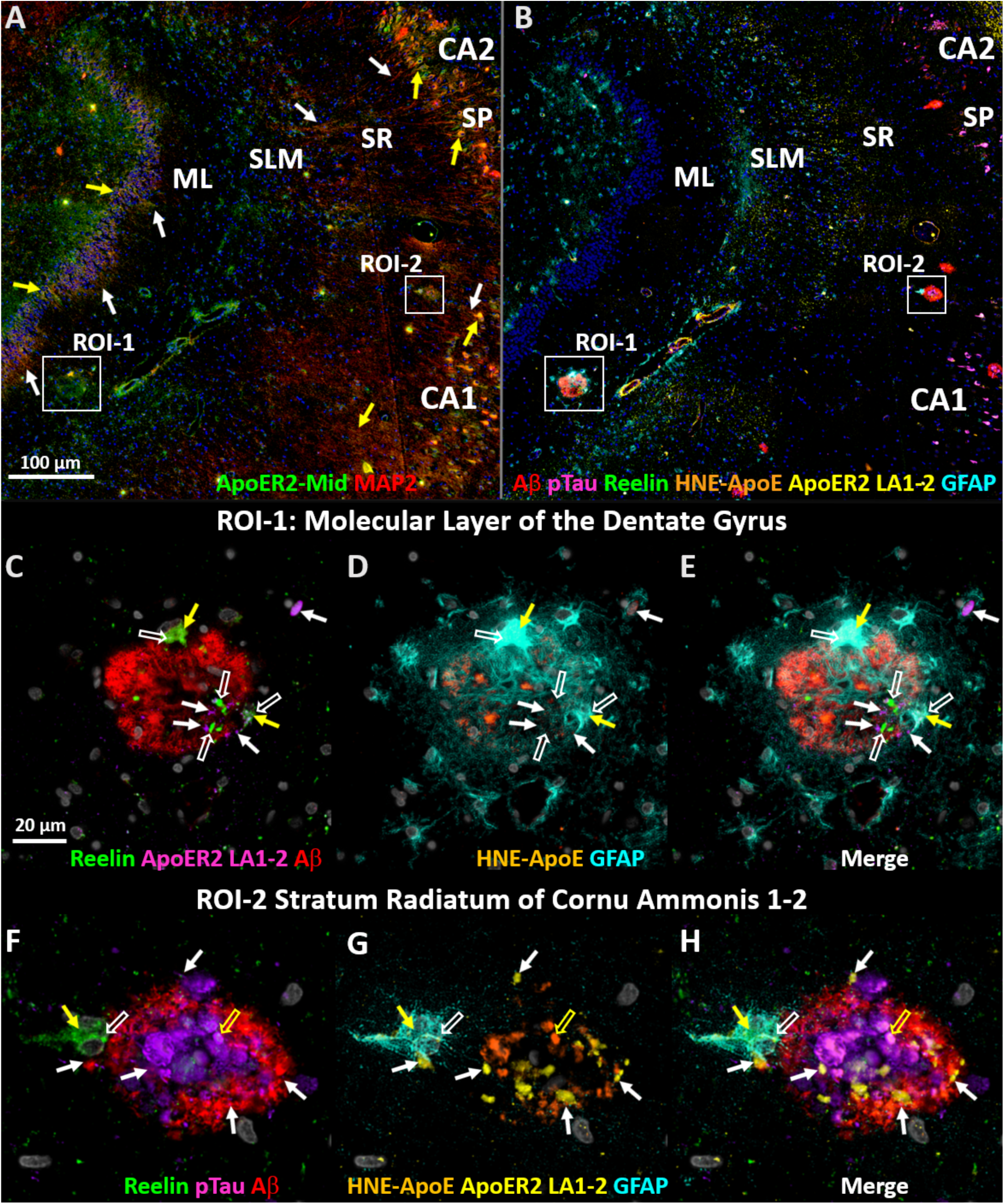
Evidence for ApoE/Reelin-ApoER2 axis pathologies in perforant path target zones in early sAD. MP-IHC was used to label coronal sections of the perforant path target zone from a (Braak stage V, APOE3/3) sAD case with a 1-year history of dementia (MMSE 24) (**A-H**). Panel **A** depicts colocalization (yellow arrows) of ApoER2-YWTD and MAP2 within a subset of dentate granule cells and CA2 and CA1 pyramidal neurons whose dendritic arbors (white arrows) emanate into the ML and SR/SLM subfields, respectively. Panel **B** depicts hallmark AD pathologies and several Reelin-ApoER2 axis pathologies within two regions of interest (ROI), the molecular layer of the dentate gyrus (ROI-1) and the SR subfield of CA1/2 (ROI-2). Panels **C-E** reveal accumulation of Reelin (open white arrows), ApoER2 LA1-2 (white arrows), and HNE-ApoE (open yellow arrows) in the immediate vicinity of a single neuritic plaque in the molecular layer of the dentate gyrus. Panels **F-H** reveal discrete, punctate accumulations of ApoER2 LA1-2 (closed white arrows), and Reelin (open white arrows) in proximity to a Ser202/Thr205-pTau-, HNE-ApoE- (open yellow arrows), and Aβ-positive neuritic plaque in the SR subfield of CA1-2. Yellow arrows in **C-H** depict co-localization of Reelin with plaque-associated astrocytes, suggesting potential phagocytosis, in both regions. **Abbreviations**: DG, dentate granule cell layer; ML, molecular layer; CA, cornu ammonis.

### Convergence of ApoE/Reelin-ApoER2-Dab1 axis pathologies within dense dendritic arbors of the molecular layer of the dentate gyrus

In **Figs 5-11**, we presented evidence for enrichment of ApoE, Reelin, ApoER2 LA1-2, Dab1, Tyr607-pPI3K, Thr508-pLIMK1 and Thr19-pPSD95 in the molecular layer of the dentate gyrus and the SR and SLM of CA1-2 subfields of the hippocampus, suggesting convergence of ApoE/Reelin-ApoER2 axis pathologies near the synaptic recipients of the perforant path axonal projections.[12, 15, 85] We next used MP-IHC to examine spatial, morphological, and cytoarchitectural relationships between individual ApoE/Reelin-ApoER2-Dab1 pathway markers in the neuritic plaque niche. An enlarged image of the molecular layer of the dentate gyrus from an advanced (Braak stage VI, ApoE3/3) sAD case containing extensive neuritic lesions is provided in **Fig 12B_1-12_**. Single marker IHC revealed prominent Thr19-pPSD95-positive globular structures (black arrows in **Fig 12B_1_**) amid numerous smaller fibrillar structures in this region. MP-IHC revealed that these globular structures co-express Thr19-pPSD95 and MAP2 (yellow arrows in **Fig 12B_2_**), suggesting localization of Thr19-pPSD95 to swollen, dystrophic dendrites. Colocalization of Ser202/Thr205-pTau (yellow arrows in **Fig 12B_3-4_**) within a subset of these Thr19-pPSD95 and MAP2-positive globular structures is consistent with somatodendritic localization of Tau. By contrast, expression of ApoE, HNE-ApoE, and Reelin was primarily localized to extracellular plaques (depicted by stars in **Fig 12B_5-8_**), consistent with the concept of localized ApoE receptor disruption. As previously observed in cornu ammonis (**Fig 7C**), HNE-ApoE exhibited only partial colocalization with native ApoE in the molecular layer of the dentate gyrus (**Fig 12B_6-8_**). Single-marker IHC revealed that ApoER2 LA1-2 expression in this region was localized to fibrillar structures and discrete puncta (black arrows in **Fig 12_B-9_**). White arrows in **Fig 12B_2_** and **B7** indicate that these ApoER2 LA1-2-positive puncta reside within dense, MAP2-positive dendritic arbors in the immediate vicinity of neuritic plaques. Single-marker IHC revealed regional accumulation of Dab1, and strong expression of Tyr607-pPI3K and Thr508-pLIMK1 in unspecified globular structures that resemble dystrophic neurites (**Fig 12B_10-12_**). These collective observations suggest that dendritic compartments and postsynaptic densities within the terminal zones of the perforant path are a major locus for ApoE/Reelin-ApoER2-Dab1-PI3K-LIMK1-Tau axis pathologies in sAD, with Thr19-pPSD95 accumulation suggesting that these axis pathologies culminate in synaptic disassembly.

### ApoE/Reelin-ApoER2-Dab1 axis pathologies are evident in the perforant path target zones of in early sAD

We next sought to determine if there was evidence for ApoE/Reelin-ApoER2 axis disruption in perforant path terminal zones in early sAD by using multiplex- and single-marker IHC in a case with a 1-year history of dementia and only moderate cognitive impairment (MMSE 24). We placed a particular emphasis on two regions of interest with dense perforant pathway projections in humans (1) the molecular layer of the dentate gyrus; and (2) the SR subfield of CA2, which has recently been shown to receive a large perforant path projection in humans [16] but not rodents. In this early sAD case, we observed that ApoER2-YWTD and MAP2 are co-expressed in somata of the dentate granule cells and CA1-2 pyramidal neurons (yellow arrows in **Fig 13A**), whose apical dendrites emanate into the molecular layer of the dentate gyrus and SR and SLM subfields (white arrows in **Fig 13A**). We observed localized accumulations of Reelin (open white arrows in **Fig 13C-H**), ApoER2 LA1-2 (closed white arrows in **Fig 13C-H**), and HNE-ApoE (open yellow arrows in **Fig 13C-H**) in the immediate vicinity of Aβ and Ser202/Thr205-pTau-positive neuritic plaques in both regions, suggesting Reelin-ApoER2-Dab1 axis disruption. Closed yellow arrows in **C-H** depict co-localization of Reelin with plaque-associated astrocytes in both regions, suggesting activation of glial-clearance pathways. Single-marker IHC images using serial sections from the same case (**Extended Fig 13A**) provide additional evidence for localized ApoER2-YWTD expression, and Reelin-ApoER2-Dab1 pathway disruption in the perforant path terminal zones in this early sAD case. Particularly strong Thr19-pPSD95 expression was observed within pyramidal neurons, their dense dendritic arbors, and neuritic plaques in SR subfields of CA2, suggesting widespread synaptic disassembly in this region. Collective observations demonstrate derangements in multiple ApoE/Reelin-ApoER2-Dab1-PSD95 pathway components in the perforant path terminal zones in this early sAD case, and suggest that these pathologies are not a late-stage phenomenon in AD.

### ApoE/Reelin-ApoER2-Dab1 axis pathologies intersect with hallmark AD pathologies and glial-clearance pathways in the vicinity of neuritic plaques

MP-IHC revealed close spatial relations between ApoE/Reelin-ApoER2-Dab1 axis markers and four hallmark AD pathologies—Aβ-deposits, Tau tangles, microglial infiltration, and reactive astrocytosis—in the vicinity of neuritic plaques (**Figs 5-8, 10-13** and **Extended Figs 6.2, 11.1**). The intimate relations observed between ApoE, lipid-laden microglia and reactive astrocytes (**Extended Fig 6.2**) are consistent with known glial-mediated plaque clearance pathways. These observations underscore the cytoarchitectural and molecular complexity of the neuritic plaque niche in the terminal zone of the perforant path and add multiple new markers [ApoER2 LA1-2, HNE-ApoE, Reelin, Dab1, and Thr19-pPSD95] to the mix.

## DISCUSSION

### Overview of study findings

Sporadic AD lacks a unifying hypothesis that can account for the increase in lipid peroxidation observed early in the disease, the major role of ApoE (reflected by the enrichment of ApoE in the central core of neuritic plaques and link to APOE variants), the hallmark plaques and tangles, and the selective vulnerability of entorhinal-hippocampal structures. The present studies were motivated by the hypothesis that the convergence of reactive sites within synaptic ApoE receptors and their ligands (**Fig 2**) creates a microenvironment that is vulnerable to lipid peroxidation, and that resulting downstream disruptions in ApoE delivery and Reelin signaling contribute to the pathogenesis of sAD (**Fig 3**). We further proposed a sub-hypothesis wherein high ApoER2 expression—and constant need for Reelin/ApoE-ApoER2-Dab1 pathway activation to form memories—are molecular features that account for vulnerability of entorhinal-hippocampal structures in sAD (**Fig 1**). Using *in vitro* models, we provide proof-of-concept that ApoE and ApoER2 are vulnerable to attack by reactive lipid aldehydes, which generates lipid-protein adducts and stable ApoE-ApoER2 complexes. Using ISH, IHC, and MP-IHC we found that: (1) ApoER2 is highly expressed in the entorhinal-hippocampal memory system in humans (**Fig 5**), (2) ApoER2 LA1-2, ApoE and lipid aldehyde-modified ApoE, Reelin, Dab1 and downstream Reelin-Dab1 signaling partners [including Tyr607-pPI3K, Thr508-pLIMK1, Ser202/Thr205-pTau and Thr19-pPSD95] accumulate in the vicinity of neuritic plaques in sAD (**Figs 5-12**), (3) these pathologies were most prominent within the dense dendritic arbors of the molecular layers of the dentate gyrus and CA1-2, regions corresponding to the terminal zone of the perforant path projections that underlie memory formation (**Fig 1**), and (4) several ApoE/Reelin-ApoER2-Dab1 pathway markers positively correlate with antemortem cognitive deficits and histological progression in sAD (**Figs 5-11)**. Taken together, these observations reveal extensive derangements in the ApoE/Reelin-ApoER2-Dab1 axis within hippocampal structures that degenerate in sAD, and which is consistent with our hypothesis (**Fig 3**) wherein aldehydic adduction and crosslinking of ApoE and ApoER2 are initiating molecular lesions that compromise cytoskeletal and synaptic integrity and contribute to the histopathological hallmarks and cognitive deficits that characterize sAD in humans.

### Is a new interpretation of ApoE in sporadic Alzheimer’s disease needed?

APOE variants are the strongest genetic risk factor for sAD,[23–25, 98] however the most salient mechanisms mediating the link between ApoE and sAD remain elusive. ApoE is enriched in the core of newly formed Aβ-protein deposits,[99] implying a role for ApoE in the initial stages of plaque formation. Aβ is enriched in both diffuse plaques, which are not related to cognition,[100, 101] and in neuritic plaques that are closely correlated with cognitive deficits (see **Extended Fig 11.1**). By contrast, virtually all ApoE-immunoreactive plaques are associated with dystrophic neurites,[21, 22, 102] suggesting functional links between ApoE deposition and the genesis of neuritic plaques. In the present study, we used several independent antibodies to confirm that ApoE is enriched in the central core of neuritic plaques (**Figs 6-8, Extended Fig 11.1**), and accumulates in accordance with histological progression in sAD (**Figs 6-7**). Since neurons obtain ApoE particles via receptor-mediated internalization, one potential explanation for accumulation of ApoE in neuritic and newly formed plaques is compromised binding to ApoE receptors, which traps ApoE and its lipid cargo in the extracellular space. ApoER2 and VLDLR are of particular interest in this model because they can internalize ApoE particles,[39, 67, 103] are localized to synaptic membranes,[40, 41, 43, 104] and sit atop intraneuronal Reelin signaling cascades that govern synaptic integrity and function.[30–35]

### The Reelin-ApoER2 ligand-receptor pair in the perforant path target zone

Degeneration of the entorhinal-hippocampal memory system occurs early in sAD,[12–17] however the molecular determinants underlying this anatomical vulnerability are unknown. As the principal source of cortical input to the hippocampal formation (**Fig 1**), perforant path projections play a central role in learning and memory [6–11]. Collective evidence from rodents,[105] ferrets,[27] non-human primates,[28, 106] and humans,[29] indicates that neurons in layers II/III of the entorhinal cortex synthesize, anterogradely transport and secrete Reelin from axon terminals of the perforant path into the dense dendritic arbors within the molecular layers of the dentate gyrus and cornu ammonis (**Fig 1**). In rodent models, Reelin promotes dendritic arborization and long-term potentiation.[30-35, 42-46, 95-97, 107-109] Emerging evidence indicates that Reelin signaling also promotes adult hippocampal neurogenesis, and the migration and integration of adult-born neurons into hippocampal circuits.[110, 111] Moreover, Reelin-ApoER2-PSD95 signaling appears to be required to maintain stable post-synaptic architecture in mature hippocampal neurons.[112] Our finding that ApoER2 is highly expressed in the somata and dendrites of human dentate granule cells and a subset of CA1-2 pyramidal neurons points toward a central role for the Reelin-ApoER2 ligand-receptor pair in memory in humans, and raises the prospect that disruption of Reelin-ApoER2-Dab1 signaling pathways in this region could contribute to memory deficits in sAD. These observations and interpretation are also consistent with our sub-hypothesis that high ApoER2 expression, and a continuous need for Reelin-ApoER2-Dab1 pathway activation to shape and protect the synaptic connections underlying memory, are molecular features that account for the vulnerability of the terminal zones of the perforant pathway in sAD.

### Lipid peroxidation induced ApoER2 disruption as a proverbial ‘monkey wrench’ in the human memory system

We proposed that convergence of vulnerable lipid cargo transported by ApoE with reactive sites in ApoER2, creates potential for lipid peroxidation, and that ensuing adduction and crosslinking serve as a proverbial monkey wrench in the neural circuity underlying memory. Using a novel probe targeting the same double-Lys and His-enriched sequence within the ligand binding domain of ApoER2 LA1-2 that we previously found to be vulnerable to adduction and crosslinking (**Fig 4** and **Extended Fig 4**), we observed prominent, discrete ApoER2 LA1-2 deposits in the immediate vicinity of ApoE-enriched neuritic plaques within the dense dendritic arbors in the molecular layer of the dentate gyrus and CA1-2 regions (**Fig 5** and **Extended Figs 5-69-13**). Localized accumulation of ApoER2 LA1-2 here is noteworthy given that these dendritic arbors are the major synaptic recipients of the perforant path projections.[12, 15, 85] Tau-containing neuritic plaques in these terminal zones of the perforant path have previously been linked to memory deficits in sAD.[12, 13, 15, 17, 113, 114] Scheff et al. observed marked decreases in synaptic density and width [6, 7] of the molecular layer of the dentate gyrus in early AD. These collective observations support our proposal wherein ApoER2 disruption within the perforant path target zone plays a central role in sAD pathogenesis. Consistent with this, in 1999 Motoi et al. [115] reported increased expression of an ApoER2 cytoplasmic domain in unspecified hippocampal neurons in AD cases compared to non-AD controls. In the present study, we found that neuritic plaque associated accumulation of the ApoER2 LA1-2 (Reelin and ApoE binding) domain was much more robust than the beta-propeller and cytoplasmic domains in the perforant path terminal zones. This selective labeling suggests that the ApoER2 LA1-2 region may be intimately involved in the genesis of neuritic plaques.

### Concurrent accumulation of Dab1, ApoER2, Reelin and ApoE as evidence for a pathogenic nexus centered around ApoER2 disruption

Reelin and ApoE are both ligands for ApoER2, implying possible molecular interactions between ApoE and the Reelin-ApoER2-Dab1 axis. In rodent and cellular models, Reelin signaling through ApoER2 stabilizes the actin and microtubule cytoskeletons, and post-synaptic elements of adult hippocampal synapses (**Fig 2**).[30, 31, 34, 40, 42, 44] These stabilizing effects require binding of the Reelin double-Lys motif to the LA repeat 1 regions of ApoER2 and VLDLR [65, 66, 68, 70] leading to phosphorylation and activation of Dab1 (**Fig 2**).

Reelin signaling through ApoER2 and VLDR also triggers the degradation of Dab1, ApoER2 and Reelin itself (**Fig 2**).[40, 90]. Therefore, in the present study, the observed accumulation of Dab1 together with ApoER2 and its ligands Reelin and ApoE in the vicinity of neuritic plaques implies a localized, functional deficit in Reelin signaling through ApoER2. In experimental models, deficits in Reelin-ApoER2-Dab1 signaling also promote the formation of both hyperphosphorylated Tau and Aβ (reviewed in [116] and discussed below). Thus, ApoER2 receptor disruption is anticipated to compromise cytoskeletal and synaptic integrity, and to promote the plaques and tangles that define sAD.

### Peroxidation of ApoE and ApoER2 as initiating molecular lesions in sAD

A marked increase in lipid peroxidation is present in the earliest stages of sAD.[48, 49, 51–57, 117–119] Postmortem hippocampus specimens from sAD cases are reported to have a generalized increase in oxidative stress,[49] with a particularly strong signal in affected neurons and neuritic lesions.[50, 54, 57, 58] The human hippocampus is enriched in polyunsaturated phospholipids that are vulnerable to peroxidation,[59, 60, 120] Since ApoE is the main apolipoprotein that traffics these phospholipids to neurons,[18–20] it is not surprising that ApoE is vulnerable to peroxidation-related modifications.[121–124] However, the most relevant mechanisms and molecular targets linking ApoE peroxidation to sAD are not known. We observed that double-Lys and His-enriched sequences with ApoE and ApoER2 peptides, and full length ApoE and ApoER2 proteins, are susceptible to attack by reactive lipid aldehydes, inducing formation of both lipid-protein adducts and stable ApoE-ApoER2 complexes. Peptide crosslinking was dependent upon both the His (H140) residue and double-Lys (K143LRK146) motif of ApoE and either the double-Lys (K83K84) motif or the His (H75) residue of ApoER2. Since lipoprotein internalization requires binding of this double-Lys motif within ApoE to ApoE receptors, [65, 66, 68, 70] the observed adduction and crosslinking at these sites are expected to disrupt receptor mediated ApoE internalization. These findings provide a straightforward mechanism wherein peroxidation of the lipid cargo transported by ApoE could promote extracellular ApoE-Aβ plaque formation (see **Fig 3C-D** and *Unifying hypothesis* discussion below).

This interpretation is consistent with our finding that an immunoprobe targeting HNE-modified ApoE (HNE-ApoE) strongly labeled small, discrete puncta near the core of many ApoE and Aβ-enriched neuritic plaques, with only partial overlap with native ApoE labeling (**Figs 7, 12-13**). Together with our findings that HNE-ApoE was increased in sAD and positively correlated with histological progression and antemortem cognitive deficits, these observations suggest that a portion of ApoE present in the plaque core and surrounding neurons has been modified by lipid peroxidation, and provide a key pillar of support for our hypothesis linking ApoE peroxidation to sAD pathogenesis (**Fig 3**).

### Disruption of the ApoER2-Dab1-PI3K-LIMK1 actin stabilization pathway in sAD

Dendritic spines are dynamic actin-rich protrusions that harbor excitatory synapses within postsynaptic densities.[125] In rodent and cellular models, binding of Reelin to ApoER2 remodels and stabilizes the actin cytoskeleton of spines by evoking a kinase cascade involving phosphorylation of Dab1, Tyr607-pPI3K and Thr508-pLIMK1 (**Fig 2**).[92, 126–129] Despite evidence from experimental models, there is limited human data supporting a role for this pathway in sAD. Using IHC and MP-IHC to label each core component of this pathway, we observed strong expression of Tyr607-pPI3K in neuritic plaques and adjacent abnormal neurons (**Fig 10**), and of Thr508-pLIMK1 within a subset of neurons in affected regions. Heredia et al [130] previously observed a significant increase in the number of Thr508-pLIMK1 positive neurons in AD-affected regions, including some neurons that were devoid of hallmark AD pathologies. Similarly, we observed that Thr508-pLIMK1 expression was especially prominent in CA2-3 subregions, including neurons without evidence of Tau pathology. Moreover, Thr508-pLIMK1 and Tyr607-pPI3K prominently labeled granulovacuolar structures in affected regions that are consistent with granulovacuolar degeneration bodies. To our knowledge, Tyr607-pPI3K has not previously been shown to accumulate in neuritic plaques or in granulovacuolar structures. Since Reelin signaling through ApoER2 induces sequential Dab1, PI3K and LIMK1 phosphorylation (**Fig 2**), accumulation of Tyr607-pPI3K and Thr508-pLIMK1 in close proximity to Reelin and ApoE-enriched plaques provides additional evidence supporting disruption of this Reelin-ApoER2-PI3K-LIMK1 pathway and the putative pathogenic nexus shown in **Fig 3**.

### Disruption of the Reelin-ApoER2-Dab1-PI3K-GSK3β-Tau microtubule stabilization pathway in sAD

Local remodeling and stabilization of the microtubule cytoskeleton is also required to shape and protect synapses (reviewed in [35]). In experimental models, Reelin-ApoE receptor-Dab1 signaling stabilizes microtubules via a kinase cascade involving Tyr607-pPI3K-induced Ser9-phosphorylation and inhibition of GSK3β, which suppresses Tau phosphorylation (**Fig 2**). Compromised Reelin signaling through ApoE receptors promotes GSK3β-mediated Tau hyperphosphorylation and somatodendritic localization,[36, 38, 131, 132] which can be reversed by Reelin overexpression,[133] implying a direct link between deficits in Reelin-ApoE receptor signaling and this defining AD pathology. This interpretation, when considered together with our observations that Reelin, ApoER2, Dab1, and Thr508-pLIMK1 accumulate in close proximity to Tyr607-pPI3K and Ser202/Thr205-Tau in a subset of neuritic plaques (**Figs 5-12**), implies that parallel deficits in the actin and microtubule cytoskeletons can potentially be traced back to the disruption of Reelin signaling.

The best-known physiological role of Tau is stabilization of microtubules within terminal axons, however Tau also has important functions in dendrites.[134, 135] Dendritic Tau interacts with PSD95 [136] and is required to target Fyn to the post-synapse, where Fyn phosphorylates NMDA receptors [134, 137, 138] enabling formation of the PSD95-NMDAR complexes that mediate LTP.[138] Intriguingly, in experimental models neurotoxic effects of Aβ are wholly dependent on dendritic Tau and are abolished by its deletion.[134] Thr205-phosphorylation of Tau, which destabilizes microtubules, affords similar protection from Aβ.[139, 140] These observations, combined with evidence that the Reelin-ApoER2 cascades that are initiated upon Fyn phosphorylation of Dab1,[141, 142] are antagonized by extracellular Aβ,[88, 143, 144] imply physiological roles for Aβ and Thr205-phosphorylation of Tau in the fine-tuning of Reelin-ApoER2-Dab1 mediated synaptic plasticity, and in protecting neurons from excitotoxicity.[135] These findings raise the prospect that accumulations of extracellular Aβ and intraneuronal Thr205-pTau could reflect chronic or maladaptive protective responses, which ultimately manifest as the hallmark plaques and tangles that define AD.

### Disruption of the Reelin-ApoER2-Dab1-PI3K-GSK3β-PSD95 postsynaptic receptor complex stabilization pathway in sAD

In rodent models, Reelin deficiency has been shown to deplete PSD95 at the post-synapse.[34] Activated GSK-3β also phosphorylates PSD95 at Thr19 to induce synapse disassembly and LTD,[72] suggesting that like Tau, PSD95 may be regulated by this Reelin-ApoE receptor-Dab1 pathway (**Fig 2**). In the present study, we observed striking immunoreactivity for Thr19-pPSD95 in MAP2-positive dystrophic dendrites in proximity to ApoE- and Reelin-enriched plaques and adjacent neurons in regions affected by sAD (**Figs 11-12**), major accumulation of Thr19-pPSD95 in sAD cases, and strong positive correlations between Thr19-pPSD95 and histological progression and antemortem cognitive deficits (**Figs 11-12**). To our knowledge this is the first demonstration of Thr19-pPSD95 immunolabeling in human entorhinal-hippocampal structures, and the first evidence linking Thr19-pPSD95 to AD. These observations suggest that compromised Reelin-ApoE receptor-Dab1-PI3K signaling could enhance GSK3β-mediated phosphorylation of both Tau and PSD95, leading to parallel destabilization of the microtubule cytoskeleton and postsynaptic receptor clusters within excitatory synapses, respectively. Anticipated functional correlates of such alterations may include enhanced LTD and synaptic dysfunction, while long-term pathological correlates may include synaptic loss with accumulation of Ser202/Thr205-pTau and Thr19-pPSD95, as observed in this study. Since synapse dysfunction and loss are implicated in memory deficits in sAD,[6, 7, 145] Thr19-phosphorylation of PSD95 should be studied further as both a potential underlying mechanism and biomarker for AD.

### Postsynaptic receptor complexes as a locus for ApoE/Reelin-ApoER2-Dab1 pathologies in sAD

The splice variant of ApoER2 that enables formation of postsynaptic ApoER2-PSD95-NMDA receptor complexes [40–43, 94] helps account for the pivotal role of ApoER2 in LTP and memory.[94–97] In the present study, we observed colocalization of Thr19pPSD95 and MAP2 (**Fig 11-12**), and localized enrichment of ApoER2, Dab1, Thr508-pLIMK1, Tyr607-pPI3K and Ser202/Thr205-pTau (**Figs 5-13**) in the vicinity of ApoE-, Reelin- and Aβ-enriched neuritic plaques. These observations, together with published data from experimental models summarized above, suggest that ApoER2 could serve as a molecular integrator, transmitting signals from extracellular Reelin and ApoE (and Aβ) to postsynaptic elements of excitatory synapses by regulating ApoER2-PSD95-NMDA-Tau complex integrity, Reelin-ApoER2-Dab1 signal transduction, and ultimately the balance between LTP and LTD. Thus, we propose a model (**Fig 3**) wherein chronic disruption of ApoER2 at the post-synapse leads to both synapse failure and the hallmark sAD pathologies including extracellular ApoE-Aβ complexes and intraneuronal neurofibrillary tangles.

### ApoE and ApoE receptor peroxidation and endolysosomal pathway dysfunction

Enlargement of neuronal endolysosomal compartments is among the earliest pathologies in sAD.[146–148] Following receptor binding, ApoE-ApoE receptor complexes are internalized and trafficked to the early endosome, where the acidic environment (pH 6) facilitates ligand release,[67] allowing for swift recycling of ApoE receptors to cell surface.[149, 150]. In experimental models, the APOE4 allele compromised ApoER2 recycling [149, 150], which was reversed by pharmacological lowering of endosomal pH. These observations were attributed to the propensity of ApoE4 to acquire a ‘molten-globule’ state near its isoelectric point [149, 150]. In the present study we observed that: (1) ApoE and ApoER2 peptides and proteins are vulnerable to attack by reactive lipid aldehydes, and (2) the resulting lipid-aldehyde adducts and crosslinked complexes were partially (but incompletely) reversible in acidic conditions modeling the early endosome and lysosome (**Fig 4**). Together, these observations suggest that lipid peroxidation may contribute to sequestration and compromised pH-dependent dissociation of ApoE and ApoER2 in endolysosomal compartments (**Fig 3F**), impaired ApoER2 recycling, and ultimately to deficits in ApoE-mediated lipid delivery and Reelin-ApoER2-Dab1 signaling.

### Linking ApoE peroxidation and ApoER2 disruption to formation and clearance of Aβ plaques

ApoER2 and AβPP share several intracellular adaptor proteins that regulate internalization and trafficking.[151–154] In experimental models, the binding of ApoE and Reelin to ApoER2 regulates internalization of AβPP and BACE1, the rate-limiting enzyme required for conversion of AβPP to Aβ.[151, 152] These observations suggest that disruptions in ApoE/Reelin-ApoER2-Dab1 signaling may alter Aβ production,[152] however mechanisms linking lipoprotein receptors to AD are complex and incompletely understood (reviewed in [155–158]).

The neurotoxic effects of high (μM) concentrations of Aβ are well-recognized and perhaps best exemplified by the aggressive course of familial AD,[159] which is caused by genetic mutations that increase the synthesis of Aβ. Although usually overlooked, substantial evidence indicates that Aβ has protective functions at physiological (<1 nM) concentrations.[160–163] Moreover, unlike the genetic source of Aβ overproduction in familial AD (and transgenic mouse models), the most salient trigger(s) for Aβ synthesis and deposition in sAD are unknown. BACE1 is activated by oxidative stress and other cellular stressors,[164–168] implying a protective role for Aβ against noxious stimuli. BACE1 is most active in acidic organelles.[169, 170] In sAD, altered endolysosomal trafficking leads to colocalization of AβPP and BACE1 in late endosomes, facilitating Aβ synthesis.[171] Intriguingly, at physiological concentrations, Aβ acts as a potent antioxidant and metal chelator [160–163, 172] that is localized to lipoproteins, and appears to protect vulnerable polyunsaturated cargo from peroxidation.[160, 173] In *in vitro* models Aβ interacts with the lipid cargo-transporting domain of ApoE.[174] These collective observations led us to speculate that in neurons, peroxidation-modified ApoE particles could serve as a noxious stimulus leading to endolysosomal compromise, and that ensuing BACE1-AβPP colocalization could serve a protective function by increasing secretion of Aβ monomers to bind and neutralize peroxidized ApoE particles. However, with excessive or prolonged exposure, these protective effects of Aβ could transition to the well-established neurotoxic effects of high Aβ concentrations, overwhelming glial-mediated Aβ clearance pathways, leading to extracellular Aβ deposition. This nuanced interpretation wherein BACE1 and Aβ monomers serve a protective role is consistent with findings from controlled trials in which BACE1 inhibitors paradoxically worsened cognitive decline,[175, 176] despite markedly decreasing CSF Aβ (see description in **Table S1**).[177]

Consistent with this model, using MP-IHC we observed complex spatial and morphological relationships between Aβ, ApoE and lipid aldehyde-modified ApoE, Reelin and ApoER2 LA1-2 in the vicinity of neuritic plaques (**Figs 5-8, 11-13**). Moreover, we observed extensive microglial infiltration and reactive astrocytosis (**Fig 13, Extended Fig 6.2**), consistent with the known role of Aβ in triggering microglia and astrocyte-mediated clearance pathways.[178, 179] Since neurons obtain ApoE particles via receptor-mediated internalization, our MP-IHC observations, when considered together with published evidence that oxidized LDL [180] and ApoE (and especially ApoE in the presence of oxygen)[174] promote Aβ oligomerization,[174, 181, 182] suggest that aldehyde-induced ApoE receptor disruption could trap peroxidized ApoE particles outside of neurons, where they could provide seeds for Aβ oligomerization, deposition and ultimately neuritic plaque formation (**Fig 3C-D**). This proposed sequence of events is consistent with evidence that in sAD, ApoE is enriched in newly formed [99] and neuritic plaques [21] and suggests that the sequence of molecular events underlying plaque formation in sAD (ApoE receptor disruptionàlocalized ApoE depositionàApoE-Aβ complex formation) may be distinct from familial AD and transgenic mouse models (Aβ overproductionàAβ deposition). Future mechanistic studies are needed to determine if this proposed sequence of events is valid. Future IHC studies comparing plaque contents of sporadic versus familial AD cases could shed further light on this topic.

### Unifying hypothesis and potential implications for therapeutics and prevention

The *in vitro* and IHC observations presented here provide evidence supporting a unifying hypothesis (**Fig 3**), wherein ApoE and ApoE receptor peroxidation, aldehydic adduction and crosslinking contribute to sAD pathogenesis in humans by: (1) disrupting ApoE receptor-dependent delivery of lipid cargo required to remodel synaptic membranes; (2) trapping peroxidized ApoE particles outside neurons, where they provide seeds for Aβ oligomerization; (3) disrupting Reelin-ApoE receptor signaling cascades that stabilize actin and microtubule cytoskeletons and postsynaptic receptor complexes; (4) promoting Tau hyperphosphorylation and neurofibrillary tangle formation; (5) disrupting endolysosomal trafficking, which in turn; (6) increases Aβ synthesis and exacerbates proteinopathies. This model reframes synthesis and secretion of Aβ, and Thr205-phosphorylation of Tau as physiological responses involved in fine-tuning synaptic activity and protecting neurons from lipid peroxidation that later contribute to the formation of plaques, tangles and neurodegeneration in sAD. *This unifying hypothesis could help account for the findings of the present study, as well as the selective anatomical vulnerability, histopathological hallmarks, loss of cytoskeletal and synaptic integrity, and cognitive deficits that characterize sAD*. The hypothesis is consistent with major genetic risk factors for sAD—including variants in genes coding for ApoE, endolysosomal pathway components, and glial pathways that regulate phagocytosis and clearance of ApoE, Aβ, and lipids. The pathogenic nexus (**Figs 2-3**) is also a convergence point for multiple established and suspected environmental risk factors for sAD (**Table S1**). This hypothesis is testable, and if experimentally corroborated, could have broad implications for the prevention and management of sAD. Therapeutic implications and proposed pathogenic factors underpinning the ApoE-ApoER2 peroxidation cascade versus the Amyloid Cascade hypothesis for sAD are summarized in **Table S1**.

### Strengths & limitations

Major strengths of the present study include: (1) The use of rapidly autopsied tissue specimens with short postmortem intervals that underwent uniform fixation [73, 183, 184], in conjunction with antemortem cognitive data, (2) the use of IHC-validated antibodies whenever available, (3) extensive validation of custom-designed antibodies using a multifaceted, orthogonal approach comprising western blot, ISH/RNA-protein co-detection, single-marker IHC, and multi-epitope labeling enabling comparison of staining patterns with several independent antibodies, and (4) the spatial, morphological, cytoarchitectural context provided by MP-IHC. The moderate sample sizes (n=26 for most markers) are an important limitation of these IHC studies. Future, larger studies are needed to determine if results are influenced by APOE variants and other variables. Findings provide proof-of-concept that the specific double-Lys motifs and His moieties within ApoE and ApoER2 are vulnerable to adduction and crosslinking. However, since each of these full-length proteins contain additional reactive amines, future studies are needed to clarify the most relevant sites, types, and extent of lipid peroxidation-related modifications. Higher magnification imaging is needed to determine the subcellular localizations of markers. Future studies are also needed to establish the sequence of molecular events leading to accumulation of markers investigated here, and whether such accumulations are a cause or consequence of sAD.

### Summary and Conclusion

Collective findings identify functional domains of ApoE and ApoER2 that are vulnerable to lipid peroxidation and provide evidence for a pathogenic nexus centered around ApoER2 disruption, with neuritic plaque-associated accumulation of ApoER2, its extracellular ligands ApoE and Reelin, and multiple downstream Reelin/ApoE-ApoER2-Dab1 signaling partners, within the human entorhinal-hippocampal memory system in sAD. These findings identify missing molecular links and provide the basis for a unifying hypothesis that could help explain the selective anatomical vulnerability, genetic risk factors, neuropathological features, and clinical presentation of sAD in humans.

## AUTHOR CONTRIBUTIONS

CER completed the literature review and originated the hypothesis, directed the project, and wrote the first draft of the manuscript. DM led the MP-IHC experiments and contributed to hypothesis refinement and manuscript preparation. GSK and LC led the chemistry and molecular biology experiments and contributed to hypothesis refinement and manuscript preparation. MSH managed the data and images, performed annotations and image analysis, and contributed to manuscript preparation. AS performed the MP-IHC experiments and JJ performed MP-IHC post-acquisition image processing and analysis. DZ led the statistical analysis and contributed to manuscript preparation. RM and DK contributed to manuscript preparation.

## Data Availability

The datasets and code are available from the corresponding author upon reasonable request.

## ACKNOWLEDGEMENTS

This work was supported by the intramural programs of the National Institute on Aging (NIA), National Institute on Alcohol Abuse and Alcoholism (NIAAA) and National Institute on Neurological Diseases and Stroke (NINDS), NIH. Additional support was provided as a research gift from John M. Davis to the Laboratory of Clinical Investigation, NIA/NIH. We are grateful to the Banner Sun Health Research Institute BBDP of Sun City, Arizona for the provision of human brain tissue. The BBDP is supported by the NINDS (U24 NS072026 National Brain and Tissue Resource for Parkinson’s Disease and Related Disorders), the NIA (P30 AG19610 Arizona Alzheimer’s Disease Core Center), the Arizona Department of Health Services (contract 211002, Arizona Alzheimer’s Research Center), the Arizona Biomedical Research Commission (contracts 4001, 0011, 05-901 and 1001 to the Arizona Parkinson’s Disease Consortium) and the Michael J. Fox Foundation for Parkinson’s Research. We gratefully acknowledge the volunteer participants and their families who provided postmortem tissues, as well as Geidy Serrano, Thomas Beach and the BBDP on-call autopsy team who provided this extraordinary resource. We acknowledge Marc Raley (Visual Media Services for intramural programs of NIDA and NIA) for graphic art in Fig 1. We thank Pragun Vohra and the Histoserv staff for immunohistochemical expertise and immunostaining, Scott Lewis and New England Peptide/Vivitide staff for expert input into antibody development, purification, and validation, and Precision Antibody for expert input into monoclonal antibody development. We are grateful to Josephine Egan (Chief, Laboratory of Clinical Investigation, NIA) for editing the manuscript. The authors acknowledge the valuable published work of investigators that provided a foundation for the present hypotheses.

