## Supplement for "Lipid peroxidation induced ApoE receptor-ligand disruption as a unifying hypothesis underlying sporadic Alzheimer’s disease in humans"


I. Extended Figures

II. Supplementary Tables

III. Supplementary Materials and Methods

### I. Extended Figures

**Extended Fig 2.1. Multidomain antibody and ISH labeling strategy for ApoER2 and ApoE.**

**Extended Fig 4.1. Time of Flight Mass Spectrometry (TOF-MS) evidence for 4-ONE-generated crosslinked ApoER2-ApoE heterodimers and intra-chain crosslinks within ApoE and ApoER2 peptides.**

**Extended Fig 4.2. Lipid aldehyde-crosslinked ApoER2-ApoE complexes are detected by high molecular weight protein migration and antibodies targeting ApoE, ApoER2 and Cu-oxidized PAPC-ApoE.**

**Extended Fig 4.3. Lipid aldehyde-induced crosslinking generates ApoE multimers that are detected by high molecular weight protein migration and antibodies targeting aldehyde-modified ApoE and phospholipid peroxidation-modified ApoE.**

**Extended Fig 5.1. ApoER2 LA1-2 accumulates as discrete aggregates in the SP and SR subfields of CA1-2 in sporadic AD**

**Extended Fig 5.2. Fig 3. Multi-epitope immunolabeling and ISH/RNA-protein codetection of ApoER2 in proximity to ApoE-enriched plaques in the molecular layer of dentate gyrus and hippocampus.**

**Extended Fig 5.3. Selective labeling of ApoER2 via western blot and ApoER2 RNA-protein codetection assays.**

**Extended Fig 6.1. ApoE accumulates in the vicinity of its receptor ApoER2 in the perforant path terminal zones in sporadic AD.**

**Extended Fig 6.2. Close spatial relationships between ApoE, lipid-laden microglia and reactive astrocytes in the vicinity of neuritic plaques.**

**Extended Fig 7.1. HNE-ApoE accumulates in the temporal cortex in sporadic AD.**

**Extended Fig 7.2. HNE-ApoE and native ApoE accumulate in the vicinity of ApoER2 LA 1-2 in the outer molecular layer of the dentate gyrus.**

**Extended Fig 9.1. Dab1 plaque complexes accumulate in the cornu ammonis in sporadic AD.**

**Extended Fig 10.1. Accumulation of ApoER2 LA1-2, Dab1, Tyr607-pPI3K, Thr508-pLIMK1 in the CA2 subregion of the hippocampus in sAD**.

**Extended Fig 11.1 Diffuse plaques are enriched in Aβ but lack the ApoE and Thr19-pPSD95 that are abundant in neuritic plaques.**

**Extended Fig 13.1. IHC evidence for convergence of ApoE/Reelin-ApoER2-Thr19-pPSD95 axis pathologies in the perforant path target zone in early sAD.**

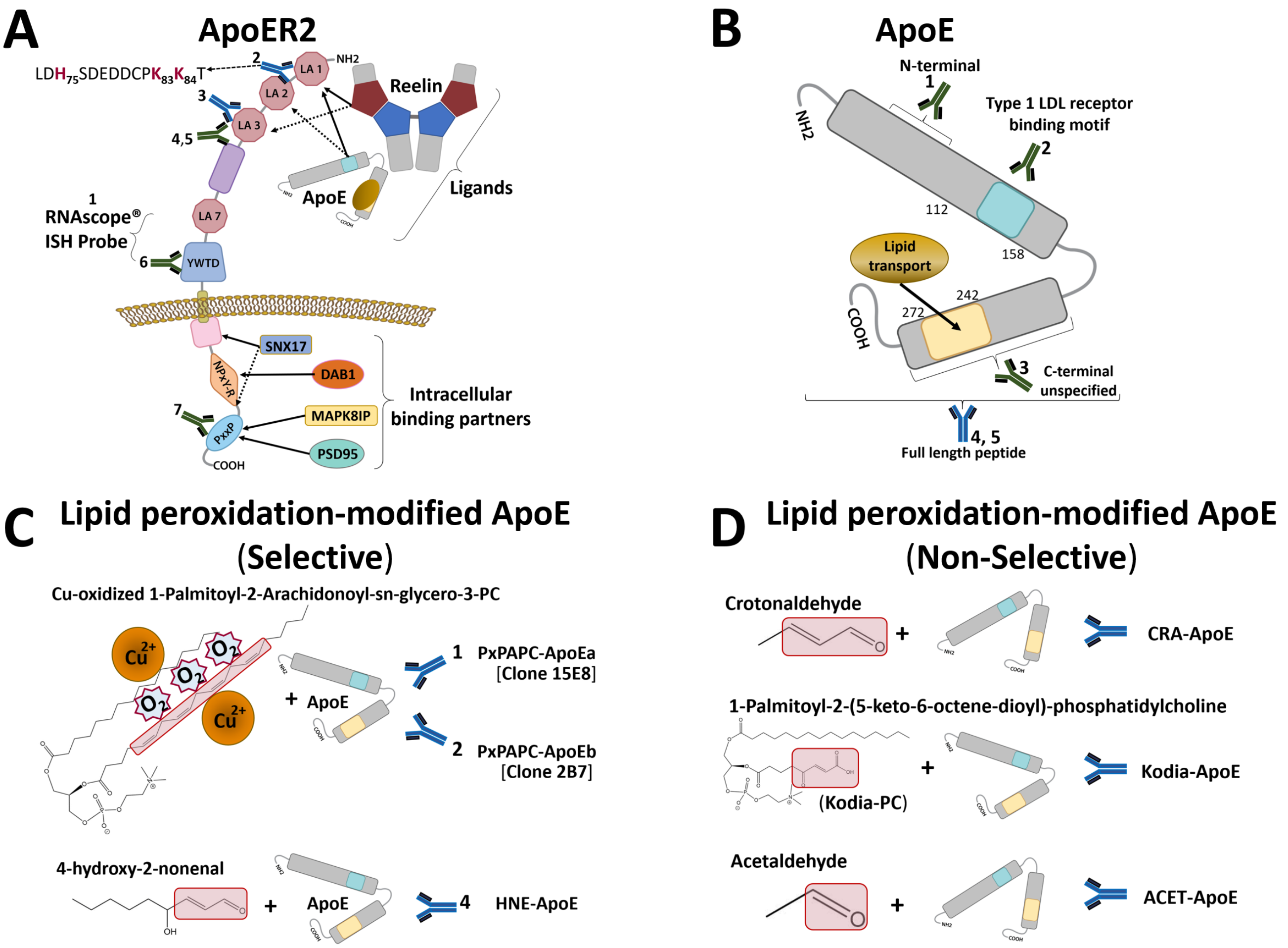


**Extended Fig 2.1. Multidomain antibody and ISH labeling strategy for ApoER2 and ApoE.** To investigate key molecular components of the the hypothesized pathological nexus underlying AD (**Figs 2-3**), we applied a multidomain labeling strategy with 18 probes targeting different domains or epitopes within ApoER2 (1 ISH probe, 6 antibodies) (**A**), native ApoE (5 antibodies) (**B**), and lipid-peroxidation modified ApoE (6 antibodies [3 that selectively detect modified ApoE, 3 that also detect native ApoE]) (see **Table S1**). The use of several antibodies targeting each protein, together with ISH/RNA-protein codetection, assisted in the validation of observations made using single antibodies.

**ApoER2** (**A**): The ISH probe used for RNA-protein codetection targeted the midchain region of ApoER2 that is common to major splice variants. Immunoprobes targeted the double-Lys and His-enriched sequence between LDL receptor type A repeats 1 and 2 (ApoER2 LA1-2) that is adjacent to the Reelin binding domain,[1] the LA3 region that participates in lipoprotein binding,[2] the β-propeller 'YWTD' domain that is essential for endolysosomal lipoprotein-receptor complex dissociation, [3, 4], and the proline-rich motif in the cytoplasmic tail that mediates binding to MAPK8IP1 (JIP-1)[5] and anchors ApoER2 to lipid rafts,[6, 7] enabling formation of multiprotein ApoER2-PSD95-NMDA receptor complexes.[8-10]

**Native ApoE** (**B**): Immunoprobes targeted the ApoE receptor binding domain, amino acids 51-100 in the N-terminal region, the C-terminal lipid cargo transporting region, and the full-length peptide.

**Lipid-peroxidation modified ApoE (Selective, C)**: Immunoprobes that selectively detected peroxidation-modified ApoE with minimal or no detection of unmodified ApoE in western blot assays include two monoclonals raised against Cu-oxidized PAPC-modified ApoE (PxPAPC-ApoEa [Clone 15E8]; PxPAPC-ApoEb [Clone 2B7]), and the chicken polyclonal antibody raised against ApoE treated with 4-hydroxy-2-nonenal (HNE-ApoE) (see **Fig 4** and **Extended Figs 4.2-4.3**).

**Lipid-peroxidation modified ApoE (Non-selective, D)**: Antibodies raised against crotonaldehyde (CRA-ApoE), KOdiA-PC (KOdiA-ApoE), and acetaldehyde (ACET-ApoE) detected both peroxidation-modified ApoE and unmodified ApoE (see **Fig 4** and **Extended Figs 4.2-4.3**), and therefore were not selective. Commercially available antibodies and new antibodies are depicted in green and blue colors, respectively.

### Extended Fig 2.1 References

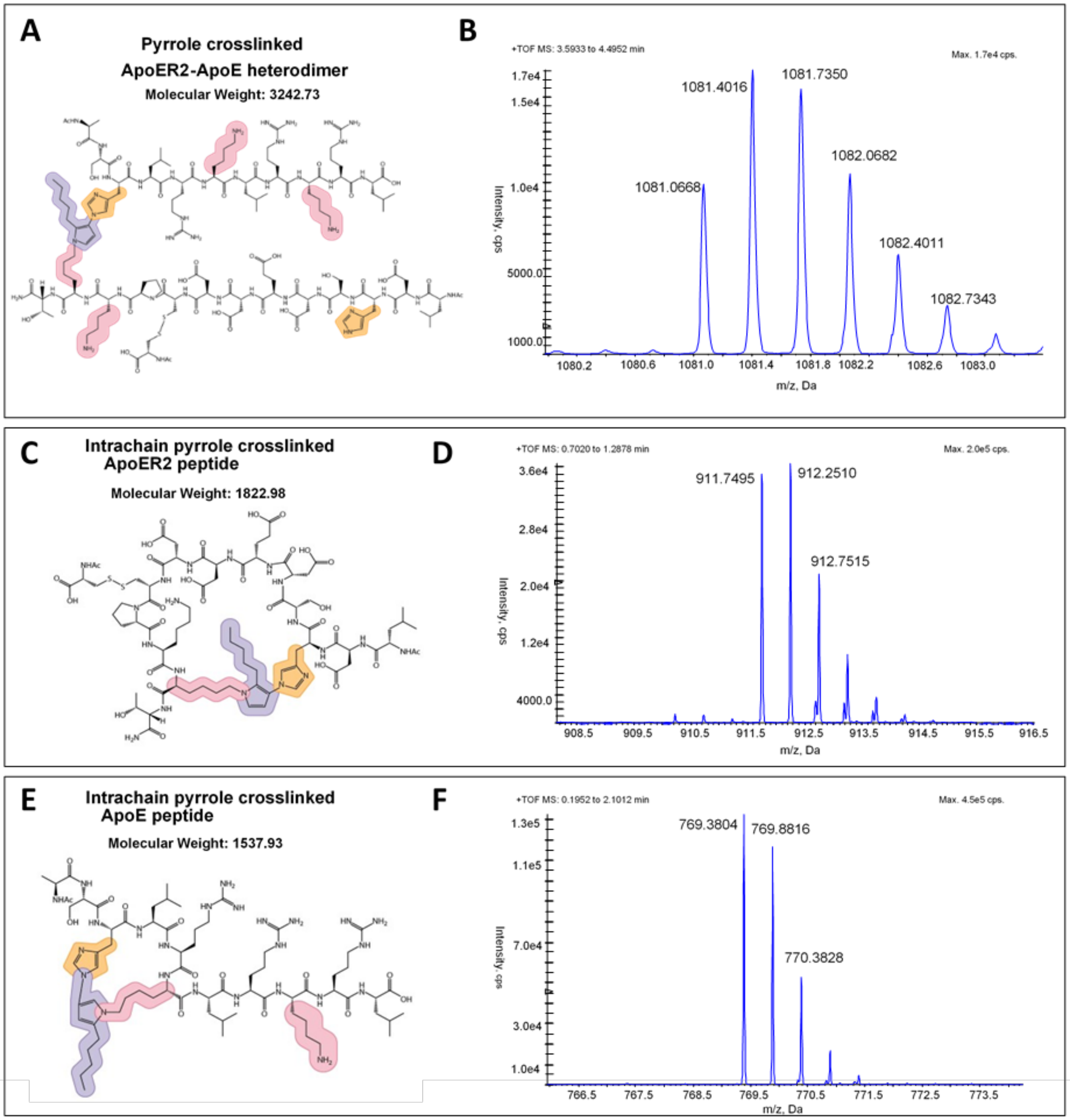


**Extended Fig 4.1. Time of Flight Mass Spectrometry (TOF-MS) evidence for 4-ONE-generated crosslinked ApoER2-ApoE heterodimers and intra-chain crosslinks within ApoE and ApoER2 peptides.** TOF-MS results for incubations of 4-ONE and peptides within the binding regions of ApoER2 and ApoE reveal pyrrole heterodimer crosslinks (**A-B**) and intrachain crosslinks (**C-D, E-F**). The crosslinked ApoER2-ApoE peptide was observed as its triple charged ion (MW = 3242, $[(M+3H^{+})/3= 1081]$ (**B**). Intrachain crosslinks within the ApoER2 peptide (**C-D**; (MW=1822, $[(M+2H^{+})/2= 912]$) and the ApoE peptide (**E-F**; (MW=1537, $[(M+2H^{+})/2= 769]$) were observed as their double charged ions as illustrated by the charged isotope peaks in each spectra. The crosslinked ApoER2-ApoE peptide was not reversible at pH 4.0, further suggesting likely pyrrole formation. Yellow and pink shading in **A, C** and **E** designate the reactive amines present in His and Lys, respectively. Purple shading designates aldehydic products of lipid peroxidation. Several pyrrole crosslinks are possible, illustrative example structures are shown in **A, C** and **E**. Corresponding crosslinked heterodimers were detected when the ApoER2$_{H\rightarrow A}$ analog or the ApoER2$_{K\rightarrow A}$ analog were incubated with the native ApoE peptide. No crosslinked heterodimers were detected with either of the ApoE analogs (ApoE$_{H\rightarrow A}$, ApoE$_{KLRK\rightarrow ALRA}$), or when ApoER2 peptide lacked His. Together, these findings imply a requirement for both His and Lys residues on ApoE and either a His or Lys on ApoER2 for crosslinking.

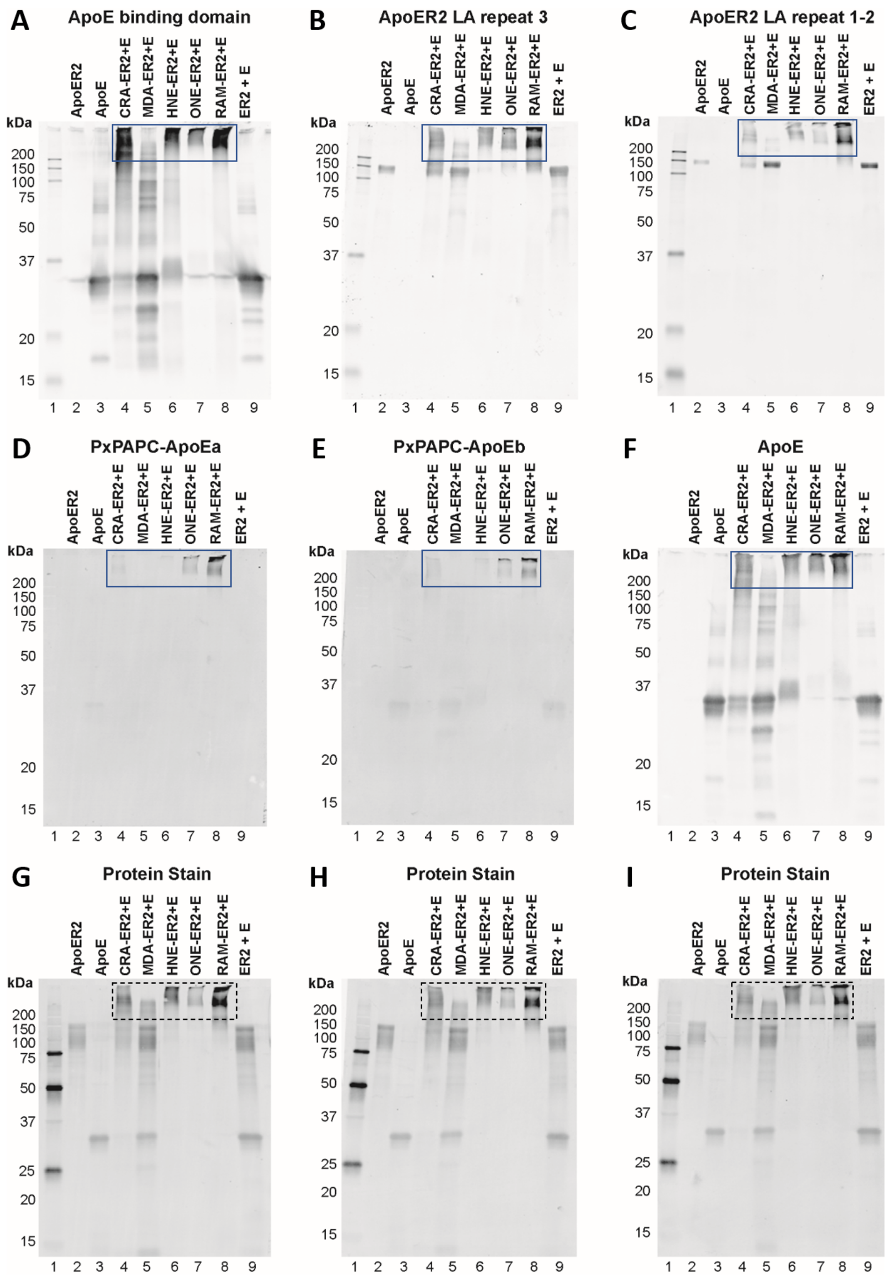


**Extended Fig 4.2. Lipid aldehyde-crosslinked ApoE-ApoER2 complexes are detected by high molecular weight protein migration and antibodies targeting ApoE, ApoER2 and Cu-oxidized PAPC-ApoE.** Crosslinking of ApoE and ApoER2 proteins was detected by fluorescence immunoblotting against the ApoE binding domain (**A**), full-length ApoE (ApoE, **F**), ApoER2 LA repeat 3 (**B**), ApoER2 LA repeat 1-2 (**C**) and Cu-oxidized PAPC-modified ApoE (PxPAPC-ApoEa [**D**] and PxPAPC-ApoEb [**E**]). 250ng of ApoER2 ectodomain protein (lane 2) and ApoE monomer (lane 3) were loaded per gel to compare with 500ng of protein from each combined lipid aldehyde reaction. Crotonaldehyde (Lane 4, CRA-ER2+E), malondialdehyde (Lane 5, MDA-ER2+E), 4-hydroxy-2-nonenal (lane 6, HNE-ER2+E), 4-oxo-2-nonenal (lane 7, ONE-ER2+E), a reactive aldehyde mixture including CRA, MDA, HNE, ONE and acrolein (lane 8, RAM-ER2+E), and ApoER2 and ApoE monomer co-incubated without aldehydes (lane 8, ER2+E) are represented in each immunoblot. **G-I**, total protein per membrane was detected by AzureRED total protein stain. Blue boxes (**A-F**) denote higher molecular weight proteins detected. Black boxes (**G-I**) denote higher molecular weight protein migration. Western blots are representative of experiments performed three times.

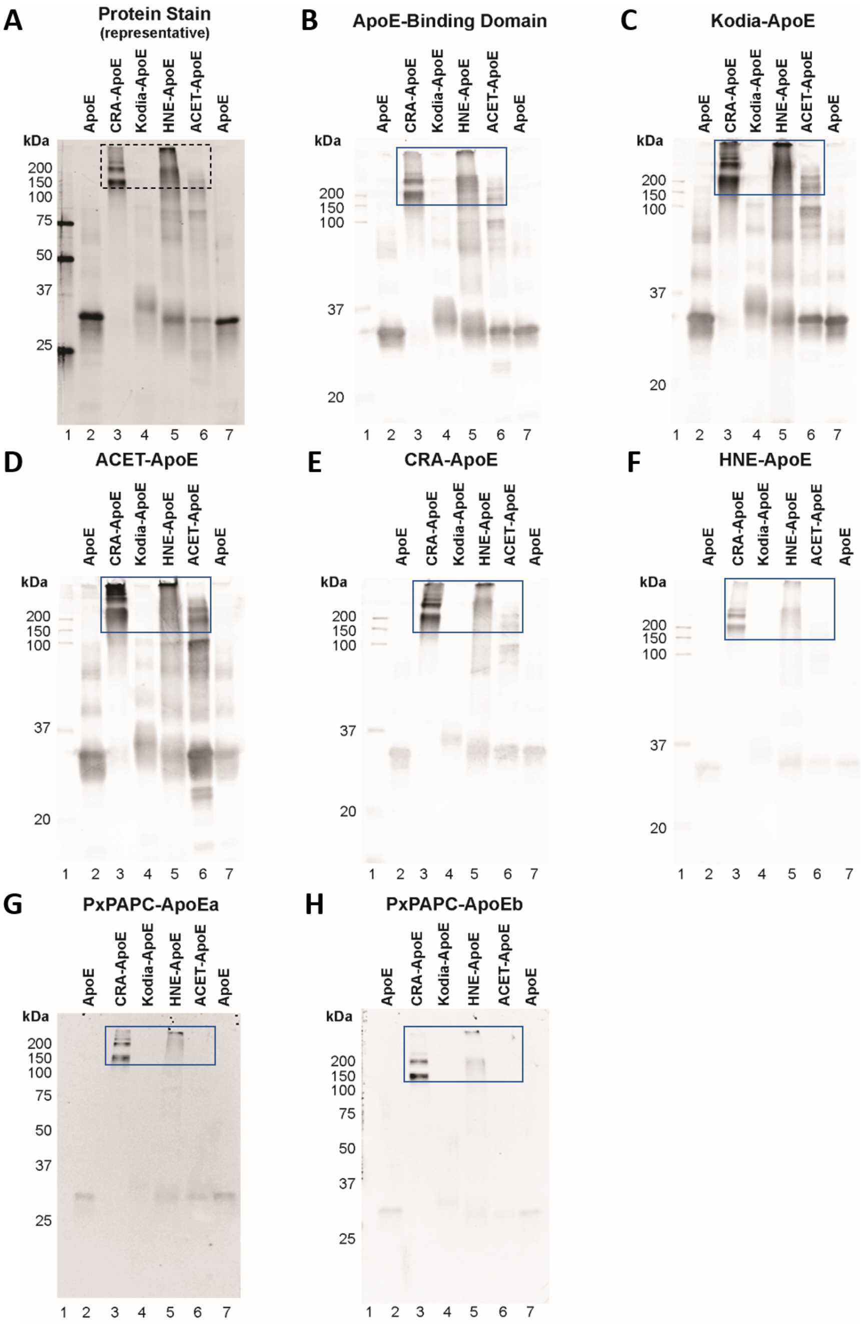


**Extended Fig 4.3. Lipid aldehyde-induced crosslinking generates ApoE multimers that are detected by high molecular weight protein migration and antibodies targeting aldehyde-modified ApoE and phospholipid peroxidation-modified ApoE.** Aldehyde-induced crosslinking of ApoE monomers was detected by high molecular weight protein migration (**A**) and fluorescence immunoblotting against the ApoE binding domain (**B**), KOdiA-PC modified ApoE (KOdiA-ApoE, **C**), acetaldehyde-modified ApoE (ACET-ApoE, **D**), crotonaldehyde-modified ApoE (CRA-ApoE, **E**), 4-hydroxy-2-nonenal (HNE)-modified ApoE (HNE-ApoE, **F**), and two antibodies targeting Cu-oxidized PAPC-modified ApoE (PxPAPC-ApoEa [**G**] and PxPAPC-ApoEb [**H**]). 250ng of ApoE protein (lane 2, A) and ApoE monomer (lane 7, A) were loaded per gel to compare with 500ng of protein from each combined lipid aldehyde reaction. CRA (Lane 3, CRA-ApoE), KOdiA-PC (Lane 4, KOdiA-ApoE), HNE (lane 5, HNE-ApoE), or acetaldehyde (lane 6, ACET-ApoE), are represented in each immunoblot. Protein migration per membrane was detected by AzureRED total protein stain. Blue boxes (**B-H**) denote higher molecular weight proteins detected. Black box (**A**) denotes higher molecular weight protein migration. Western blots are representative of experiments performed three times.

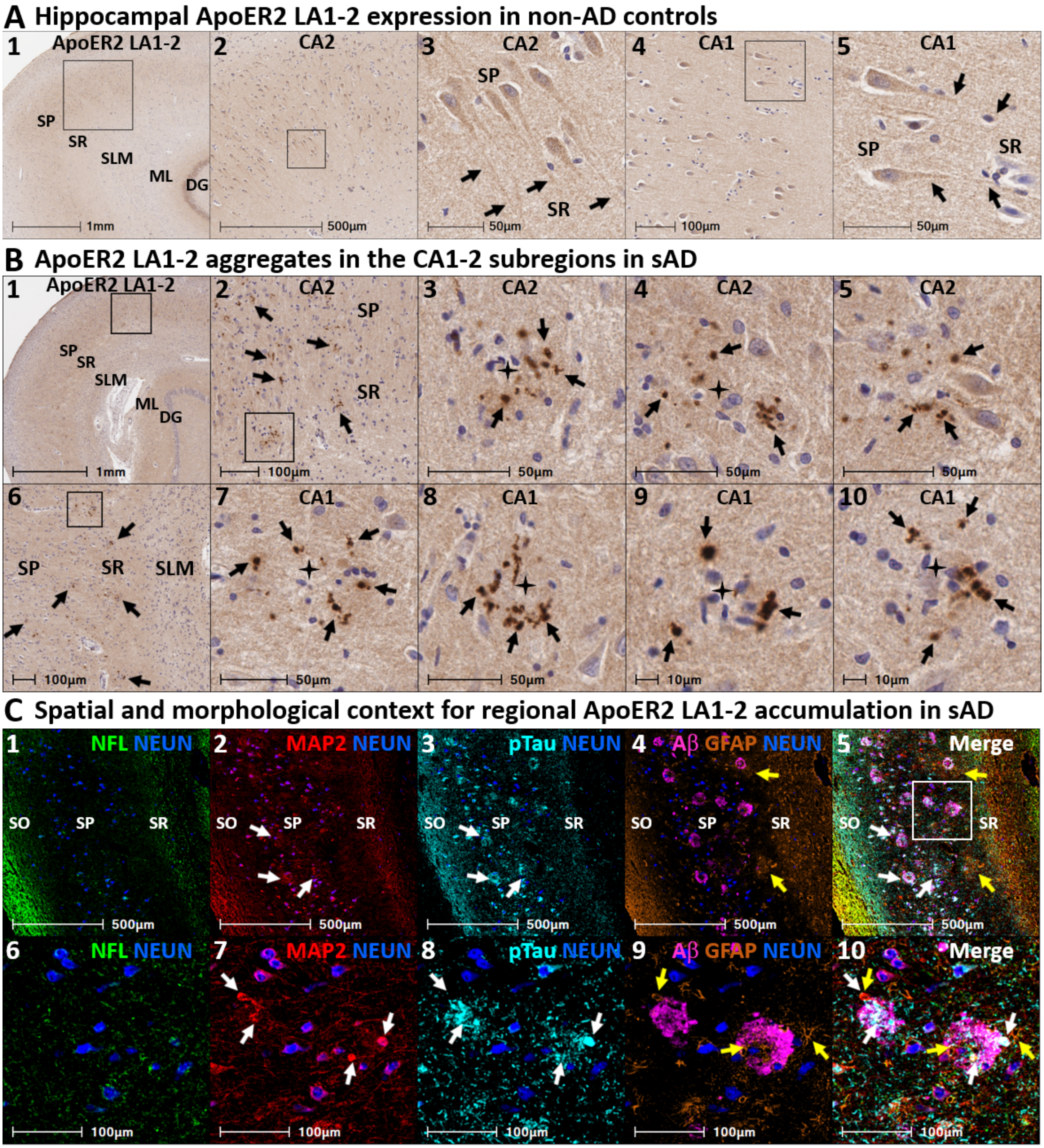
A Hippocampal ApoER2 LA1-2 expression in non-AD controls
1 ApoER2 LA1-2
SP
SR
SLM
ML
DG
1mm
2 CA2
500µm
3 CA2
SP
SR
50µm
4 CA1
100µm
5 CA1
SP
SR
50µm
B ApoER2 LA1-2 aggregates in the CA1-2 subregions in sAD
1 ApoER2 LA1-2
SP
SR
SLM
ML
DG
1mm
2 CA2
SP
SR
100µm
3 CA2
50µm
4 CA2
50µm
5 CA2
50µm
6
SP
SR
SLM
100µm
7 CA1
50µm
8 CA1
50µm
9 CA1
10µm
10 CA1
10µm
C Spatial and morphological context for regional ApoER2 LA1-2 accumulation in sAD
1 NFL NEUN
SO
SP
SR
500µm
2 MAP2 NEUN
SO
SP
SR
500µm
3 pTau NEUN
SO
SP
SR
500µm
4 Aβ GFAP NEUN
SO
SP
SR
500µm
5 Merge
SO
SR
500µm
6 NFL NEUN
100µm
7 MAP2 NEUN
100µm
8 pTau NEUN
100µm
9 Aβ GFAP NEUN
100µm
10 Merge
100µm

**Extended Fig 5.1. ApoER2 LA1-2 accumulates as discrete aggregates in the SP and SR subfields of CA1-2 in sporadic AD.** Panels **A**$_{1\text{-}5}$ and **B**$_{1\text{-}10}$ are coronal sections through the CA1-2 subregions from a non-AD control case and a (Braak stage V, ApoE3/3) sAD case, respectively. Controls (**A**$_{1\text{-}5}$) exhibit diffuse, homogenous ApoER2 LA1-2 expression in the CA1-2 SP and SR subregions. Arrows in **A**$_{3}$ and **A**$_{5}$ depict ApoER2 LA1-2 positive apical dendrites emanating into the SR regions of CA2 and CA1, respectively. By contrast, sAD cases exhibit numerous discrete ApoER2 LA1-2 aggregates (black arrows in **B**$_{1\text{-}10}$) throughout the SP and SR subfields of CA1-2 (**B**$_{1\text{-}10}$). ApoER2 LA 1-2 aggregates accumulate in the vicinity of plaques (black stars). MP-IHC labeling (**C**$_{1\text{-}10}$) in a serial section from the same sAD case provides cytoarchitectural context and reveals the presence of hallmark AD pathologies—including Aβ and Ser202/Thr205-pTau containing neuritic plaques and neurofibrillary tangles (**C**$_{3\text{-}5}$ & **C**$_{8\text{-}10}$), MAP2 and pTau-positive dystrophic dendrites (white arrows in **C**$_{2,\,3,\,5}$ & **C**$_{7,\,8,\,10}$), and plaque-associated reactive astrocytosis (yellow arrows in **C**$_{4\text{-}5}$ & **C**$_{9\text{-}10}$)—in the same subfields of CA1-2 containing ApoER2 LA1-2 aggregates. Abbreviations: CA, cornu ammonis; ML, molecular layer; SP, stratum pyramidale; SR straum radiatum; SLM, stratum lacunosum-moleculare; NFL, neurofilament light chain; MAP2, microtubule associated protein-2; NEUN, neuronal nuclear antigen; GFAP, glial fibrillary acidic protein.

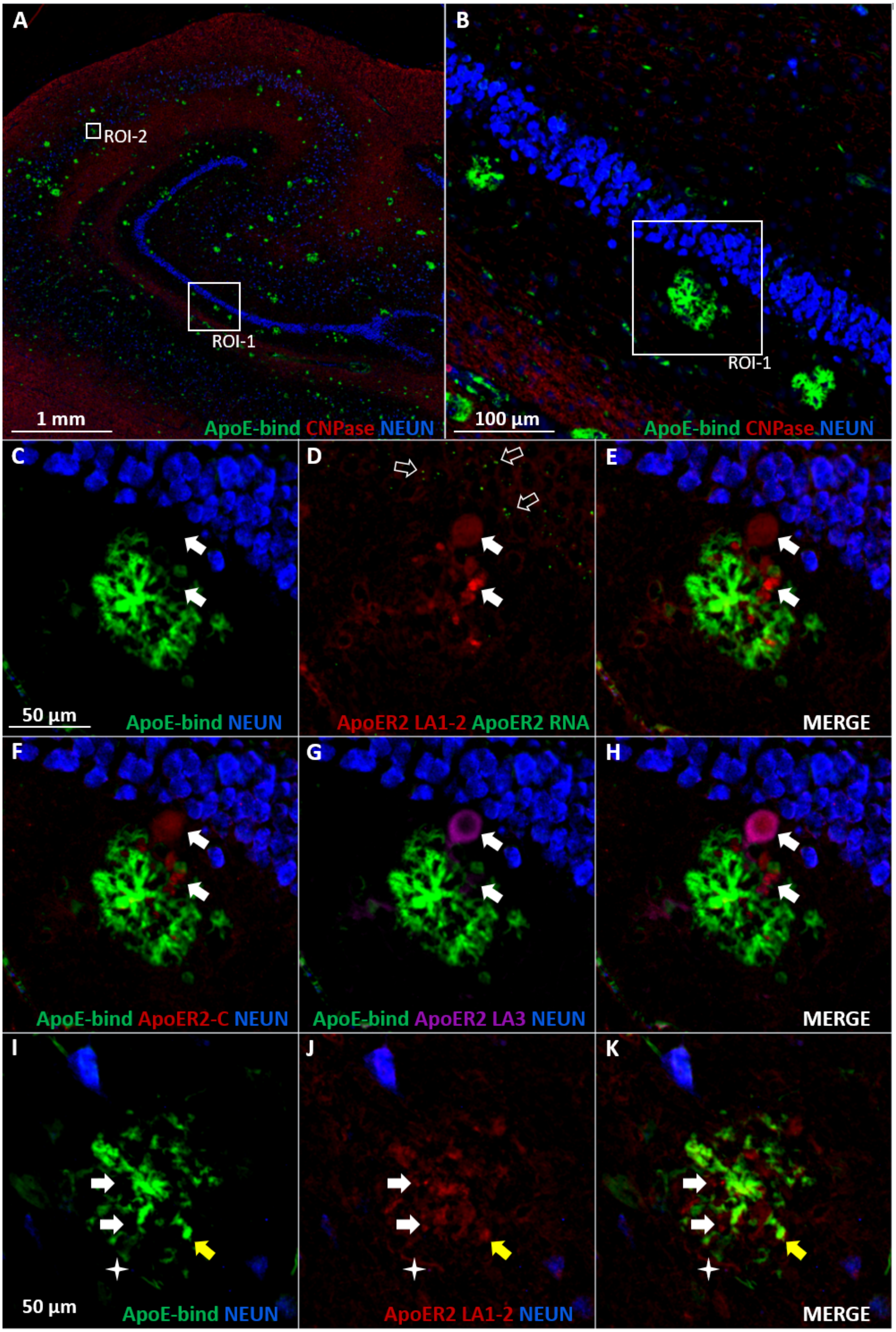
A
ROI-2
ROI-1
1 mm
ApoE-bind CNPase NEUN
B
ROI-1
100 µm
ApoE-bind CNPase NEUN
C
50 µm
ApoE-bind NEUN
D
ApoER2 LA1-2 ApoER2 RNA
E
MERGE
F
ApoE-bind ApoER2-C NEUN
G
ApoE-bind ApoER2 LA3 NEUN
H
MERGE
I
50 µm
ApoE-bind NEUN
J
ApoER2 LA1-2 NEUN
K
MERGE

**Extended Fig 5.2. Multi-epitope immunolabeling and ISH/RNA-protein codetection of ApoER2 in proximity to ApoE-enriched plaques in the molecular layer of dentate gyrus and hippocampus.** Panel **A** is a low magnification image of the hippocampus of a representative sporadic AD case (Braak Stage V, APOE3/3) that reveals extensive immunolabeling of plaques containing the receptor binding domain of ApoE (ApoE-bind, green [Panels **A-K**]). Panel **B** is an enlarged image containing three ApoE-enriched plaques within the molecular layer of the dentate gyrus. Panels **C-H** depict a subregion containing one of these plaques (ROI-1) selected to illustrate the spatial relations between ApoER2, ApoE, and dentate granule cells, which are labeled with NEUN shown in blue. Multidomain Anti-ApoER2 immunostaining revealed enrichment of several ApoER2 domains (white arrows, Panels **D-H**) in proximity to ApoE-enriched plaques, with particularly strong immunolabeling of the ApoER2 LA1-2 domain (red, Panels **D-E**, **J-K**). RNA (ISH)/protein (IHC) co-detection revealed ApoER2 mRNA expression (open arrows, Panel **D**) within adjacent dentate granule cells indicating a likely source for ApoER2 protein labeling. In the CA2 subregion (ROI-2, Panels **I-K**) anti-ApoER2 LA1-2 immunolabeling demonstrated similar accumulation of ApoER2 in what appears to be dystrophic neurites (star) in the vicinity of ApoE-enriched plaques. The close proximity and colocalization of ApoER2 and ApoE immunolabeling are depicted with white arrows and yellow arrows, respectively. **Abbreviations**: NEUN, neuronal nuclear/soma antigen; CNPase, Cyclic-nucleotide-phosphodiesterase.

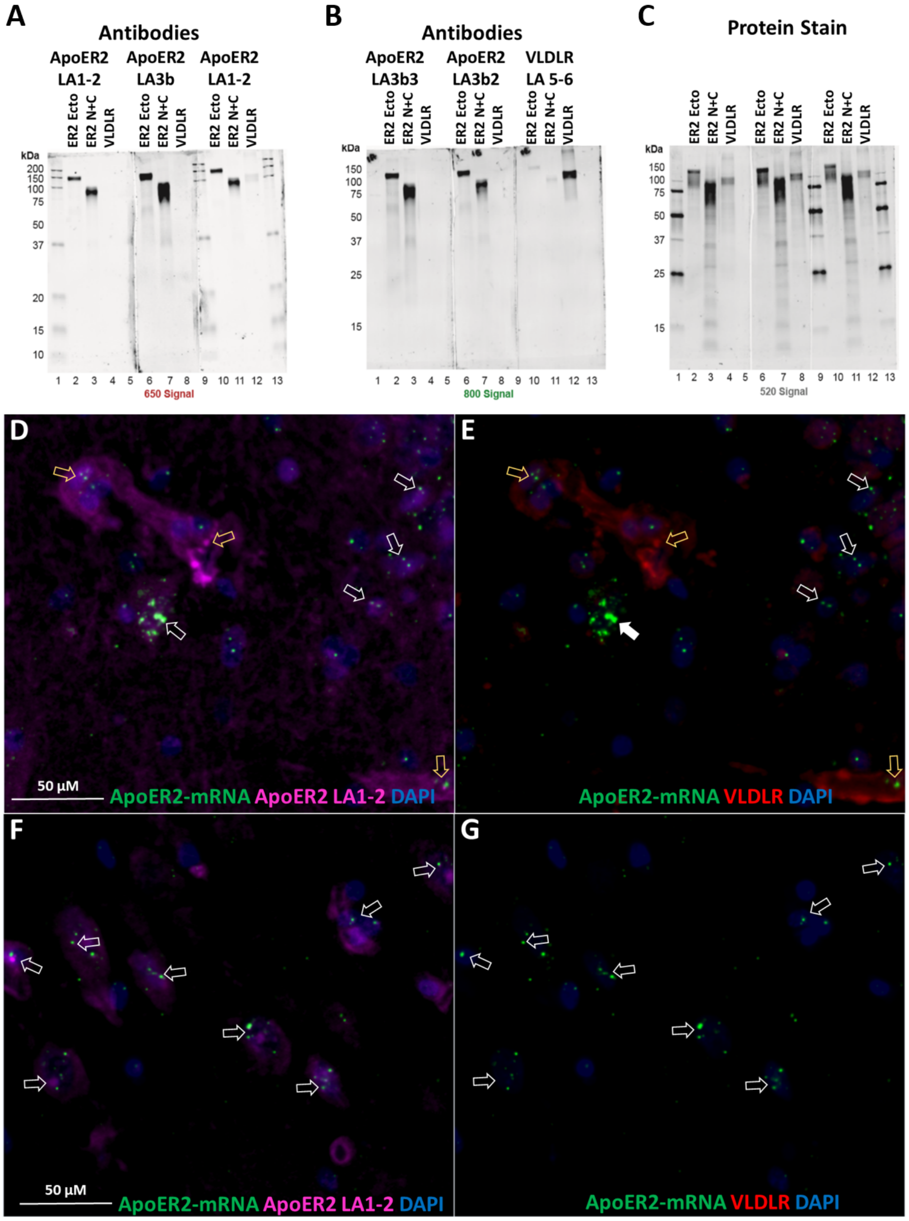


**Extended Fig 5.3. Selective labeling of ApoER2 via western blot and ApoER2 RNA-protein codetection assays.**

Recombinant proteins containing the ApoER2 ectodomain (ER2 Ecto), the ApoER2 N-terminus and part of the cytoplasmic tail (ER N+C), and the VLDL receptor (VLDLR) were loaded in triplicate at 250ng per lane. Antibody specificity for each respective protein was determined by fluorescence immunoblotting (IB) against ApoER2 (**A-B**), versus VLDLR (**B**). Panel **A** shows detection by ApoER2 LA1-2 specific-antibody (lanes 2-4, and 10-12), and ApoER2 LA3b-specific antibody (lanes 6-8). Panel **B** shows ApoER2 LA3b3 specificity (lanes 2-4), ApoER2 LA3b2 specificity (lanes 6-8) and VLDLR LA5-6 detection (lanes 10-12). Total protein amounts loaded per lane were determined by fluorescent protein staining (**C**). Fluorescence detection signals (650, 800, 520) are indicated as they correspond to each membrane. Western blots are representative of experiments performed three times. RNA-protein codetection assays in the human hippocampal formation showed cellular co-expression of ApoER2 mRNA and ApoER2 protein (white arrows) in dentate gyrus (**D**) and hippocampus (**F**). ApoER2 mRNA (**D-G**) and VLDLR protein (**E, G**) had little co-expression (yellow arrows). DAPI staining of nuclei is shown in blue.

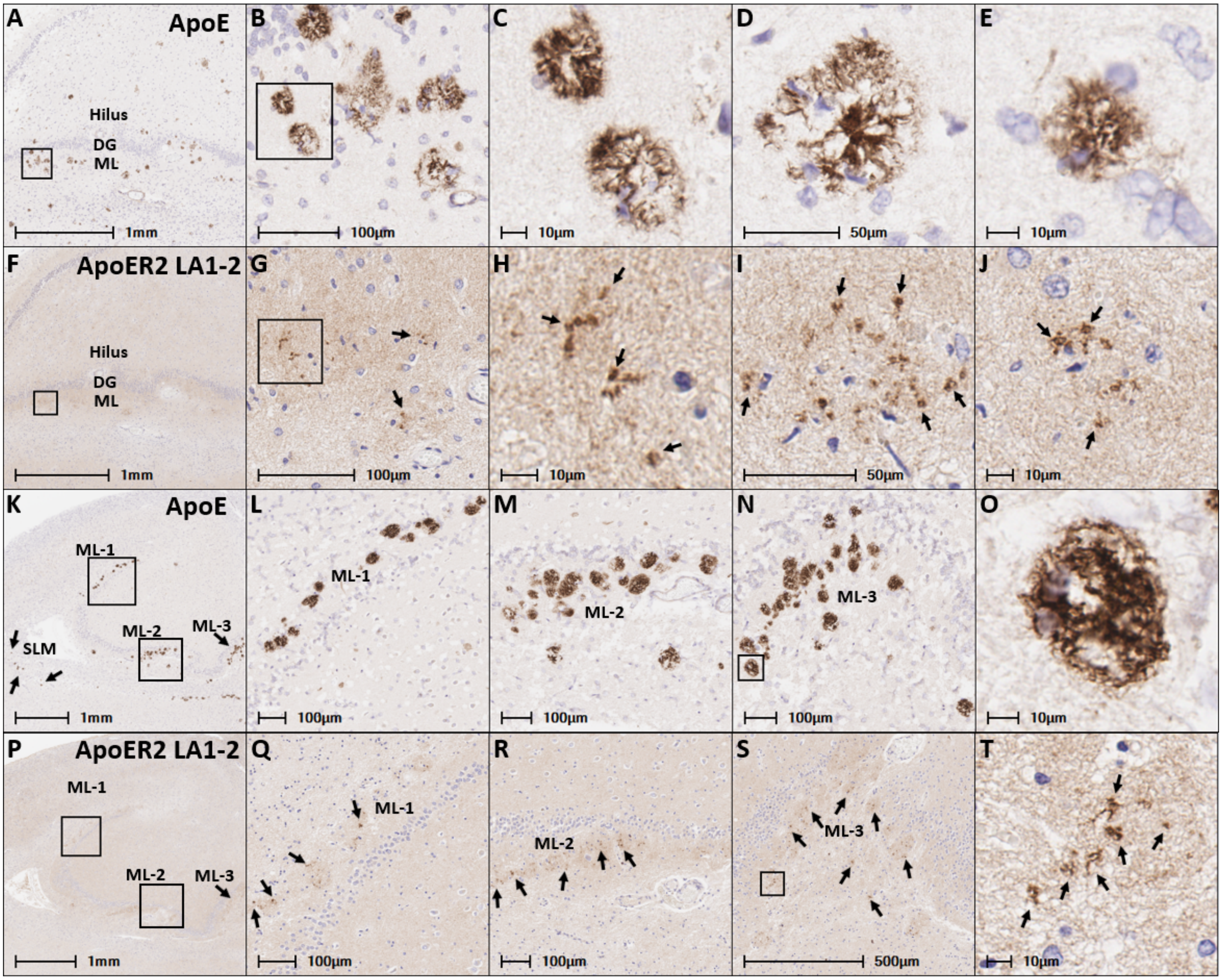


**Extended Fig 6.1. ApoE accumulates in the vicinity of its receptor ApoER2 in the perforant path terminal zones in sporadic AD.**

Coronal sections of the molecular layer of the dentate gyrus and hippocampus from the same sAD case (Braak stage V, APOE3/3) shown **Fig 6** are shown in **A-J**. Sections from another sAD case (Braak stage VI, APOE3/3), are shown in **K-T**. Serial sections stained with antibodies targeting ApoE (**A-E**, **K-O**) and ApoER2 LA1-2 (**F-J**, **P-T**) reveal prominent regional plaque-associated colocalization of ApoE and its receptor ApoER2 LA1-2 in the molecular layer of the dentate gyrus, with several ApoE-enriched plaques also observed in the SLM subregion of CA1 (arrows in **K**). Serial sections demonstrate that ApoER2 LA1-2 aggregates (arrows in **G-J**, **Q-T**) are in the immediate vicinity of, and appear to surround, ApoE-enriched plaques. **Abbreviations**: DG, dentate granule cell layer; ML, molecular layer; SLM, stratum lacunosum-moleculare.

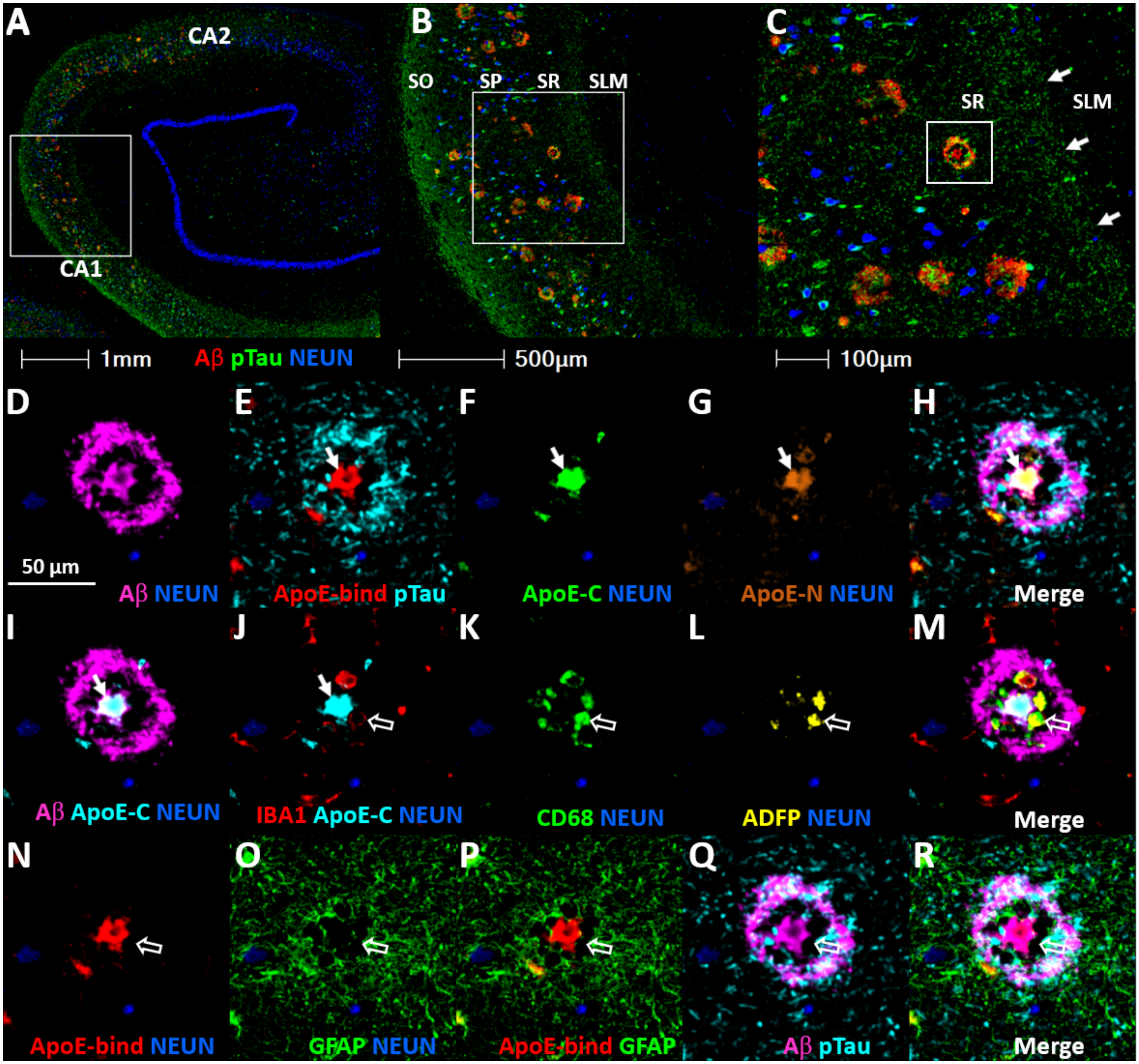


**Extended Fig 6.2. Close spatial relationships between ApoE, lipid-laden microglia and reactive astrocytes in the vicinity of neuritic plaques.**

MP-IHC was completed on a coronal section of the hippocampus from a representative (Braak stage V, APOE3/3) sAD case (**A-C**). The SO, SP, SR and SLM layers of CA1 are labeled in panel B, with the transition zone between the SR and SLM depicted by white arrows in panel **C**. A magnified image of one ApoE-enriched plaque in the SR subfield of CA1 is provided in **D-R**. MP-IHC staining confirmed that antibodies targeting ApoE-binding domain (**E, N, P**), ApoE C-terminus (**F, I**) and ApoE N-terminal domain (**G**) strongly label the central core of neuritic plaques (white arrows in **E-J**), and revealed close spatial relationships between ApoE and IBA1 (**J**), CD68 (**K**) and ADFP-positive (**L**), lipid-laden, infiltrating microglia (**J-M**) and GFAP-positive reactive astrocytes (**O-P, R**) in the neuritic plaque niche. **Abbreviations**: CA, cornu ammonis; SO, stratum oriens; SP, stratum pyramidale; SR straum radiatum; SLM, stratum lacunosum-moleculare; NEUN, neuronal nuclear antigen; IBA1, ionized calcium-binding adaptor molecule-1; CD68, cluster of differentiation 68; ADFP, adipophilin; GFAP, glial fibrillary acidic protein.

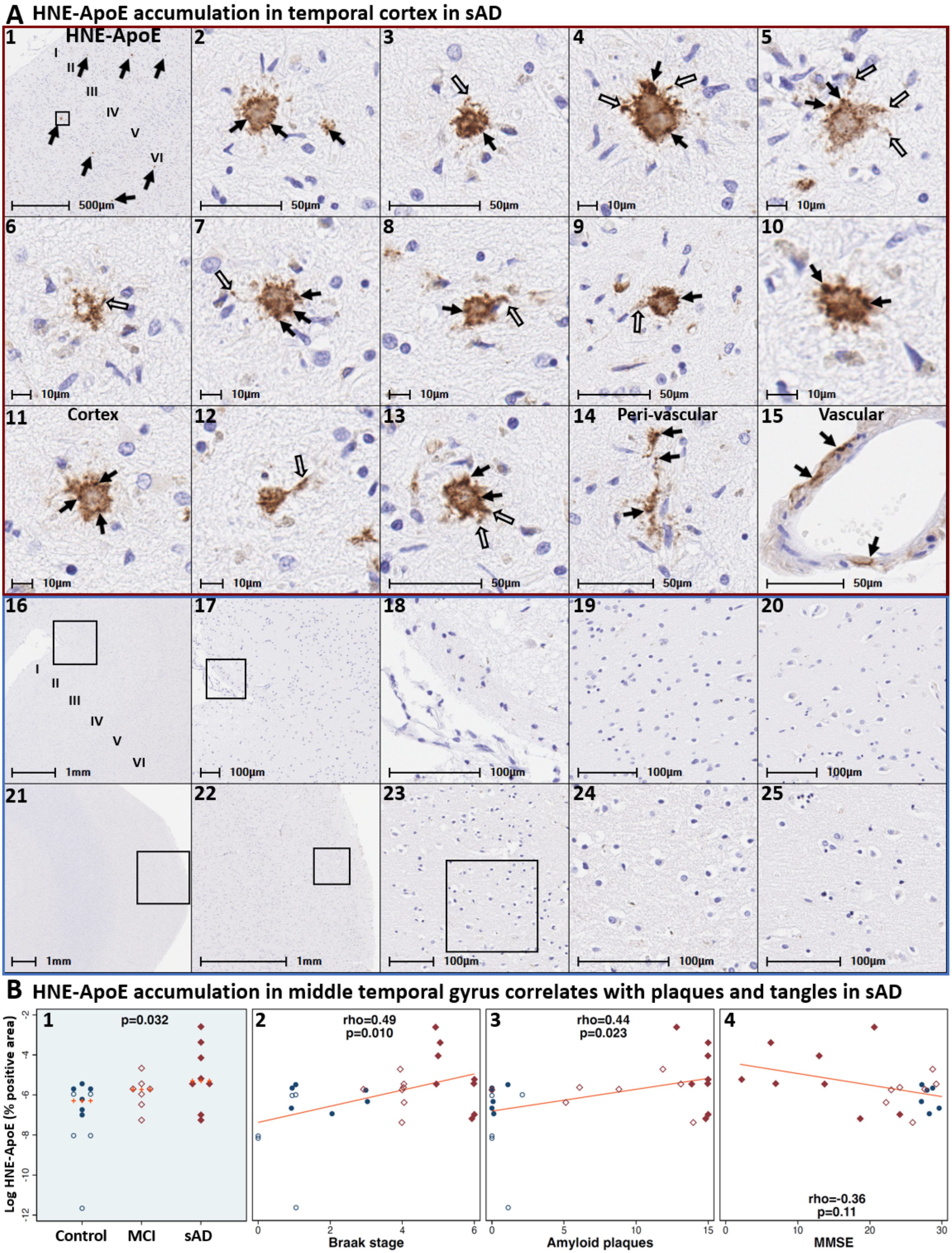
A HNE-ApoE accumulation in temporal cortex in sAD
HNE-ApoE
I
II
III
IV
V
VI
500µm
50µm
10µm
Cortex
Peri-vascular
Vascular
1mm
100µm
B HNE-ApoE accumulation in middle temporal gyrus correlates with plaques and tangles in sAD
p=0.032
Log HNE-ApoE (% positive area)
Control
MCI
sAD
rho=0.49
p=0.010
Braak stage
rho=0.44
p=0.023
Amyloid plaques
rho=-0.36
p=0.11
MMSE

**Extended Fig 7.1. HNE-ApoE accumulates in the temporal cortex in sporadic AD.**
Coronal sections of the temporal cortex from a representative (Braak stage V, APOE3/3) sporadic AD cases (**$A_{1-15}$**) and a non-AD control case (**$A_{16-25}$**) were stained with an anti-HNE-ApoE antibody. HNE-ApoE immunoreactive plaques were observed in superficial and deep cortical layers (**$A_{1-13}$**) in AD cases, but not in controls. HNE-ApoE immunolabeled granular structures near the plaque core (closed arrows in **$A_{1-13}$**) and elongated structures projecting outside of the plaque core (open arrows in **$A_{3-9}$**, **$A_{12-13}$**). HNE-ApoE expression was also observed in a subset of peri-vascular and vascular structures (closed arrows in **$A_{14-15}$**). HNE-ApoE accumulated in sAD cases and correlated with histochemical progression but not MMSE score (**B**).
Red and blue rectangles in Panel **A** indicate sAD and non-AD control cases, respectively.
**Panel B**: open and closed blue circles indicate young controls and age-matched controls; open and closed red diamonds indicate MCI cases and sAD cases, respectively.

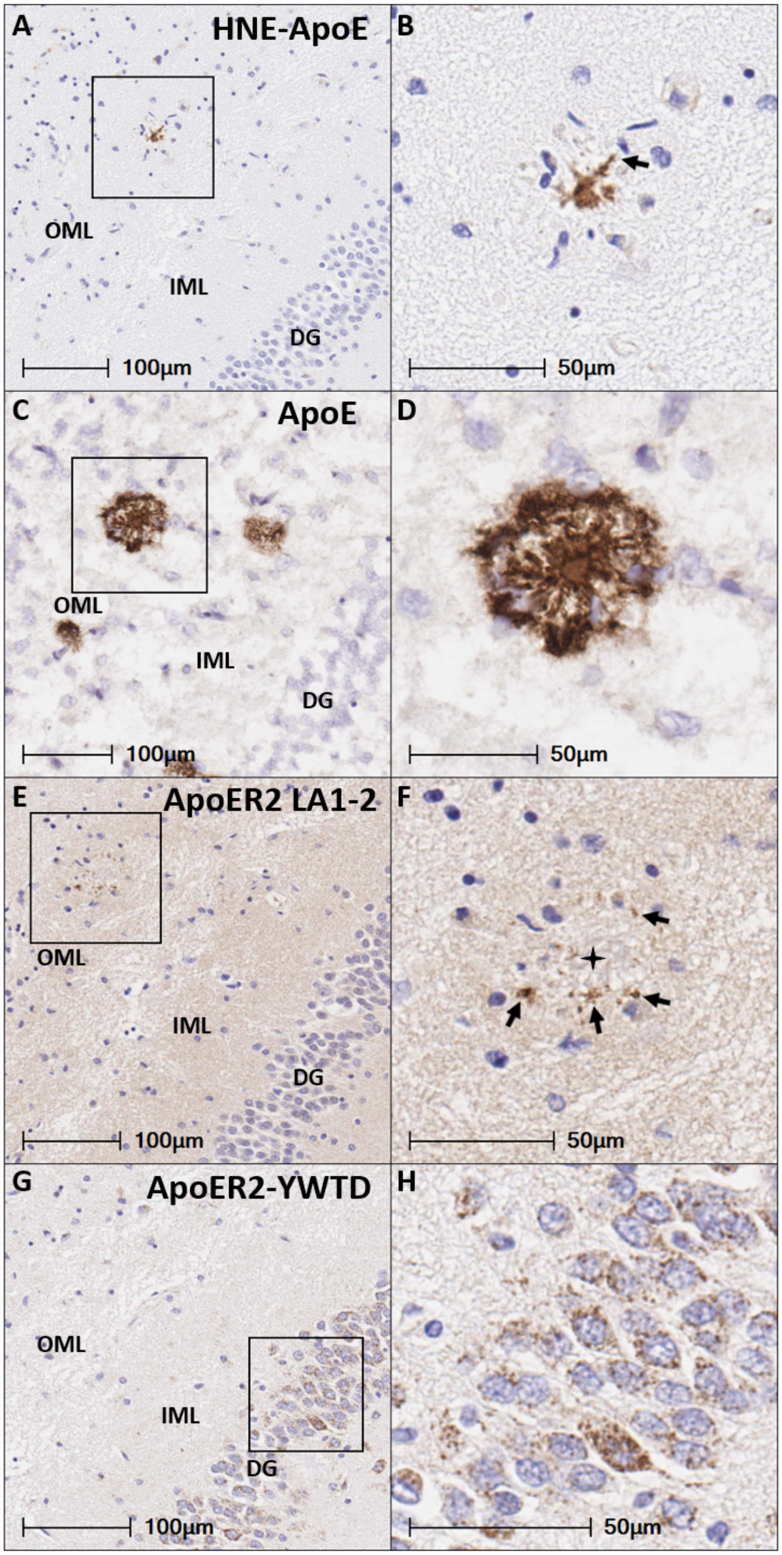


**Extended Fig 7.2. HNE-ApoE and native ApoE accumulate in the vicinity of ApoER2 LA 1-2 in the outer molecular layer of the dentate gyrus.** Serial coronal sections of the dentate granule cell layer and molecular layer from a representative (Braak stage V, APOE3/3) sAD case suggest partial, but incomplete colocalization of HNE-modified and native ApoE within plaques, with discrete ApoER2 LA1-2 aggregates surrounding plaques. The arrows in Panel B show depicts an HNE-ApoE-positive elongated structure projecting outside of the plaque core. The arrows in F depict discrete ApoER2 LA1-2 aggregates that surround the plaque core (depicted by star). **Abbreviations**: DG, dentate granule cell layer; OML, outer molecular layer; IML, inner molecular layer.

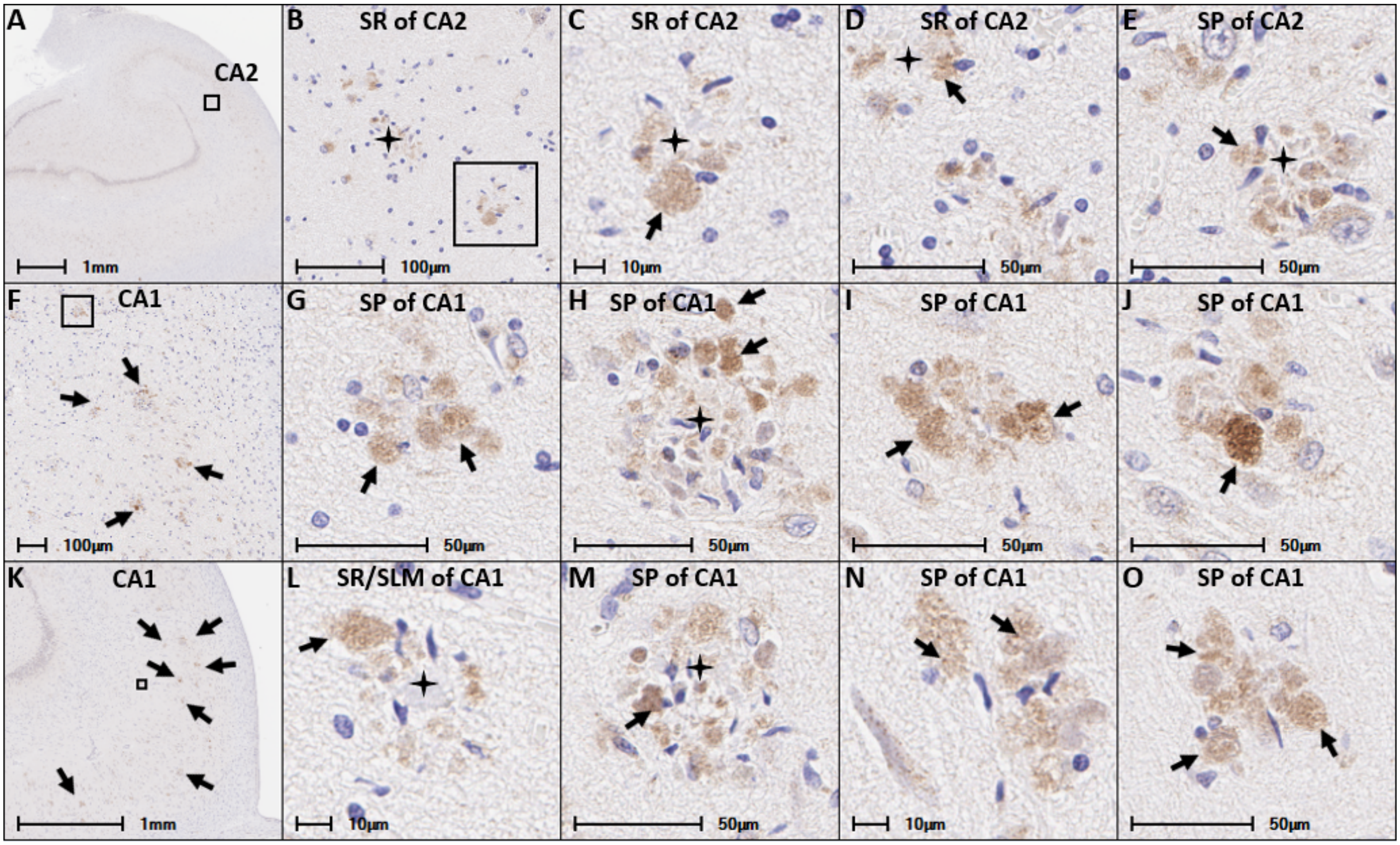


**Extended Fig 9.1. Dab1 plaque complexes accumulate in the cornu ammonis in sporadic AD**
Coronal sections of the hippocampus from a representative (Braak stage V, APOE3/3) sAD case (**A-O**) were stained with an anti-Dab1 antibody. Dab1-positive plaque complexes were observed in the SP and SR of the CA2 (**A-D**) and CA1 (**E-O**) subregions of the hippocampus. Arrows depict Dab1-positive globular structures that are consistent with swollen, dystrophic neurites in the vicinity of plaques (stars). Dab1-positive plaque complexes in **H** and **M** arrows have a diameter of approximately 50µm. **Abbreviations**: CA, cornu ammonis; SP, stratum pyramidale; SR, stratum radiatum; SLM, stratum lacunosum-moleculare.

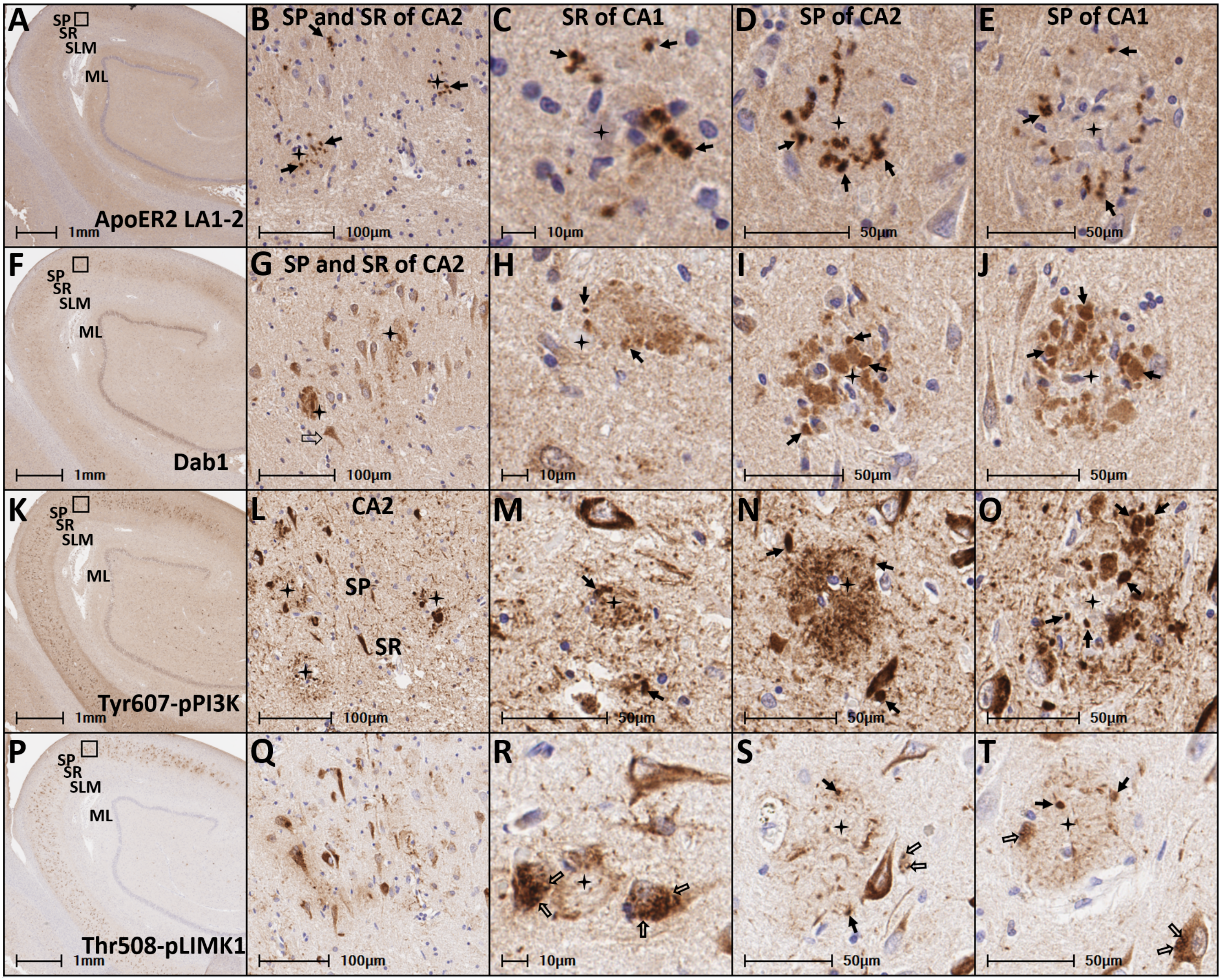


**Extended Fig 10.1. Accumulation of ApoER2 LA1-2, Dab1, Tyr607-pPI3K, Thr508-pLIMK1 in the CA2 subregion of the hippocampus in sAD**. Serial coronal sections through the hippocampus from the same (Braak stage V, APOE3/3) sAD case shown in **Fig 10** were stained with antibodies against the upstream pathway markers ApoER2 LA1-2 (**A-E**), and the ApoER2 adaptor protein Dab1 (**F-J**), and the downstream ApoER2-Dab1 signaling partners Tyr607-pPI3K (**K-O**) and Thr508-pLIMK1 (**P-T**). Plaques in the SP and SR subregions of CA2 are depicted by black stars in **A-T**. Neuritic plaque associated accumulations of ApoER2 LA1-2, Dab1, Tyr607-pPI3K and Thr508-pLIMK1 are depicted by arrows in **A-T**, **F-J**, **K-O**, and **P-T** respectively. Thr508-pLIMK1, which had comparatively weak expression within neuritic plaques, exhibited strong expression within granulovacuolar structures (open arrows in **R-T**) in a subset of neurons adjacent to plaques. **Abbreviations**: CA, cornu ammonis; SP, stratum pyramidale; SR, stratum radiatum; SLM, stratum lacunosum-moleculare.

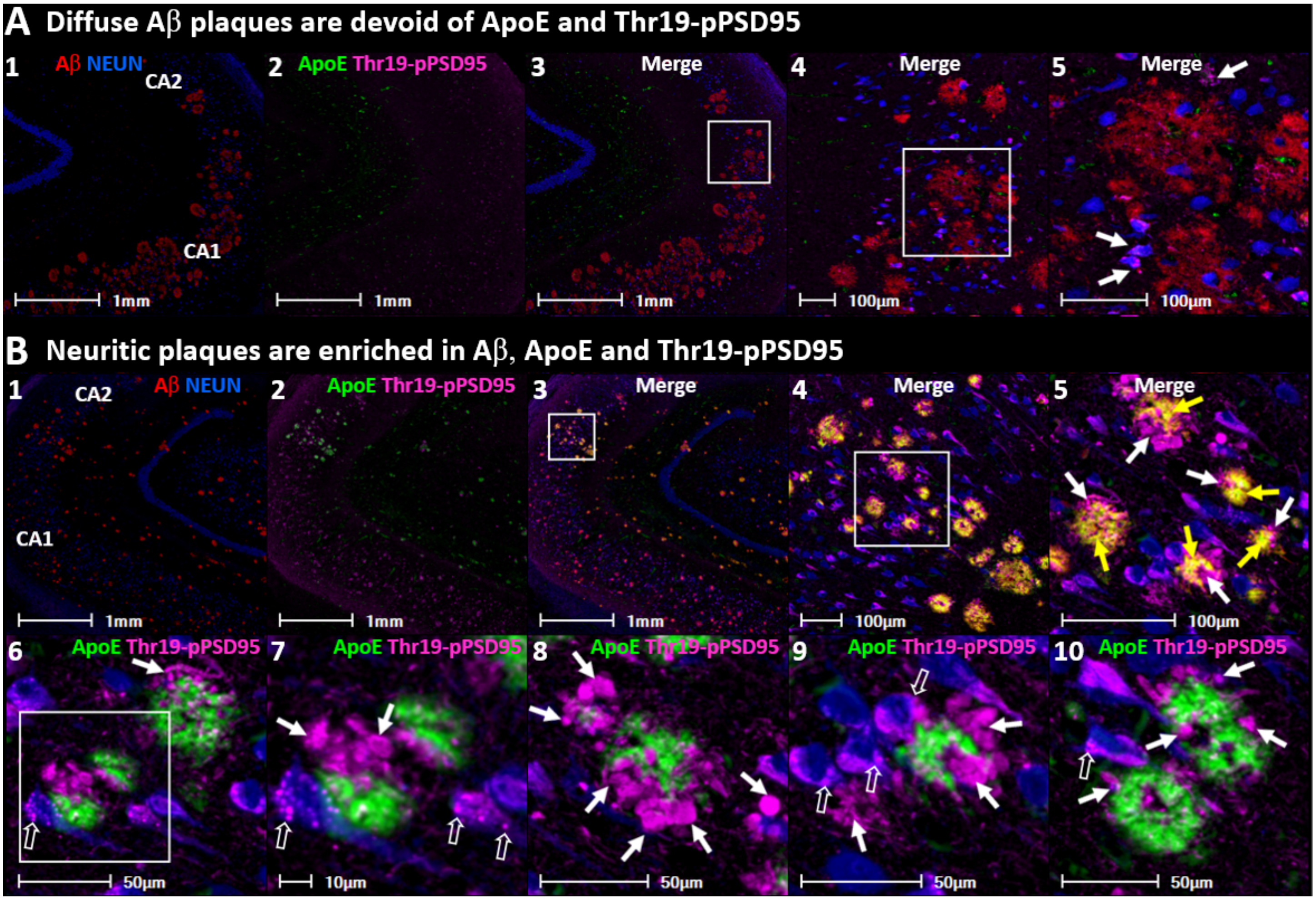


**Extended Fig 11.1 Diffuse plaques are enriched in Aβ but lack the ApoE and Thr19-pPSD95 that are abundant in neuritic plaques.** Unlike Aβ, virtually all ApoE-immunoreactive plaques are associated with dystrophic neurites (see Discussion in main text). Panels $\mathbf{A_{1\text{-}5}}$ show a coronal section of hippocampus from Mild Cognitive Impairment case (Braak Stage IV, ApoE 2/3) that had extensive diffuse Aβ plaques (total brain Aβ plaque load = 13.5 out of 15) but only mild cognitive deficits (MMSE 26). Panels $\mathbf{B_{1\text{-}10}}$ are a coronal section from a representative sAD case (Braak stage V, ApoE3/3) with extensive neuritic plaques (total brain Aβ plaque load = 15 out of 15) and major cognitive deficits (MMSE 6). Sections were stained with an antibody against Aβ (MOAB2, red), the ApoE C-terminal region (green), and Thr19-pPSD95 (cyan), with NEUN staining of neuronal nuclei shown in blue. Diffuse plaques in the MCI case had minimal expression of ApoE and Thr19-pPSD95 (white arrows in $\mathbf{A_5}$). By contrast, the neuritic plaques in the sAD case had high expression of ApoE and Aβ in the plaque core (yellow arrows in $\mathbf{B_5}$), and extensive accumulation of Thr19-pPSD95 in dystrophic neurons (closed white arrows in $\mathbf{B_{5\text{-}10}}$) and surrounding abnormal neurons (open white arrows in $\mathbf{B_{6\text{-}10}}$) in the vicinity of ApoE-enriched plaques.

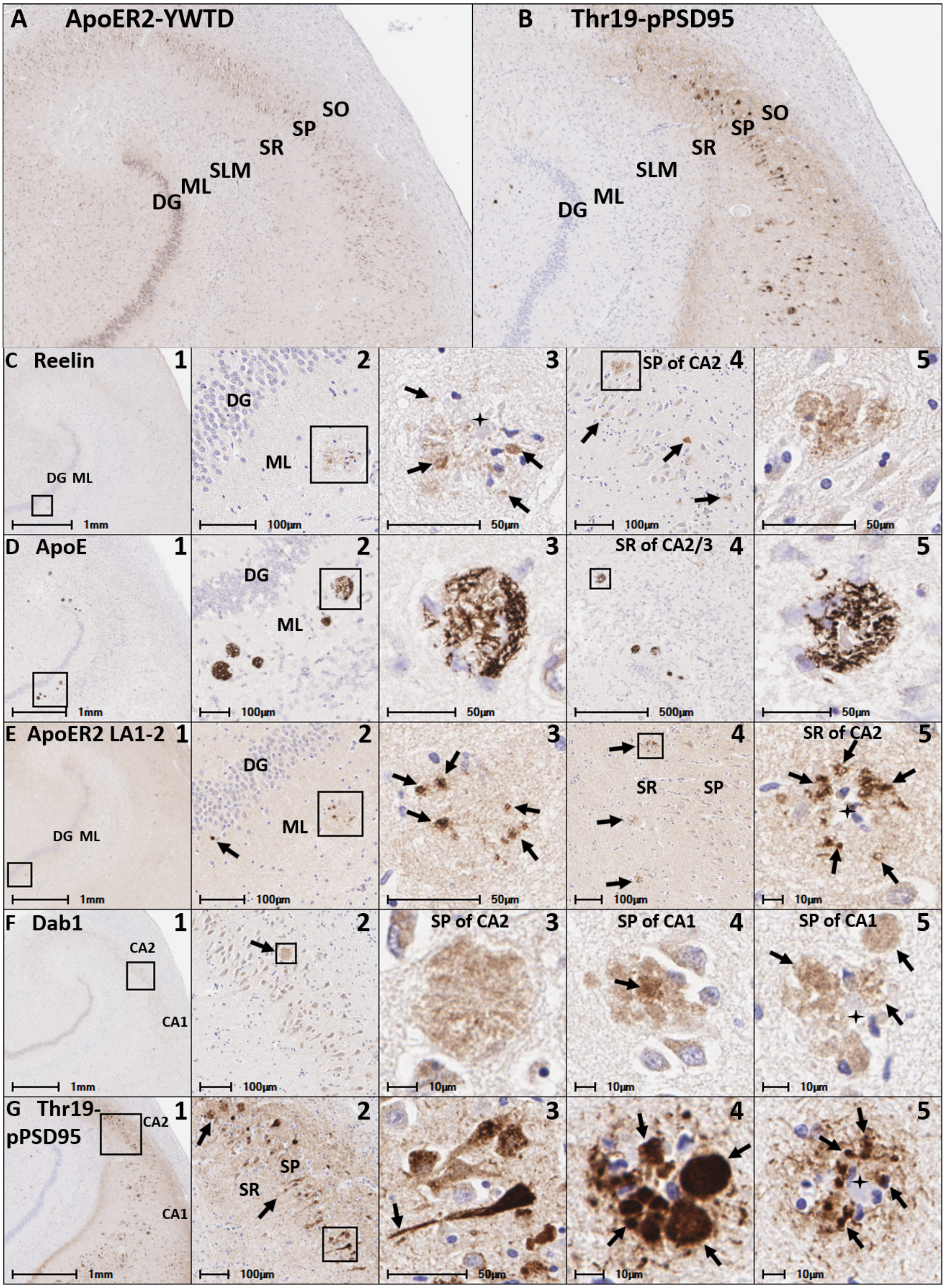
A ApoER2-YWTD
SO
SP
SR
SLM
ML
DG
B Thr19-pPSD95
SO
SP
SR
SLM
ML
DG
C Reelin
1
DG ML
1mm
2
DG
ML
100µm
3
50µm
4
SP of CA2
100µm
5
50µm
D ApoE
1
1mm
2
DG
ML
100µm
3
50µm
4
SR of CA2/3
500µm
5
50µm
E ApoER2 LA1-2
1
DG ML
1mm
2
DG
ML
100µm
3
50µm
4
SR
SP
100µm
5
SR of CA2
10µm
F Dab1
1
CA2
CA1
1mm
2
100µm
3
SP of CA2
10µm
4
SP of CA1
10µm
5
SP of CA1
10µm
G Thr19-pPSD95
1
CA2
CA1
1mm
2
SP
SR
100µm
3
50µm
4
10µm
5
10µm

**Extended Fig 13.1. IHC evidence for convergence of ApoE/Reelin-ApoER2-Thr19-pPSD95 axis pathologies in the perforant path target zones in early sAD.**
Single-marker IHC was used to label serial coronal sections of the perforant path target zone from the same early sporadic AD case (MMSE 24, Braak stage V, APOE3/3) shown in **Fig 13**. Panel **A** shows ApoER2 expression within dentate granule cells and pyramidal neurons in CA1-3. Panel **B** shows expression of Thr19-pPSD95 (marker of synaptic disassembly) in neuritic plaques and abnormal neurons in CA1-2, and their dendritic projections emanating into the SR subfield. Panels $\mathbf{C}_{1\text{-}5}$ reveal plaque associated Reelin accumulation in the molecular layer of the dentate gyrus ($\mathbf{C}_{1\text{-}3}$, arrows) and the SP layer of CA2 ($\mathbf{C}_{4\text{-}5}$, arrows). Panels $\mathbf{D}_{1\text{-}5}$ reveal plaque associated ApoE accumulation in the molecular layer of the dentate gyrus ($\mathbf{D}_{1\text{-}3}$) and the SLM subfield of CA2 ($\mathbf{D}_{4\text{-}5}$). Panels $\mathbf{E}_{1\text{-}5}$ reveal plaque associated ApoER2 LA1-2 aggregates in the molecular layer of the dentate gyrus ($\mathbf{E}_{1\text{-}3}$, arrows) and the SR and SP subfields of CA1-2 ($\mathbf{E}_{4\text{-}5}$, arrows)
Panels $\mathbf{F}_{1\text{-}5}$ reveal plaque associated Dab1 accumulation that is most pronounced in the SP and SR subregions of CA2 and CA1 ($\mathbf{F}_{1\text{-}5}$, arrows). Panels $\mathbf{G}_{1\text{-}5}$ reveal strong Thr19-pPSD95 expression in neuritic plaques, affected neurons and their projections in CA1 and CA2 ($\mathbf{G}_{1\text{-}3}$). Panels $\mathbf{G}_{4\text{-}5}$ show strong expression in Thr19-pPSD95 in neuritic plaque complexes within SR and SLM of CA1.
**Abbreviations**: DG, dentate granule cells; ML, molecular layer; CA, cornu ammonis; SP, stratum pyramidale; SR, stratum radiatum; SLM, stratum lacunosum-moleculare.

### II. Supplementary Tables

**Table S1. The ApoE-ApoE receptor peroxidation cascade vs. the amyloid cascade hypothesis: Implications for therapeutics & prevention**

**Table S2. Twenty-nine cases spanning the clinicopathological spectrum of Alzheimer's disease progression**

**Table S3. Demographic, neuropathological, and cognitive characteristics of twelve cases spanning the clinicopathological spectrum of Alzheimer's disease progression (pLIMK1 & pPI3K)**

**Table S4. Demographic, neuropathological, and cognitive characteristics of twelve cases spanning the clinicopathological spectrum of Alzheimer's disease progression (pPSD95)**

**Table S5. Key Resources**

**Table S1. The ApoE-ApoE receptor peroxidation cascade vs. the amyloid cascade hypothesis: Implications for therapeutics & prevention**

| | Amyloid Cascade | ApoE-ApoER2 Peroxidation Cascade |
|---|---|---|
| **Pathogenic factors and events** | **↑ synthesis & secretion of Aβ** | **ApoE & ApoER2 peroxidation, adduction & crosslinking → ApoE/Reelin-ApoER2 axis disruption** |
| **Therapeutic approaches** | **Anticipated effects** | **Anticipated effects** |
| **Aβ monomer removal** [a] | **Protective**<br>↓ Aβ induced neurotoxicity | **Counterproductive** [a]<br>↓ neutralization and clearance of PxApoE<br>↑ PxApoE induced toxicity |
| **Aβ oligomer removal** [b] | **Protective**<br>↓ Aβ induced neurotoxicity | **Protective** (progression only) [b]<br>↓ Aβ induced neurotoxicity |
| **BACE1 inhibition** [c] | **Protective**<br>↓ Aβ induced neurotoxicity | **Counterproductive** [c]<br>↓ neutralization and clearance of PxApoE<br>↑ PxApoE induced toxicity |
| **Gamma secretase inhibition** [d] | **Protective**<br>↓ Aβ induced neurotoxicity | **Counterproductive** [d]<br>↓ neutralization and clearance of PxApoE<br>↑ PxApoE induced toxicity |
| **Selective PxApoE removal** [e] | not applicable | **Protective** [e]<br>↓ PxApoE induced toxicity |
| **Inhibition of ApoE peroxidation** [f] | not applicable | **Protective** [f]<br>↓ PxApoE induced toxicity |
| **Preventive approaches** | | |
| **Mediterranean & high flavonoid diets** [g] | no clear mechanism | ↓ PxApoE induced toxicity [g] |
| **Air pollution mitigation** [h] | no clear mechanism | ↓ PxApoE induced toxicity [h] |
| **Avoidance of excess Fe, Cu, Al** [i] | no clear mechanism | ↓ Lipid peroxidation & PxApoE induced toxicity [i] |
| **Alcohol moderation** [j] | no clear mechanism | ↓ PxApoE induced toxicity [j] (acetaldehyde variant) |
| **HSV prophylaxis** [k] | may decrease Aβ production [k] | ↓ PxApoE induced toxicity [k] |
| **Other observations** | | **Link to the ApoE-ApoER2 Peroxidation Cascade** |
| **Iron accumulation in AD brains** [l] | no clear explanation | Iron is a major catalyst for brain lipid peroxidation [l] |
| **Aging is strongest risk factor for AD** [m] | no clear explanation | Older adults have markedly increased lipid peroxidation [m] |
| **Selective vulnerability of entorhinal-hippocampal structures** | no clear explanation | High ApoER2 expression & constant demand for ApoE/Reelin-ApoE receptor pathway activation |

This table compares and contrasts the proposed pathogenic factors and mechanisms underpinning the Amyloid cascade and the PxApoE-ApoE receptor cascade hypotheses of sporadic AD. Therapeutic approaches targeting these two hypotheses are predicted to have different, and sometimes opposing, effects on disease progression, as summarized in notes [a-m] below.

[a] Aβ1-40 and Aβ1-42 monomers exhibit potent antioxidant effects at physiological concentrations.[1-4] Aβ is localized to lipoproteins and appears to protect vulnerable polyunsaturated cargo from peroxidation.[1, 5, 6] According to the PxApoE-ApoE receptor cascade hypothesis, Aβ monomers serve a protective function by binding, neutralizing, and targeting toxic, lipid peroxidation-modified ApoE particles to glial-mediated clearance pathways. Thus, monoclonal antibodies that selectively remove Aβ monomers, and medications that inhibit Aβ1-40 and Aβ1-42 synthesis (see BACE1 & gamma secretase inhibitors below), are proposed to exacerbate PxApoE-induced toxicity and to accelerate cognitive decline.

[b] According to the Amyloid cascade hypothesis, Aβ is the primary causal factor underlying sporadic AD. By contrast, according to the PxApoE-ApoE receptor cascade hypothesis, (1) Aβ monomer neurosecretion is a secondary, protective response to oxidative and cellular stress, and (2) lipid peroxidation-induced disruption of ApoE receptors leads to extracellular trapping of ApoE and PxApoE, where they provide a seed for Aβ oligomerization. With excessive or prolonged PxApoE exposure, these protective effects of Aβ monomers are proposed to transition to the well-established neurotoxic effects of high concentrations of oligomeric Aβ, overwhelming glial-mediated Aβ clearance pathways and leading to extracellular Aβ deposition. According to the PxApoE-ApoE receptor cascade hypothesis, oligomeric Aβ is an exacerbating factor in AD pathogenesis. Thus, selective removal of oligomeric Aβ is anticipated to produce substantial (but limited) benefit in slowing cognitive decline.

[c] BACE1 is the rate-limiting enzyme required for conversion of AβPP to Aβ monomers. According to the Amyloid cascade hypothesis, BACE1 inhibition is predicted to slow AD progression. By contrast, according to the PxApoE-ApoE receptor cascade hypothesis, BACE1 inhibition is predicted to accelerate cognitive decline by impairing Aβ-mediated neutralization and clearance of PxApoE, thereby enhancing PxApoE induced toxicity. Findings from recent controlled trials in which BACE1 inhibitors paradoxically worsened cognitive decline,[7, 8] despite markedly decreasing CSF Aβ1-40 and Aβ1-42,[9] are consistent with the PxApoE-ApoE receptor cascade hypothesis.

[d] Gamma secretase is another enzyme required for conversion of AβPP to Aβ monomers whose inhibition is classically predicted to slow AD progression. According to the PxApoE-ApoE receptor cascade hypothesis, gamma secretase inhibition is predicted to accelerate cognitive decline by impairing Aβ-mediated neutralization and clearance of PxApoE. In controlled trials gamma secretase inhibitors lacked clinical efficacy,[10-12] and in some cases paradoxically worsened cognitive decline.[8]

[e & f] According to the PxApoE-ApoE receptor cascade hypothesis, selective removal of PxApoE and inhibition of ApoE particle peroxidation are therapeutic strategies that are anticipated to decrease

PxApoE-induced toxicity and prevent AD progression (see main paper **Fig. 3**). Neither strategy is directly relevant for the Amyloid cascade hypothesis.

[g] Findings from observational studies suggest that Mediterranean-type diets slow cognitive decline with aging.[13-18] Several features of industrialized diets enhance generation aldehydic products of lipid peroxidation that are implicated in the PxApoE-ApoE receptor cascade. Notable examples include widespread use of oxidizable oils and low consumption of plant foods that are rich in lipophilic and amphiphilic polyphenols (i.e., flavonoids). The Mediterranean diet, which uses predominantly olive oil and is enriched in flavonoids,[19] could plausibly decrease ApoE particle peroxidation, PxApoE-induced toxicity and prevent AD progression.

[h] Long-term exposure to air pollution is a risk factor for AD in observational studies [20-22] and induces AD type pathologies in experimental models.[23] Automobile exhaust and other air pollution are major sources of carbonyl compounds that are similar (and in some cases overlapping) with aldehydic products of lipid peroxidation.[24-27] In the present studies we observed that 4-ONE, 4-HNE, CRA, MDA and acetaldehyde react with Lys and His residues enriched within the binding regions of ApoE, Reelin and ApoE receptors. Carbonyl compounds in air pollution could plausibly PxApoE induced toxicities and ApoE/Reelin-ApoE receptor disruptions in a similar manner.

[i] Metals such as iron (Fe), copper (Cu), and aluminum (Al) are potent inducers of lipid peroxidation and the production of reactive lipid aldehydes.[28-32] According to the PxApoE-ApoE receptor cascade hypothesis, avoidance (or removal) of metals that promote ApoE particle peroxidation are plausible preventive and therapeutic strategies that are anticipated to decrease PxApoE-induced toxicity and prevent AD progression. Neither strategy is directly relevant for the Amyloid cascade hypothesis.

[j] Alcohol use disorders are a major risk factor for all types of dementia (including AD), and especially early-onset dementia.[33] In experimental models, heavy alcohol use induces generalized lipid peroxidation.[34-37] Acetaldehyde—an aldehydic product of lipid peroxidation that readily reacts with Lys and His residues—is also a major metabolic byproduct of alcohol.[38] According to the PxApoE-ApoE receptor cascade, decreasing heavy alcohol use could plausibly decrease ApoE particle peroxidation and acetaldehyde-ApoE-induced toxicity, thereby slowing AD progression.

[k] Herpes simplex virus type 1 (HSV-1) is a risk factor for sporadic AD in observational studies.[39, 40] HSV-1 can traverse into the hippocampal region in humans.[41] ApoE is reported to be involved in the HSV-1 life cycle.[42] In *in vitro* studies, HSV-1 is a potent inducer of neuronal lipid peroxidation.[43] Thus, HSV-1 prophylaxis could plausibly decrease ApoE peroxidation, PxApoE-induced toxicity and AD progression.

[l] Brain Fe accumulation is associated with AD progression, neurodegeneration, and cognitive decline with aging.[44-48]

[m] Aging is associated with marked increases in lipid peroxidation products.[49]

**Abbreviations**: ApoE, apolipoprotein E; Px, peroxidation; PxApoE, lipid peroxidation modified ApoE particles; Aβ, amyloid beta; AβPP, amyloid beta precursor protein.

[12] Coric V, van Dyck CH, Salloway S, Andreasen N, Brody M, Richter RW, Soininen H, Thein S, Shiovitz T, Pilcher G, Colby S, Rollin L, Dockens R, Pachai C, Portelius E, Andreasson U, Blennow K, Soares H, Albright

C, Feldman HH, Berman RM (2012) Safety and tolerability of the gamma-secretase inhibitor avagacestat in a phase 2 study of mild to moderate Alzheimer disease. *Arch Neurol* **69**, 1430-1440.
[13] Petersson SD, Philippou E (2016) Mediterranean Diet, Cognitive Function, and Dementia: A Systematic Review of the Evidence. *Adv Nutr* **7**, 889-904.
[14] Samuelsson J, Kern S, Zetterberg H, Blennow K, Rothenberg E, Wallengren O, Skoog I, Zettergren A (2021) A Western-style dietary pattern is associated with cerebrospinal fluid biomarker levels for preclinical Alzheimer's disease-A population-based cross-sectional study among 70-year-olds. *Alzheimers Dement (N Y)* **7**, e12183.
[15] Ballarini T, Melo van Lent D, Brunner J, Schroder A, Wolfsgruber S, Altenstein S, Brosseron F, Buerger K, Dechent P, Dobisch L, Duzel E, Ertl-Wagner B, Fliessbach K, Freiesleben SD, Frommann I, Glanz W, Hauser D, Haynes JD, Heneka MT, Janowitz D, Kilimann I, Laske C, Maier F, Metzger CD, Munk M, Perneczky R, Peters O, Priller J, Ramirez A, Rauchmann B, Roy N, Scheffler K, Schneider A, Spottke A, Spruth EJ, Teipel SJ, Vukovich R, Wiltfang J, Jessen F, Wagner M, group Ds (2021) Mediterranean Diet, Alzheimer Disease Biomarkers and Brain Atrophy in Old Age. *Neurology*.
[16] Charisis S, Ntanasi E, Yannakoulia M, Anastasiou CA, Kosmidis MH, Dardiotis E, Hadjigeorgiou G, Sakka P, Scarmeas N (2021) Mediterranean diet and risk for dementia and cognitive decline in a Mediterranean population. *J Am Geriatr Soc*.
[17] Andreu-Reinon ME, Chirlaque MD, Gavrila D, Amiano P, Mar J, Tainta M, Ardanaz E, Larumbe R, Colorado-Yohar SM, Navarro-Mateu F, Navarro C, Huerta JM (2021) Mediterranean Diet and Risk of Dementia and Alzheimer's Disease in the EPIC-Spain Dementia Cohort Study. *Nutrients* **13**.
[18] Agarwal P, Dhana K, Barnes LL, Holland TM, Zhang Y, Evans DA, Morris MC (2021) Unhealthy foods may attenuate the beneficial relation of a Mediterranean diet to cognitive decline. *Alzheimers Dement*.
[19] Masella R, Giovannini C, Vari R, Di Benedetto R, Coni E, Volpe R, Fraone N, Bucci A (2001) Effects of dietary virgin olive oil phenols on low density lipoprotein oxidation in hyperlipidemic patients. *Lipids* **36**, 1195-1202.
[20] Lin FC, Chen CY, Lin CW, Wu MT, Chen HY, Huang P (2021) Air Pollution Is Associated with Cognitive Deterioration of Alzheimer's Disease. *Gerontology*, 1-9.
[21] Ran J, Schooling CM, Han L, Sun S, Zhao S, Zhang X, Chan KP, Guo F, Lee RS, Qiu Y, Tian L (2021) Long-term exposure to fine particulate matter and dementia incidence: A cohort study in Hong Kong. *Environ Pollut* **271**, 116303.
[22] Mortamais M, Gutierrez LA, de Hoogh K, Chen J, Vienneau D, Carriere I, Letellier N, Helmer C, Gabelle A, Mura T, Sunyer J, Benmarhnia T, Jacquemin B, Berr C (2021) Long-term exposure to ambient air pollution and risk of dementia: Results of the prospective Three-City Study. *Environ Int* **148**, 106376.
[23] Patten KT, Valenzuela AE, Wallis C, Berg EL, Silverman JL, Bein KJ, Wexler AS, Lein PJ (2021) The Effects of Chronic Exposure to Ambient Traffic-Related Air Pollution on Alzheimer's Disease Phenotypes in Wildtype and Genetically Predisposed Male and Female Rats. *Environ Health Perspect* **129**, 57005.
[24] Liu W, Zhang J, Kwon J, Weisel C, Turpin B, Zhang L, Korn L, Morandi M, Stock T, Colome S (2006) Concentrations and source characteristics of airborne carbonyl compounds measured outside urban residences. *J Air Waste Manag Assoc* **56**, 1196-1204.
[25] Grosjean D, Grosjean E (2002) Airborne carbonyls from motor vehicle emissions in two highway tunnels. *Res Rep Health Eff Inst*, 57-78; discussion 79-92.
[26] Kean AJ, Grosjean E, Grosjean D, Harley RA (2001) On-road measurement of carbonyls in California light-duty vehicle emissions. *Environ Sci Technol* **35**, 4198-4204.
[27] Costa LG, Cole TB, Coburn J, Chang YC, Dao K, Roque PJ (2017) Neurotoxicity of traffic-related air pollution. *Neurotoxicology* **59**, 133-139.
[28] Smith MA, Perry G (1995) Free radical damage, iron, and Alzheimer's disease. *J Neurol Sci* **134 Suppl**, 92-94.

### Table S2. Twenty-nine cases spanning the clinicopathological spectrum of Alzheimer’s disease progression

| ID | Expired age | Gender | Post mortem interval (hours) | Braak stage (0-6) | MMSE (0-30) | Total plaques (0-15) [a] | Total tangles (0-15) [a] | Neuritic plaque density (0-3) | ApoE status | Single-marker IHC (n=26) [b] | pPI3K, pLIMK1, pPSD95 (n=18) [c] |
|---|---|---|---|---|---|---|---|---|---|---|---|
| **Alzheimer's Disease** | | | | | | | | | | | |
| 1 | 85-89 | Female | 3.42 | Stage VI | 2 | 15 | 15 | 3, frequent | 3/3 | X | X |
| 2 | 70-74 | Male | 4.83 | Stage VI | 7 | 15 | 15 | 3, frequent | 3/4 | X | |
| 3 | 80-84 | Female | 4 | Stage VI | 19 | 15 | 15 | 3, frequent | 3/3 | X | X |
| 4 | 80-84 | Female | 3.33 | Stage VI | 24 | 15 | 12 | 3, frequent | 3/3 | X | |
| 5 | 75-79 | Female | 3.08 | Stage V | 6 | 15 | 15 | 3, frequent | 3/3 | X | X |
| 6 | 90+ | Female | 2.17 | Stage V | 13 | 15 | 11 | 3, frequent | 3/3 | X | X |
| 7 | 80-84 | Male | 4 | Stage V | 14 | 14 | 13 | 3, frequent | 3/4 | X | X |
| 8 | 85-89 | Male | 2.2 | Stage V | 21 | 13 | 11 | 3, frequent | 2/3 | X | |
| **Mild Cognitive Impairment** | | | | | | | | | | | |
| 9 | 90+ | Female | 3.16 | Stage IV | 22 | 5 | 9 | 3, frequent | 3/3 | X | X |
| 10 | 85-89 | Male | 4.17 | Stage IV | 23 | 0 | 7 | 0, none | 3/3 | X | |
| 11 | 75-79 | Female | 3.17 | Stage IV | 24 | 6 | 9 | 0, none | 2/3 | X | X |
| 12 | 90+ | Female | 3.16 | Stage IV | 26 | 14 | 8 | 3, frequent | 2/3 | X | X |
| 13 | 80-84 | Female | 3 | Stage IV | 29 | 13 | 7 | 3, frequent | 2/3 | X | |
| 14 | 80-84 | Female | 3.17 | Stage IV | 29 | 12 | 9 | 3, frequent | 2/3 | X | |
| 15 | 85-89 | Male | 2.25 | Stage III | 28 | 9 | 6 | 2, moderate | 3/3 | X | X |
| 16 | 90+ | Male | 3 | Stage III | 28 | 1 | 4 | 1, sparse | 3/3 | | X |
| **Age-matched Controls** | | | | | | | | | | | |
| 17 | 70-74 | Male | 3.5 | Stage III | 27 | 0 | 5 | 0, none | | X | |
| 18 | 85-89 | Female | 3.08 | Stage III | 28 | 0 | 6 | 0, none | 3/4 | X | |
| 19 | 75-79 | Female | 2.5 | Stage II | 28 | 0 | 3 | 0, none | 3/3 | X | X |
| 20 | 75-79 | Male | 4.3 | Stage II | 28 | 0 | 2 | 0, none | 2/3 | | X |
| 21 | 90+ | Male | 3.41 | Stage I | 27 | 1 | 2 | 1, sparse | 3/3 | X | X |
| 22 | 70-74 | Male | 4.6 | Stage I | 29 | 0 | 2 | 0, none | | X | |
| 23 | 90+ | Male | 3 | Stage I | 30 | 0 | 2 | 0, none | | X | |
| **Young Control** | | | | | | | | | | | |
| 24 | 50-54 | Female | 3.95 | Stage II | | 7 | 3 | 0, none | | | X |
| 25 | 60-64 | Male | 2.33 | Stage I | | 1 | 1 | 1, sparse | | X | X |
| 26 | 55-59 | Female | 3.15 | Stage I | | 2 | 1 | 0, none | | X | |
| 27 | 50-54 | Female | 4.67 | Stage I | | 0 | 1 | 0, none | 3/3 | X | X |
| 28 | 35-39 | Male | 3 | Stage 0 | | 0 | 0 | 0, none | 3/3 | X | X |
| 29 | 45-49 | Male | 4.5 | Stage 0 | | 0 | 0 | 0, none | 3/3 | X | X |

[a] Total amyloid plaque density score in the following regions: frontal, temporal, parietal, hippocampus, and entorhinal cortex. Region scores: 0 = none; 1 = sparse; 2 = moderate; 3 = frequent.

[b] Includes 26 cases for middle temporal gyrus and 25 cases for hippocampus and dentate gyrus.

[c] Subsets of 12 cases each were immunostained for pPI3K and pLIMK1 (see Table S3) and pPSD95 (see Table S4), respectively.

**Table S3. Demographic, neuropathological, and cognitive characteristics of twelve cases spanning the clinicopathological spectrum of Alzheimer's disease progression (pLIMK1 & pPI3K)**

| | AD (n=3) | MCI (n=3) | Control (n=3) | Young Control (n=3) |
|---|---|---|---|---|
| **Demographic and clinical characteristics** | | | | |
| Age, y, mean (SD) [a] | 83.3 (1.5) | 85.3 (8.1) | 81.3 (7.8) | 54.3 (6.1) |
| Education, y, mean (SD) | 15.0 (1.0) | 13.0 (1.7) | 16.3 (0.6) | 14.0 (2.8) |
| Female, n | 2 | 3 | 1 | 1 |
| Post mortem interval, hours, mean (SD) | 3.8 (0.3) | 3.2 (0) | 3.4 (0.9) | 3.6 (1.1) |
| **Neuropathological characteristics** | | | | |
| Braak stage (0-6), mean (range) | 5.7 (5-6) | 4.0 (4-4) | 1.7 (1-2) | 1.0 (0-2) |
| Neurofibrillary tangle (0-15), total, mean (SD) [b] | 14.3 (1.2) | 8.7 (0.6) | 2.3 (0.6) | 1.3 (1.5) |
| Entorhinal cortex (0-3), mean (SD) | 3.0 (0) | 3.0 (0) | 1.3 (0.6) | 0.7 (0.6) |
| Hippocampus (0-3), mean (SD) | 3.0 (0) | 3.0 (0) | 1.0 (0) | 0 (0) |
| Temporal cortex (0-3), mean (SD) | 3.0 (0) | 1.0 (0) | 0 (0) | 0.3 (0.6) |
| Neuritic plaque density (0-3), mean (SD) | 3.0 (0) | 2.0 (1.7) | 0.3 (0.6) | 0.3 (0.6) |
| Amyloid plaques (0-15), total, mean (SD) [b] | 14.7 (0.6) | 8.3 (4.9) | 0.3 (0.6) | 2.7 (3.8) |
| Entorhinal cortex (0-3), mean (SD) | 3.0 (0) | 1.3 (1.5) | 0 (0) | 0.7 (1.2) |
| Hippocampus (0-3), mean (SD) | 2.7 (0.6) | 1.0 (1.0) | 0 (0) | 0 (0) |
| Temporal cortex (0-3), mean (SD) | 3.0 (0) | 1.7 (1.2) | 0 (0) | 1.0 (1.7) |
| **Cognitive endpoints** | | | | |
| Cognitive dysfunction, y, mean (SD) | 10.3 (4.0) | n/a | n/a | n/a |
| MMSE (0-30), mean (SD) | 11.7 (8.7) | 24.0 (2.0) | 27.7 (0.6) | n/a |
| Clinical Dementia Rating Sum of Boxes (0-18), mean (SD) | 12.3 (3.2) | 2.0 (1.0) | 0 (0) | n/a |
| Clinical Dementia Rating global score (0-3), mean (SD) | 2.3 (0.6) | 0.5 (0) | 0 (0) | n/a |
| FAST Score (1-7), mean (SD) | 5.3 (0.6) | 2.3 (0.6) | 1.7 (0.6) | n/a |
| Figure Recall Score (0-3), mean (SD) | 0.3 (0.6) | 0.7 (0.6) | 2.3 (0.6) | n/a |
| AVLT Total Learning (0-75), mean (SD) | 14.7 (2.3) | 32.0 (13.1) | 39.3 (7.6) | n/a |
| AVLT STM A6 (0-15), mean (SD) | 1.3 (2.3) | 4.7 (2.1) | 10.3 (0.6) | n/a |
| WMSR Digit Span Forward Score (0-12), mean (SD) | 7.0 (0) | 9.0 (1.4) | 7.7 (0.6) | n/a |
| ApoE | | | | |
| 3/3, n | 2 | 1 | 2 | 1 |
| 2/3, n | 0 | 2 | 1 | 0 |
| 3/4, n | 1 | 0 | 0 | 0 |
| 2/2, n | 0 | 0 | 0 | 0 |
| NIA-Reagan, Likelihood of Alzheimer's disease [c] | | | | |
| Not Alzheimer's disease, n | 0 | 0 | 0 | 0 |
| Low, n | 0 | 0 | 0 | 1 |
| Intermediate, n | 0 | 0 | 0 | 0 |
| High, n | 3 | 0 | 0 | 0 |
| Criteria Not Met, n | 0 | 3 | 3 | 2 |
| No Dementia NOS, n | 3 | 3 | 3 | 3 |
| No Clinpath HS, n | 3 | 3 | 3 | 3 |
| No Clinpath VaD, n | 3 | 3 | 3 | 3 |

[a] Cases that were ≥90 years of age were classified by BBDP as 90 years of age or older.

[b] Total scores for plaques and tangles include the entorhinal, hippocampus, temporal, parietal, and frontal areas. Each was scored according to the CERAD templates [1] using Campbell-Switzer silver stain, Gallyas silver stain and Thioflavin S stains.

[c] Modified NIA-Reagan diagnosis of Alzheimer's disease based on consensus recommendations for postmortem diagnosis of Alzheimer's disease [2] including neurofibrillary tangles (Braak) and neuritic plaques (CERAD).

**Table S4. Demographic, neuropathological, and cognitive characteristics of twelve cases spanning the clinicopathological spectrum of Alzheimer's disease progression (pPSD95)**

| | AD (n=3) | MCI (n=4) | Control (n=2) | Young Control (n=3) |
|---|---|---|---|---|
| **Demographic and clinical characteristics** | | | | |
| Age, y, mean (SD) [a] | 83.0 (7.0) | 85.2 (6.6) | 82.5 (10.6) | 46.3 (7.4) |
| Education, y, mean (SD) | 14.3 (2.1) | 12.5 (1.0) | 16.5 (0.7) | 15.3 (1.2) |
| Female, n | 2 | 2 | 1 | 1 |
| Post mortem interval, hours, mean (SD) | 3.1 (0.9) | 2.9 (0.4) | 3.0 (0.6) | 4.1 (0.9) |
| **Neuropathological characteristics** | | | | |
| Braak stage (0-6), mean (range) | 5.0 (5-5) | 3.5 (3-4) | 1.5 (1-2) | 0.3 (0-1) |
| Neurofibrillary tangle (0-15), total, mean (SD) [b] | 13.0 (2.0) | 6.8 (2.2) | 2.5 (0.7) | 0.3 (0.6) |
| Entorhinal cortex (0-3), mean (SD) | 3.0 (0) | 2.8 (0.5) | 1.5 (0.7) | 0.3 (0.6) |
| Hippocampus (0-3), mean (SD) | 3.0 (0) | 2.2 (1.0) | 1.0 (0) | 0 (0) |
| Temporal cortex (0-3), mean (SD) | 3.0 (0) | 1.0 (0) | 0 (0) | 0 (0) |
| Neuritic plaque density (0-3), mean (SD) | 3.0 (0) | 1.5 (1.3) | 0.5 (0.7) | 0 (0) |
| Amyloid plaques (0-15), total, mean (SD) [b] | 14.7 (0.6) | 7.5 (5.4) | 0.5 (0.7) | 0 (0) |
| Entorhinal cortex (0-3), mean (SD) | 3.0 (0) | 1.8 (1.5) | 0 (0) | 0 (0) |
| Hippocampus (0-3), mean (SD) | 2.7 (0.6) | 0.8 (1.0) | 0 (0) | 0 (0) |
| Temporal cortex (0-3), mean (SD) | 3.0 (0) | 1.8 (1.0) | 0 (0) | 0 (0) |
| **Cognitive endpoints** | | | | |
| Cognitive dysfunction, y, mean (SD) | 7.3 (0.6) | n/a | n/a | n/a |
| MMSE (0-30), mean (SD) | 11.0 (4.4) | 26.5 (1.9) | 27.5 (0.7) | n/a |
| Clinical Dementia Rating Sum of Boxes (0-18), mean (SD) | 11.3 (6.1) | 1.5 (1.3) | 0 (0) | n/a |
| Clinical Dementia Rating global score (0-3), mean (SD) | 2.0 (1.0) | 0.5 (0) | 0 (0) | n/a |
| FAST Score (1-7), mean (SD) | 5.0 (1.0) | 2.5 (0.6) | 1.5 (0.7) | n/a |
| Figure Recall Score (0-3), mean (SD) | 0.7 (0.6) | 1.2 (1.0) | 2.5 (0.7) | n/a |
| AVLT Total Learning (0-75), mean (SD) | 11.5 (6.4) | 34.8 (13.2) | 38.5 (10.6) | n/a |
| AVLT STM A6 (0-15), mean (SD) | 0 (0) | 4.8 (2.1) | 10.5 (0.7) | n/a |
| WMSR Digit Span Forward Score (0-12), mean (SD) | 8.0 (0) | 9.2 (2.2) | 8.0 (0) | n/a |
| ApoE | | | | |
| 3/3, n | 2 | 2 | 2 | 3 |
| 2/3, n | 0 | 2 | 0 | 0 |
| 3/4, n | 1 | 0 | 0 | 0 |
| 2/2, n | 0 | 0 | 0 | 0 |
| NIA-Reagan, Likelihood of Alzheimer's disease [c] | | | | |
| Not Alzheimer's disease, n | 0 | 0 | 0 | 0 |
| Low, n | 0 | 0 | 0 | 0 |
| Intermediate, n | 0 | 0 | 0 | 0 |
| High, n | 3 | 0 | 0 | 0 |
| Criteria Not Met, n | 0 | 4 | 2 | 3 |
| No Dementia NOS, n | 3 | 4 | 2 | 3 |
| No Clinpath HS, n | 3 | 4 | 2 | 3 |
| No Clinpath VaD, n | 3 | 4 | 2 | 3 |

[a] Cases that were ≥90 years of age were classified by BBDP as 90 years of age or older.
[b] Total scores for plaques and tangles include the entorhinal, hippocampus, temporal, parietal, and frontal areas. Each was scored according to the CERAD templates [1] using Campbell-Switzer silver stain, Gallyas silver stain and Thioflavin S stains.
[c] Modified NIA-Reagan diagnosis of Alzheimer's disease based on consensus recommendations for postmortem diagnosis of Alzheimer's disease [2] including neurofibrillary tangles (Braak) and neuritic plaques (CERAD).

**Table S5. Key Resources**

| *Reagent type or resource* | *Target including Domain* | *Designation* | *Type* | *Source or reference* | *Identifiers or sequence* | *Comments* |
|---|---|---|---|---|---|---|
| | **Probes** | | | | | |
| **Multidomain labeling of ApoE Receptors** | | | | | | |
| | **ApoER2** | | | | | |
| ISH probe | ApoER2 mRNA probe | ApoER2-mRNA | ISH probe | ACD, Biotechne | 807461 | |
| antibody | ApoER2 LA Repeat 1-2 transition (aa73-85)* | Anti-ApoER2 LA1-2 | Rabbit IgG polyclonal | New England Peptide | Not applicable | IHC 1:100-1:150; WB 1:1000 |
| antibody | ApoER2 LA Repeat (aa149-164) | Anti-ApoER2 LA3b2 | Mouse IgG2b [LB3-8G7] | Diagnocine | BML033 | IHC 1:100; WB 1:500 |
| antibody | ApoER2 LA Repeat (aa149-164) | Anti-ApoER2 LA3b3 | Mouse IgG2a [LB3-10B6] | Diagnocine | BML034 | IHC 1:100; WB 1:400 |
| antibody | ApoER2 LA Repeat (aa149-160)* | Anti-ApoER2 LA3b | Rabbit IgG polyclonal | New England Peptide | Not applicable | IHC 1:150; WB 1:1000 |
| antibody | ApoER2 Beta-propeller domain (aa524-573) | Anti-ApoER2 YWTD | Rabbit IgG polyclonal | Millipore-Sigma | SAB2103110 | IHC 1:100 |
| antibody | ApoER2 C-term Proline-rich (aa914-938) | Anti-ApoER2 C2 | Mouse IgG2b [ER2-CyB-5G7] | Diagnocine | BML036 | IHC 1:100 |
| | **VLDLR** | | | | | |
| antibody | VLDLR (aa225-239) | Anti-VLDLR-LA5-6 | Mouse | Diagnocine | BML031 | WB 1:1000 |
| **Ligands for ApoE Receptors** | | | | | | |
| | **Native ApoE** | | | | | |
| antibody | ApoE (aa51-100) | ApoE-N-terminal | Rabbit IgG | Thermo Fisher | PA5-86589 | IHC 1:100 |
| antibody | ApoE (aa140-160) | ApoE-Bind 1 | Mouse IgG1 [WUE4] | Novus Biologicals | NB110-60531 | IHC 1:100, WB 1:500 |
| antibody | ApoE C-term unspecified | ApoE-C-terminal | Rabbit IgG [EP1374Y] | Abcam | ab52607 | IHC 1:50-1:100 |
| antibody | ApoE (aa1-299) | ApoE | Rat IgG | Precision Antibody | Not applicable | IHC 1:100; WB 1:200 |
| antibody | ApoE (aa1-299) | ApoE | Chicken IgG | New England Peptide | Not applicable | IHC 1:100; WB 1:200 |
| | **Lipid peroxide modified ApoE** | | | | | |

| *Reagent type or resource* | *Target including Domain* | *Designation* | *Type* | *Source or reference* | *Identifiers or sequence* | *Comments* |
|---|---|---|---|---|---|---|
| antibody | Cu-oxidized PAPC-modified ApoE [15E8]* | PxPAPC-ApoEa | Rat IgG [15E8] | Precision Antibody | Not applicable | IHC 1:50; WB 1:200 |
| antibody | Cu-oxidized PAPC-modified ApoE [2B7]* | PxPAPC-ApoEb | Rat IgG [2B7] | Precision Antibody | Not applicable | IHC 1:50; WB 1:200 |
| antibody | Hydroxy-nonenal-modified ApoE* | HNE-ApoE | Chicken IgY | New England Peptide | Not applicable | IHC 1:500; WB 1:1000 |
| antibody | Crotonaldehyde-modified ApoE* (non-selective) | CRA-ApoE | Chicken IgY | New England Peptide | Not applicable | WB 1:1000 |
| antibody | Acetaldehyde-modified ApoE* (non-selective) | ACET-ApoE | Chicken IgY | New England Peptide | Not applicable | WB 1:1000 |
| antibody | KOdiA-PC modified ApoE* (non-selective) | KOdiA-ApoE | Chicken IgY | New England Peptide | Not applicable | WB 1:1000 |
| | **Reelin** | | | | | |
| antibody | Reelin (aa3239-3460) | Reelin-C | Mouse IgG2a [E-5] | Santa Cruz | sc25346 | IHC 1:50; WB 1:250 |
| **Reelin-ApoE receptor signaling cascades** | | | | | | |
| | **Downstream Reelin-ApoE receptor cascade** | | | | | |
| antibody | Disabled homolog 1 | DAB1 | Rabbit IgG | Thermo/Invitrogen | PA5-86617 | IHC 1:50 |
| antibody | Tyr607-phosphorylated Phosphatidylinositol 3-Kinase (p85a) | Tyr607-pPI3K | Rabbit IgG | Thermo/Invitrogen | PA5-104853 | IHC 1:100 |
| antibody | Thr508-phosphorylated LIM kinase-1 | Thr508-pLIMK1 | Rabbit IgG | Thermo/Invitrogen | PA5-104925 | IHC 1:100 |
| antibody | Thr19-phosphorylated PSD95 (DLG4) | Thr19-pPSD95 | Rabbit IgG | Millipore-Sigma | ABN998 | IHC 1:50-1:100 |
| | **Classic Alzheimer's pathologies** | | | | | |
| antibody | Amyloid Beta Protein | Aβ | Mouse IgG2b [MOAB-2] | Novus Biologicals | NBP2-13075 | IHC 1:100 |
| antibody | Ser202/Thr205-phosphorylated Tau | Ser202/Thr205-pTau | Mouse IgG1 [AT8] | Invitrogen | MN1020 | IHC 1:100 |
| **Cytoarchitectural markers** | | | | | | |
| | **Neurons** | | | | | |
| antibody | Neuronal marker NeuN (antibody 1) | NEUN | Millipore Sigma | ABN90P | Guinea Pig IgG | IHC 1:100 |
| antibody | Neuronal marker NeuN (antibody 2) | NEUN-2 | | | | |

| *Reagent type or resource* | *Target including Domain* | *Designation* | *Type* | *Source or reference* | *Identifiers or sequence* | *Comments* |
|---|---|---|---|---|---|---|
| antibody | Microtubule associated protein 2 | MAP2 | Mouse IgG3 [885232] | R&D Systems | MAB8304 | IHC 1:100 |
| antibody | Neurofilament light chain | NFL | Mouse IgG1 | Biolegend | 846002 | IHC 1:100 |
| antibody | Neurofilament light chain | NFL-2 | Mouse IgG1 | Biolegend | 845902 | IHC 1:100 |
| antibody | Synaptophysin | SYNAP | Mouse IgM [SP15] | Millipore Sigma | MAB328 | IHC 1:100 |
| | **Astrocytes** | | | | | |
| antibody | Glial fibrillary acidic protein | GFAP | Rat IgG2a [2.2B10] | Thermo Fisher Sci | 13-0300 | IHC 1:100 |
| antibody | Glial fibrillary acidic protein | GFAP-2 | Rabbit IgG | Agilent/Dak o | Z033429-2 | IHC 1:100 |
| | **Microglia** | | | | | |
| antibody | IBA1 | IBA1 | Chicken IgY | Synaptic Systems | 234006 | IHC 1:100 |
| antibody | CD68 | CD68 | Mouse IgG3 [PG-M1] | Abcam | ab783 | IHC 1:100 |
| antibody | Adipophilin | ADFP | Mouse IgG2b [1366] | Abcam | ab219299 | IHC 1:30 |
| | **Oligodendrocytes** | | | | | |
| antibody | CNPase | CNPase | Mouse IgG2b | Novus Biologicals | NBP2-46617 | IHC 1:100 |
| | **Nuclei** | | | | | |
| | DAPI | DAPI | | Invitrogen | D1306 | 1ug/mL |
| | **Secondary Antibodies** | | | | | |
| | **Single-marker IHC (HRP)** | | | | | |
| antibody | Goat anti-Rat IgG | | | Jackson | 112-035-167 | 1.6 ug/mL |
| antibody | Goat anti-Mouse IgG1 | | | Jackson | 115-035-205 | 1.6 ug/mL |
| antibody | Goat anti-Mouse IgG2a | | | Jackson | 115-035-206 | 1.6 ug/mL |
| antibody | Goat anti-Mouse IgG2b | | | Jackson | 115-035-207 | 1.6 ug/mL |
| antibody | Goat anti-Mouse IgG3 | | | Jackson | 115-035-209 | 1.6 ug/mL |
| antibody | Goat anti-Mouse IgM | | | Jackson | 115-035-075 | 1.6 ug/mL |
| antibody | Donkey anti-Rabbit IgG | | | Jackson | 711-035-152 | 1.6 ug/mL |
| antibody | Donkey anti-Chicken IgY | | | Jackson | 703-035-155 | 1.6 ug/mL |
| | **Western blot** | | | | | |

| ***Reagent type or resource*** | ***Target including Domain*** | ***Designation*** | ***Type*** | ***Source or reference*** | ***Identifiers or sequence*** | ***Comments*** |
|---|---|---|---|---|---|---|
| antibody | AzureSpectra Fluorescent Goat-anti-mouse 650 | | | Azure Biosystems | AC2166 | 1:10,000 |
| antibody | AzureSpectra Fluorescent Goat-anti-rabbit 650 | | | Azure Biosystems | AC2165 | 1:10,000 |
| antibody | AzureSpectra Fluorescent Goat-anti-rat 800 | | | Azure Biosystems | AC2138 | 1:10,000 |
| antibody | AzureSpectra Fluorescent Goat-anti-chicken 650 | | | Azure Biosystems | AC2168 | 1:10,000 |
| antibody | AzureSpectra Fluorescent Goat-anti-mouse 800 | | | Azure Biosystems | AC2135 | 1:10,000 |
| | **Peptides** | | | | | |
| synthetic peptide | ApoE binding region (aa138-148) | | | New England Peptide | Ac-ASHLRKLRKRL | |
| synthetic peptide | ApoE binding region (-Lys )(aa138-148) | | | New England Peptide | Ac-ASHLRALRARL | |
| synthetic peptide | ApoE binding region (-His ) (aa138-148) | | | New England Peptide | Ac-ASALRKLRKRL | |
| synthetic peptide | ApoER2 binding region (aa73-85) | | | New England Peptide | Ac-LDHSDEDD(C/Ac-C)PKKT-NH2 | |
| synthetic peptide | ApoER2 binding region (-Lys )(aa73-85) | | | New England Peptide | Ac-LDHSDEDD(C/Ac-C)PAAT-NH2 | |
| synthetic peptide | ApoER2 binding region (-His )(aa73-85) | | | New England Peptide | Ac-LDASDEDD(C/Ac-C)PKKT-NH2 | |
| | **Recombinant Proteins** | | | | | |
| protein | Human ApoE4 (monomer) | | | Abcam | ab50243 | |
| protein | Human ApoER2 ectodomain | | | R&D | 3520-AR-050 | |
| protein | Human ApoER2 including C-term | | | Origene | TP320903 | |
| protein | Human VLDLR (Gly28-Ser797) | | | Sigma Aldrich | SRP6469 | |
| | **Lipids and aldehydes** | | | | | |
| lipid | 1-palmitoyl-2-arachidonoyl-sn-glycero-3-phosphocholine | PAPC; 16:0-20:4 PC | | Avanti Polar Lipids | 850459 | |

| ***Reagent type or resource*** | ***Target including Domain*** | ***Designation*** | ***Type*** | ***Source or reference*** | ***Identifiers or sequence*** | ***Comments*** |
|---|---|---|---|---|---|---|
| lipid | 1-(palmitoyl)-2-(5-keto-6-octene-dioyl)phosphatidylcholine | KOdiA-PC | | Cayman Chemical | 62945 | |
| aldehyde | Acetaldehyde | ACET | | Millipore-Sigma | 402788 | |
| aldehyde | Acrolein | ACR | | Millipore-Sigma | 89116 | |
| aldehyde | Crotonaldehyde | CRA | | Millipore-Sigma | 27980 | |
| aldehyde | Malondialdehyde tetrabutylammonium salt | MDA | | Millipore-Sigma | 63287 | |
| aldehyde | 4-Hydroxynonenal | 4-HNE | | Cayman Chemical | 32100 | |
| aldehyde | 4-oxo-2-nonenal | 4-ONE | | Cayman Chemical | 10185 | |
| aldehyde | Reactive Aldehyde Mix | RAM | | See above | See above | equimolar mix of CRA, MDA, 4-HNE, ACET |
| | **Reagents for Western Blot** | | | | | |
| | 8-16% MP TGX precast gel | | | Biorad | 4561106 | |
| | 4X Laemmli Sample Buffer | 1x | | Biorad | 1610747 | |
| | 2-Mercaptoethanol | | | Sigma-Aldrich | M6250-100ML | |
| | Precision Plus Protein Dual Color | | | Biorad | 1610374 | 1:10 |
| | 10x Tris/Glycine/SDS running buffer | | | Biorad | 1610772 | |
| | 10x Tris/Glycine Transfer Buffer | | | Biorad | 1610771 | |
| | Methanol | | | Sigma-Aldrich | 494437-2L | |
| | Ponceau S solution | | | Sigma-Aldrich | P7170-1L | |
| | Azure Fluorescent Blot Blocking Buffer | | | Azure Biosystems | AC2190 | |
| | AzureRed Fluorescent Total Protein Stain | | | Azure Biosystems | AC2124 | |
| | 10X PBS | | | Quality Biological | 119-069-101 | |
| | Tween 20 | | | Sigma | P2287-100mL | |
| | PVDF membrane, Immobilon-FL | | | MilliporeSigma | IPFL00005 | |

### III. Supplementary Materials and Methods

#### Biochemical Experiments

Peptides, proteins, lipids, aldehydes, and other reagents were obtained as indicated in **Table S5**. Snakeskin dialysis tubing and Slide-A-lyzer dialysis units were obtained from Thermo Fisher. Spectra/Por dialysis apparatus and tubing were obtained from Repligen (Waltham, MA). Dialyses were performed using appropriate MWCO for protein or peptide molecular weights against 3 changes of the buffers specified below. KPhos refers to potassium phosphate buffer, pH 7.4 (Sigma, St. Louis, MO). 10mM PBS refers to 1x PBS, pH 7.4 (Quality Biological, Gaithersburg, MD).

##### ***Antigen conjugate preparation***

*Cu-oxidized PAPC-modified ApoE:* In a vial, a chloroform solution of 6.24mg of PAPC (1-palmitoyl-2-arachidonoyl-sn-glycero-3-phosphocholine) was evaporated to dryness under a stream of Argon. 15ml of 0.1M KPhos (pH7.4) containing 10mM $CuSO_4$ was added and it was vortexed. 2mg of ApoE4 monomer was added, it was vortexed again, and then incubated with gentle stirring at 37°C for 20 hours. The mixture was dialyzed extensively using snakeskin dialysis tubing (10kDa MWCO) against 0.1M KPhos+0.25 mM EDTA (pH7.4) (4L). The sample was lyophilized.

*CRA-modified ApoE:* A mixture of ApoE4 monomer (6mg) in 100mM crotonaldehyde in 10mM PBS (20ml) was incubated at room temperature for 3 days, then dialyzed against 10mM KPhos+0.25mM EDTA (4L).

*HNE-modified ApoE:* A solution of 25mg of 4-hydroxynonenal in 2ml EtOH was added to 15ml of 10mM PBS, turning cloudy. Then 5mg ApoE4 monomer was added, and it was mixed thoroughly. The mixture was incubated at room temperature for 4 days, then dialyzed against 10mM KPhos (4L).

*KOdiA-modified ApoE:* A mixture of ApoE4 monomer (6mg) in 10mM PBS (20ml) containing 20mg of KOdiA-PC was incubated at room temperature for 3 days, then dialyzed against 10mM KPhos (4L).

*ACET-modified ApoE:* To a solution of 100mM acetaldehyde in 10mM PBS (20ml) was added 5.5mg of ApoE4 monomer. It was incubated at room temperature for 3 days and then a slight excess of $NaCNBH_3$ (~50mg) was added and it was incubated overnight. Then the mixture was dialyzed against 10mM KPhos (4L).

##### ***Peptide adduction and pH dependent reversibility experiments***

Peptides were made by New England Peptide/Vivitide (Gardner, MA) using standard Fmoc solid-state peptide synthesis and purified using high-performance liquid chromatography (HPLC) to >85% pure as determined by HPLC and/or LC/MS.

*Peptide MDA adduction and pH dependent reversibility:* 5mg of peptide (ApoER2 LA1-2 [Ac-LDHSDEDD(C/Ac-C)PKKT-$NH_2$] or ApoE binding domain [Ac-ASHLRKLRKRL] was added to a solution of 1mM or 10mM MDA in 10mM PBS (4ml), and the mixture was incubated at 37°C for 48hrs. Aliquots were removed for LC-MS analysis at 1hr, 3hr, 5hr, 24hr, and 48hr. At the end of the reaction, the mixture was dialyzed against 10mM PBS. For the ApoER2 LA1-2 and ApoE binding domain adducts, approximately 1.5ml of the dialyzed solution was used to monitor the effect of acidic pH on the adduct stability, by incubating at 37°C in 10mM PBS adjusted with 0.1M HCl to pH6 or pH4. Reactions were monitored by LC-MS over 48hr as above.

*Reactive aldehyde mixture ApoER2 and ApoE peptide adduction and pH dependent reversibility:* 5mg of the ApoER2 or ApoE binding domain peptide was added to a solution of 10mM PBS (4ml) containing 10mM each of MDA, acetaldehyde, CRA and 4-HNE and incubated at 37°C for 48hrs. Aliquots were removed for LC-MS analysis at 3hr, 5hr, 24hr, and 48hr and dialyzed against 10mM PBS. Approximately 1.5ml of the dialyzed solution was used to monitor the effect of acidic pH on the stability of the adducts, by incubating at 37°C at pH6 or pH4. Reactions were monitored by LC-MS over 48hr as above.

*ApoER2 peptide analog (-Lys) MDA adduct:* An analog of the ApoER2 LA1-2 peptide with sequence Ac-LDHSDEDD(C/Ac-C)PAAT-$NH_2$ (1mg) was incubated in 10mM MDA (10mM PBS; 1ml) at 37°C for 48hrs. Aliquots were removed for LC-MS analysis at 24hr and 48hr, dialyzed against 10mM PBS and analyzed by LC-MS.

*ApoE peptide analog (-Lys) MDA adduct:* An analog of the ApoE binding domain peptide with sequence Ac-ASHLRALRARL (1mg) was incubated in 10mM MDA (10mM PBS; 1ml) at 37°C for 48hrs. Aliquots were removed for LC-MS analysis at 24hr and 48hr, dialyzed against 10mM PBS and analyzed by LC-MS.

***Peptide crosslinking experiments***

*ApoER2 LA1-2 + ApoE peptide 4-ONE crosslinking:* A solution of 1mg (0.586μmol) of *ApoER2 LA1-2* peptide and 0.83mg (0.586μmol) of ApoE peptide was incubated in 10mM PBS (586μl) at 37°C for 20min. 109μg of 4-ONE (0.702μmol) in 21.8μl of EtOH was added, and it was incubated overnight. The mixture was analyzed by LC-MS, and further characterized by TOF-MS after it was dialyzed in a 1ml Slide-A-lyzer against 10mM ammonium acetate (200ml).

*ApoER2 LA1-2 peptide analog (-His) + ApoE peptide 4-ONE crosslinking:* A solution of 1mg (0.610μmol) of ApoER2 peptide analog (-His) with sequence LDASDEDD(C/Ac-C)PKKT-$NH_2$ and 0.86mg (0.610μmol) of

ApoE binding region peptide was incubated in 10mM PBS (610µl) at 37°C for 20min. 113µg (0.732µmol) of 4-ONE in 22.6µl of EtOH was added, and it was incubated overnight, followed by analysis with LC-MS.

*ApoER2 LA1-2 peptide + ApoE peptide analog (-Lys) 4-ONE crosslinking:* A solution of 1mg (0.586µmol) of ApoER2 LA1-2 peptide and 0.76mg (0.586µmol) of ApoE peptide analog with sequence Ac-ASHLRALRARL was incubated in 10mM PBS (586µl) at 37°C for 20min. 109µg (0.702µmol) of 4-ONE in 21.8µl of EtOH was added, and it was incubated overnight, followed by analysis with LC-MS.

*ApoER2 LA1-2 peptide + ApoE peptide analog (-His) 4-ONE crosslinking:* A solution of 1mg (0.586µmol) of ApoER2 LA1-2 peptide and 0.794mg (0.586µmol) of ApoE peptide analog with sequence Ac-ASALRKLRKRL was incubated in 10mM PBS (586µl) at 37°C for 20min. 109µg (0.702µmol) of 4-ONE in 21.8µl of EtOH was added, and it was incubated overnight, followed by analysis with LC-MS.

*ApoER2 LA1-2 peptide analog (-His) + ApoE peptide analog (-His) 4-ONE crosslinking:* A solution of 1mg (0.610µmol) of ApoER2 LA1-2 peptide analog (-His) with sequence LDASDEDD(C/Ac-C)PKKT-$NH_2$ and 0.83mg (0.610µmol) of ApoE peptide analog with sequence Ac-ASALRKLRKRL was incubated in 10mM PBS (610µl) at 37°C for 20min. 113µg (0.732µmol) of 4-ONE in 22.6µl of EtOH was added, and it was incubated overnight, followed by analysis with LC-MS.

*ApoE peptide + ApoER2 LA1-2 peptide analog (-Lys) 4-ONE crosslinking:* A solution of 1mg (0.628µmol) of ApoER2 LA1-2 peptide analog (-Lys) with sequence LDHSDEDD(C/Ac-C)PAAT-$NH_2$ and 0.89mg (0.628µmol) of ApoE peptide was incubated in 10mM PBS (628µl) at 37°C for 20min. 116µg (0.754µmol) of 4-ONE in 23.2µl of EtOH was added, and it was incubated overnight, followed by analysis with LC-MS.

*ApoE peptide analog (-Lys) + ApoER2 LA1-2 peptide analog (-Lys) 4-ONE crosslinking:* A solution of 1mg (0.628µmol) of ApoER2 LA1-2 peptide analog (-Lys) with sequence LDHSDEDD(C/Ac-C)PAAT-$NH_2$ and 0.82mg (0.628µmol) of ApoE peptide analog with sequence Ac-ASHLRALRARL was incubated in 10mM PBS (628µl) at 37°C for 20min. 116µg (0.754µmol) of 4-ONE in 23.2µl of EtOH was added, and it was incubated overnight, followed by analysis with LC-MS.

***Protein crosslinking experiments***

*ApoER2 + ApoE single aldehyde crosslinking:* A solution of 100µg ApoER2 ectodomain and 100µg of ApoE4 monomer in 10mM PBS (343µl) was incubated at 37°C for 10min, then one aldehyde (CRA, 4-ONE, 4-HNE, or MDA) was added to a final concentration of 10mM in 500µl. After 48hrs, the mixture was dialyzed against 10mM PBS.

*ApoER2 + ApoE reactive aldehydes mixture crosslinking:* A solution of 250µg of ApoER2 ectodomain and 250µg ApoE4 monomer in 10mM PBS (607µl) was incubated at 37°C for 10min, then a mixture of reactive aldehydes in PBS (consisting of 10mM each of MDA, CRA, acrolein, 4-HNE and 4-ONE) was added to a final concentration of 10mM per aldehyde in 1.25ml. After 48hrs, the mixture was dialyzed against 10mM PBS.

**Custom Antibody Generation and Orthogonal Validation**

A rabbit polyclonal IgG antibody raised against the ApoER2 LA1-2 domain was custom-generated and affinity purified by Vivitide (Gardner, MA, USA). Chicken polyclonal IgY polyclonal antibodies were raised against lipid aldehyde-modified ApoE (made by incubating ApoE monomers with one lipid aldehyde). IgY material purified from eggs was subjected to multiple 'negative' affinity purifications (with native ApoE columns), sequentially repeated until no further antibodies bound to the native ApoE columns (determined by an OD of <0.1). These were followed by 'positive' affinity purification using lipid aldehyde-modified ApoE specific columns (Vivitide). Rat monoclonal antibodies were raised against lipid peroxidation-modified ApoE by incubating ApoE monomers with Cu-oxidized 1-palmitoyl-2-arachidonoyl-sn-phosphatidylcholine (PAPC). Rat antibodies were custom generated, and affinity purified by Precision Antibody (Columbia, MD, USA). Two clones (15E8 [PxPAPC-ApoEa], and 2B7 [PxPAPC-ApoEb]) were selected based on selective immunoreactivity for Cu-oxidized PAPC-modified ApoE by ELISA.

These custom-generated antibodies underwent orthogonal validation using a combination of western blot (see **Fig 4** and **Extended Figs 4.2-4.3** and **5.3**), positive control (AD case) vs. negative control immunostaining, single-marker IHC with serial sections, multi-epitope labeling using independent antibodies, and MP-IHC. The ApoER2 LA1-2 antibody was also evaluated with ISH/RNA-protein co-detection (see **Fig 5** and **Extended Figs 5.2-5.3**) and was shown by western blot to detect the LA1-2 domain of ApoER2 but not VLDLR (**Extended Fig** 5.3). Multi-epitope labeling (see **Extended Fig 2.1**) allowed us to compare staining patterns for several independent antibodies targeting different epitopes within targeted proteins (see **Extended Fig 5.2**). MP-IHC allowed us to include several targets within a given molecular pathway of interest, and to concurrently label multiple cytoarchitectural markers, which provided additional spatial, morphological, and cytoarchitectural context for observations made using single antibodies.

#### Immunohistochemical Marker Quantitation

Stain positive area as a percentage of each annotated region was quantified using the HALO Area Quantification v2.2.1 module. Plaque-associated objects per $mm^2$ within each annotated region were identified and quantified using the HALO Object Colocalization v1.3 module with classifier function enabled (**Fig S1**).

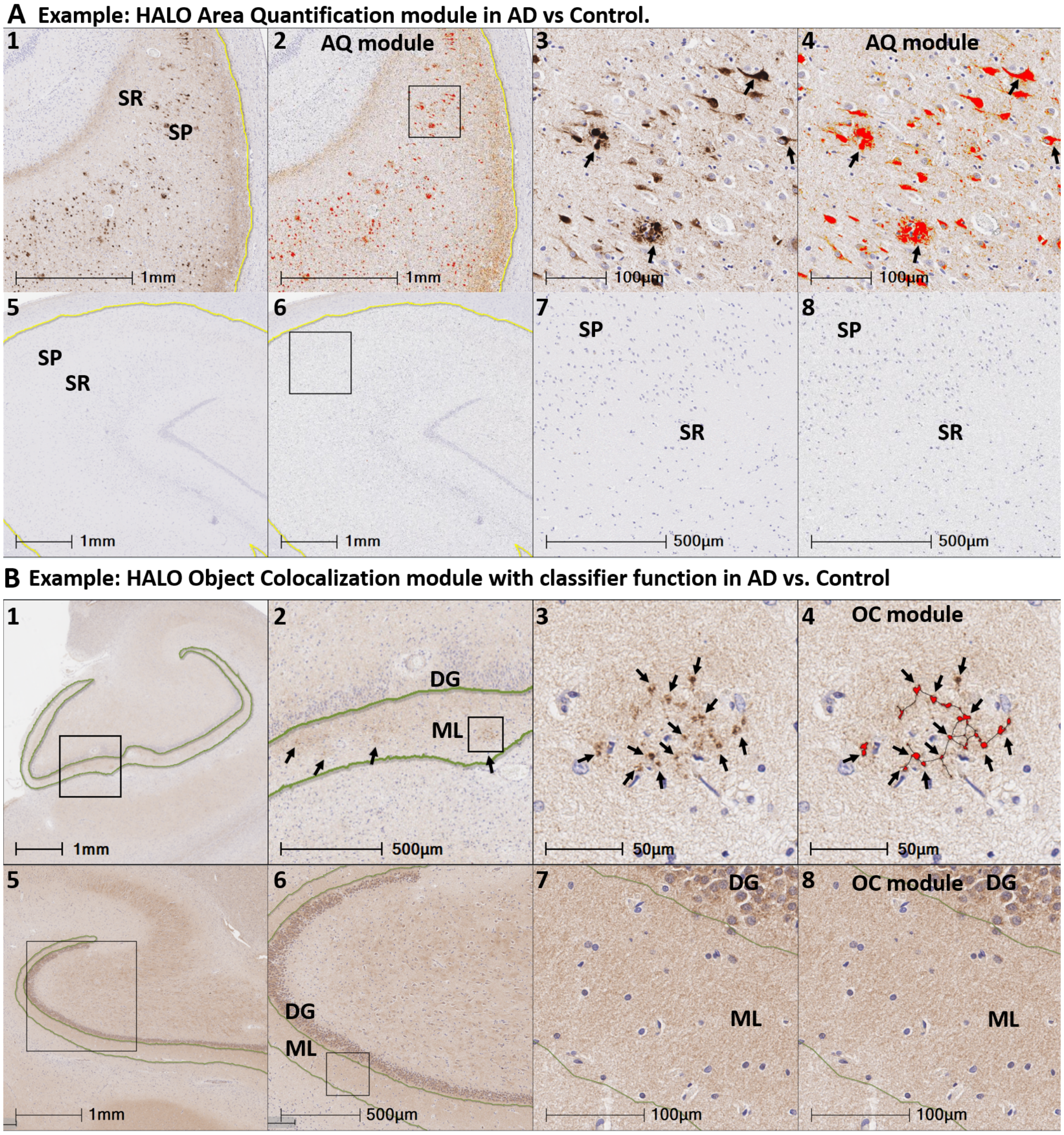


**Fig S1. Representative examples of HALO modules used for quantification of IHC markers.** Panels $\mathbf{A_{1\text{-}8}}$ illustrate the application of the HALO Area Quantification (AQ) module (v2.2.1) in the hippocampus of a Braak stage VI sAD case ($\mathbf{A_{1\text{-}4}}$) and a non-AD control ($\mathbf{A_{5\text{-}8}}$). Thr19-pPSD95 was strongly expressed in neuritic plaques and abnormal neurons (brown stain in panels $\mathbf{A_{1,3}}$) in sAD but was not detected in the non-AD control (lack of brown stain in panels $\mathbf{A_{5,7}}$). Thr19-pPSD95-positive structures that were identified by the AQ module are depicted in red in $\mathbf{A_{2,4}}$ and $\mathbf{A_{6,8}}$. Panels $\mathbf{B_{1\text{-}8}}$ illustrate the application of the HALO Object Colocalization (OC) v1.3 module. As shown in **Fig 5** and **Extended Fig 5.1,** ApoER2 LA1-2 was strongly expressed in discrete puncta surrounding a subset of neuritic plaques (black arrows in panels $\mathbf{B_{3\text{-}4}}$). The OC module selectively identified discrete ApoER2 LA1-2 aggregates without labeling diffuse, non-specific background staining that is consistent with synapses. A representative example of localized ApoER2 LA1-2-positive aggregates in one sAD case is shown in panels $\mathbf{B_{1\text{-}3}}$, with aggregates detected by the OC module depicted in red in panel $\mathbf{B_4}$. The lack of red in panel $\mathbf{B_8}$ indicates that no ApoER2 LA1-2 aggregates were detected.

**Neuropathological Assessments**

The neuropathological assessments and endpoints captured by BBDP are detailed in a previous publication [1]. Briefly, the Braak neurofibrillary stage (0- VI) was determined using thick 40 – 80-micron sections stained with Gallyas, Campbell-Switzer and thioflavine S stains as originally defined by Braak and Braak.[2] Senile plaque density—including neuritic, cored, and diffuse plaques—was assessed in standard regions of the frontal, temporal, and parietal lobes, hippocampal CA1 region and entorhinal/transentorhinal region. Each region was assigned a semi-quantitative score of none, sparse, moderate and frequent and converted to numerical values 0 – 3, according to the CERAD templates.[3] Plaque total is the arithmetic sum of scores from these five regions ranged from 0 – 15. Neurofibrillary tangle density was assessed in the same five regions, with CERAD templates used to obtain semi-quantitative scores of none, sparse, moderate and frequent and these are converted to numerical values 0 – 3. Tangle total is the arithmetic sum of scores from these five regions ranged from 0 – 15. The NIA-Reagan [4] consensus recommendations were used for postmortem diagnosis of AD with high, intermediate and low referring to the likelihood that dementia, if present, is due to AD histopathology. AD was at a minimum defined as intermediate or high NIA-Reagan criteria. Mild Cognitive Impairment denoted the presence of this diagnosis at the time of death. A control designation is a participant without dementia or parkinsonism during life and without a major neuropathological diagnosis.

*Braak Stage:* describing topographical progression of neurofibrillary tangles, dystrophic neurites and neuropil threads, throughout transentorhinal and entorhinal areas, CA1 subfield of hippocampus, amygdala and cerebral neocortex. Evaluations were made, similarly as the original publication,[2] in large (3 cm x 5 cm) thick (40 or 80 μm) sections stained with the Campbell-Switzer silver stain, Gallyas silver stain and Thioflavin S stains. Final judgment of tangle density is made on the basis of combined impression from all three stains. For three years, all cases were also stained with the AT8 antibody for phosphorylated tau protein. Note that the AT8 stain has been reported to give higher Braak stages as more neurites are apparent.[5]

*Tangle Total:* Average neurofibrillary tangle density in the cortex of the frontal lobe, including superior, middle, and inferior frontal gyri; cortex of the temporal lobe; cortex of the parietal lobe; CA1 subfield of hippocampus; and entorhinal cortex. Tangle density scored according to the CERAD templates,[3] as described for the Braak stage above.

*Plaque Total:* Average senile (amyloid) plaque density (all types of plaques considered together) in the cortex of frontal lobe, including superior, middle, and inferior frontal gyri; cortex of temporal lobe; cortex of parietal lobe; CA1 subfield of hippocampus; and entorhinal cortex. Plaque density scored according to CERAD templates,[3] using large (3 cm x 5 cm) thick (40 or 80 μm) sections stained with Campbell-Switzer and Gallyas silver stains, and Thioflavin S stains. Validity and accuracy of this combination for estimating density of Aβ deposits established in BBDP laboratories through strong correlations with autoradiographic binding of Florbetapir (amyloid imaging ligand), with biochemical measures (ELISA) of Aβ in human cerebral cortex extracts and with quantitative measures (percentage of section area occupied) of an immunohistochemical stain for Aβ.[1]

*Neuritic Plaque Density:* Greatest neuritic plaque density observed across frontal, temporal and parietal cortex regions, scored according to CERAD templates.[3] Evaluations made in large (3 cm x 5 cm) thick (40 or 80 μm) sections stained with the Campbell-Switzer silver stain, Gallyas silver stain and Thioflavin S stains. Final judgment of plaque density made on the basis of the combined impression from all three stains.[1]

**Cognitive Assessments**

The cognitive examinations and endpoints captured by BBDP are detailed in a previous publication [1] and are summarized below.

*MMSE Test Score:* Folstein Mini Mental State Examination score (0-30) obtained most proximal to death; includes MMSE scores obtained through BBDP research clinical visits and by review of private medical records.[6]

*CDR Sum:* Sum of Boxes (subsection) of the Clinical Dementia Rating (CDR) Scale. CDR is widely used for staging dementia severity.[7]

*FAST Score:* FAST is a functional assessment based both on caregiver report and clinician's observations: 1 = normal; 2 = subjective (only) report of forgetfulness or work difficulties; 3 = observed early executive dysfunction; 4 = definite memory and/or executive dysfunction; 5 = some decline in basic activities of daily living (ADLs); 6 (a-e) definite decline in basic ADLs, with the final stage (e) being fecal incontinence; 7 (a-e) progressive loss of speech and motor abilities, with the final stage (e) being loss of the ability to hold up the head independently.[8]

*Figure Recall Score:* Subject is asked to copy three simple figures, and after delay, is asked to reproduce the figures from memory. The score is the number correctly reproduced.

*Rey Auditory Verbal Learning Test (AVLT):* evaluates short-term auditory-verbal memory, rate of learning, learning strategies, retroactive, and proactive interference, presence of confabulation, of confusion in memory processes, retention of information, and differences between learning and retrieval. Participants are given list of

15 unrelated words repeated over five trials and asked to repeat as many words as possible. After the five trials, the number of recalled words is summed as "AVLT Total Learning Score" (0-75 scale). After the fifth learning trial, another list of 15 unrelated words is provided. The participant recalls as many words as possible from this distracter list. After a brief delay, the participant is asked to repeat the original list of 15 words (AVLT A6).[9, 10]
